# Integrating the environmental and genetic architectures of mortality and aging

**DOI:** 10.1101/2023.03.10.23286340

**Authors:** M. Austin Argentieri, Najaf Amin, Alejo J. Nevado-Holgado, William Sproviero, Jennifer A. Collister, Sarai M. Keestra, Aiden Doherty, David J. Hunter, Alexandra Alvergne, Cornelia M. van Duijn

## Abstract

It has long been suggested that environmental exposures (i.e., the exposome) play a dominant role in shaping trajectories of human aging and premature mortality. Here we aimed to quantify the contribution of the exposome and genome to aging and mortality. We conducted an exposome-wide analysis in the UK Biobank (n=492,567) to systematically identify exposures associated with mortality while accounting for exposure correlation and mismeasurement. We found that the exposome is a major mortality determinant irrespective of genetic disease risk via shaping distinct biological and multimorbidity patterns. We identified 41 independent exposures associated with mortality, and demonstrate that most identified exposures are associated with a common signature of age-related multimorbidity, aging biomarkers, and major cardiometabolic risk factors. Compared with age and sex, polygenic risk for 22 major diseases and aging phenotypes explained an additional 2% of mortality variation, whereas the exposome explained an additional 19%. While genetics explained the majority of variation in dementias and breast, prostate, and colorectal cancers, the exposome explained the majority of variation for diseases of the lung, heart, and liver. Our findings provide a comprehensive map of the contributions of environment and genetics to mortality and common age-related diseases.

## Main

Human aging is a complex process that initially manifests as sub-clinical and biological changes that begin to accumulate from mid-life onwards ^1-3^. These systemic biological changes are major drivers of common age-related diseases ^4-6^ and disease multimorbidity ^7,8^, which in turn are the major causes of premature mortality worldwide ^9^. While there have been major advancements in understanding the complex genetic etiology of age-related diseases, genetic studies show only a modest effect of the genome on lifespan ^10,11^. Instead, the nearly twofold increase in global human lifespan during the past 200 years ^12^ has been largely attributed to changes in human environments ^13^. While epidemiological research has made progress in relating individual environmental and behavioral exposures to age-related diseases and mortality, few studies have comprehensively examined the exposome (i.e., the total set of interrelated environmental exposures throughout the lifecourse) in relation to these outcomes ^14,15^. In the field of genetic epidemiology, the use of genome-wide approaches has greatly increased the positive predictive value ^16^ and reproducibility ^17^ of findings. Transitioning to exposome-wide study designs will provide similar advancements.

We conducted an exposome-wide analysis using data from the UK Biobank (n=492,567) to systematically identify exposures associated with mortality and multiple stages of the aging process. We first systematically identified exposures that associate with the most critical outcome of all age-related diseases – mortality. Exposures associated with mortality were then tested in relation to: (i) incidence of age-related diseases that are either major causes of death or highly prevalent in aging populations (25 total); (ii) cross-sectional patterns of all age-related blood biomarkers available in the UK Biobank (25 total); and (iii) prevalence of three major cardiometabolic risk factors (obesity, hypertension, dyslipidemia). Lastly, we used publicly available polygenic risk scores to quantity the relative contribution of the exposome vs. polygenic risk to explaining variation in mortality and age-related diseases.

### Mortality and age-related disease rates

The final study sample included 492,567 UK Biobank participants (Fig. 1). All analyses were carried out using UK Biobank participants recruited in England (n=436,891). Participants recruited in Scotland/Wales (n=55,676) were held out as a validation set used only to validate final multivariable disease models. There were 31,716 deaths from all causes among participants recruited in England after a median 12.5 years of follow up (Table S1). The majority (74.5%) of deaths were premature deaths (i.e., occurring before 75 years of age; Fig. 2a) and 75% of deaths occurred in those who were overweight or obese with a body mass index (BMI) ≥ 25 kg/m^2^ (Fig. 2b). Women had a lower all-cause mortality rate compared with men (5.4% in women vs 9.4% in men; Table S1). Mortality by cause of death for all participants is given in Tables S4-S5.

**Fig. 1.**
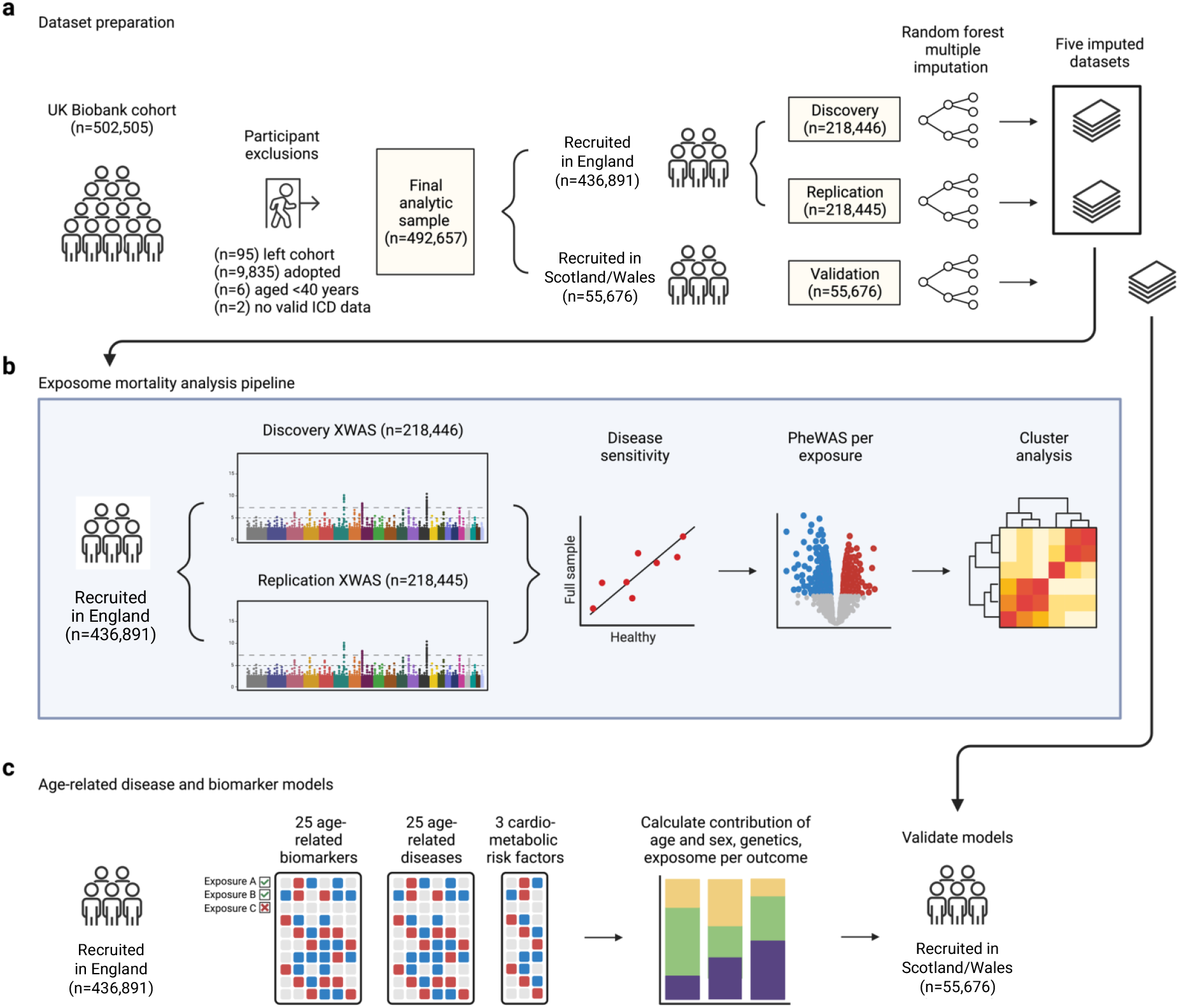
Study overview. **(a)** After participant exclusions, UK Biobank participants were split into independent discovery, replication, and validation sets. Missing values were imputed separately within each group using random forest multiple imputation, resulting in 5 imputed datasets for each dataset. **(b)** Among UK Biobank participants recruited in England (n=436,891), an exposome-wide association study (XWAS) for all-cause mortality was conducted using the discovery and replication sets. The discovery and replication sets were then pooled, and further analyses were conducted in the full sample to identify and remove replicated exposures that are sensitive to reverse causation (disease sensitivity), mismeasurement (PheWAS per exposure), and correlation bias (cluster analysis). **(c)** Exposures surviving all analyses in **(b)** were then tested in relation to 25 age-related biomarkers, 25 age-related diseases, and 3 cardiometabolic risk factors (hypertension, obesity, dyslipidemia). For mortality and each age-related disease, the relative contributions of age and sex, polygenic risk, and exposome were calculated via multivariable Cox proportional hazards models. Multivariable models were validated in participants recruited in Scotland/Wales (n=55,676), who were held out from all other analyses.

**Fig. 2.**
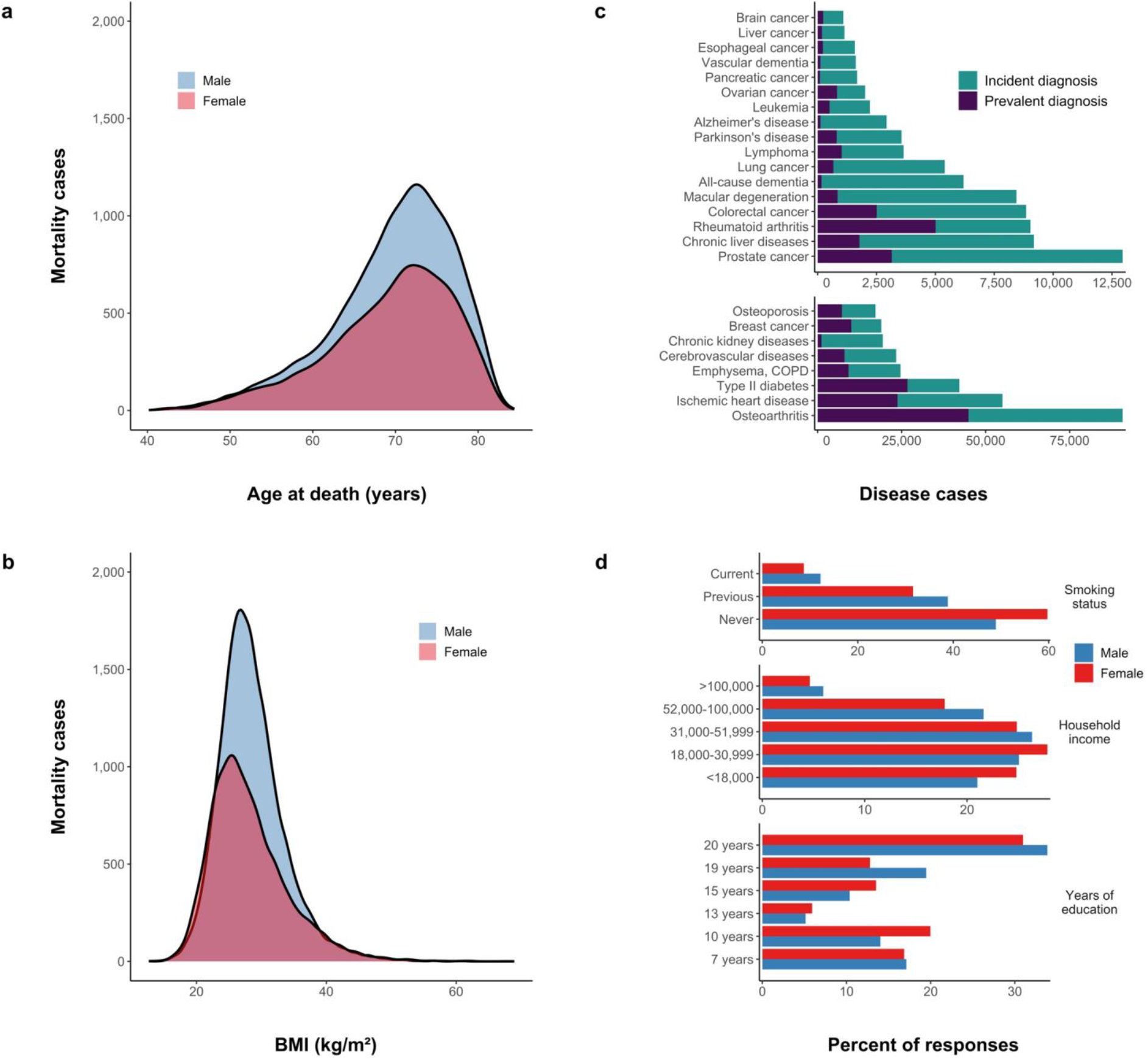
Mortality and key exposure response rates among UK Biobank participants. **(a)** The number of deaths in women and men according to age at death (in years). **(b)** The number of deaths in women and men according to body mass index (BMI) at baseline. **(c)** Numbers of prevalent and incident cases for all age-related diseases studied. Note that diseases are put into two groups with different x-axis scales, since some diseases had far more cases than others. **(d)** Response rates for key exposures and covariates in both women and men. Percentages are for each sex separately and not across both sexes. All descriptive statistics are for UK Biobank participants recruited in England (n=436,891).

The number of incident cases for all age-related diseases studied in participants recruited in England ranged from 856 (brain cancer) to 45,879 (osteoarthritis), as shown in Fig. 2c and Table S6; summary statistics for all cross-sectional outcomes (3 cardiometabolic risk factors, 25 baseline aging biomarkers) are given in Tables S6-S7. Key demographic prevalence rates for participants recruited in England are shown in Fig. 2d.

### Exposome-wide analysis of mortality

Exposome-wide association study (XWAS) analyses of all-cause mortality were conducted by serially testing 164 environmental exposures in relation to mortality via Cox proportional hazards models using independent discovery and replication subsets of the UK Biobank study population (Fig. 1). No significant differences were observed in XWAS regression betas when calculated separately in women and men (Fig. 3a). In a final mortality XWAS combining women and men, 110/164 exposures (67.1%) were significantly replicated (Fig. 3b). Smoking, renting public housing (compared with home ownership), Townsend deprivation index, and living with a partner were the exposures most strongly associated with mortality. Sensitivity analyses (i) excluding participants who died within the first 4 years of follow up and (ii) testing interactions between each exposure and a baseline poor health indicator suggested that largely there is no strong statistical evidence for reverse causation bias in our XWAS results (Fig. S1-S2), with only 15 exposures identified whose associations with mortality were likely completely explained by prevalent disease status (Fig. S1). These exposures were discarded, leaving 95 remaining exposures. Summary statistics from all mortality XWAS analyses are given in Supplementary Files SF3-SF7.

**Fig. 3.**
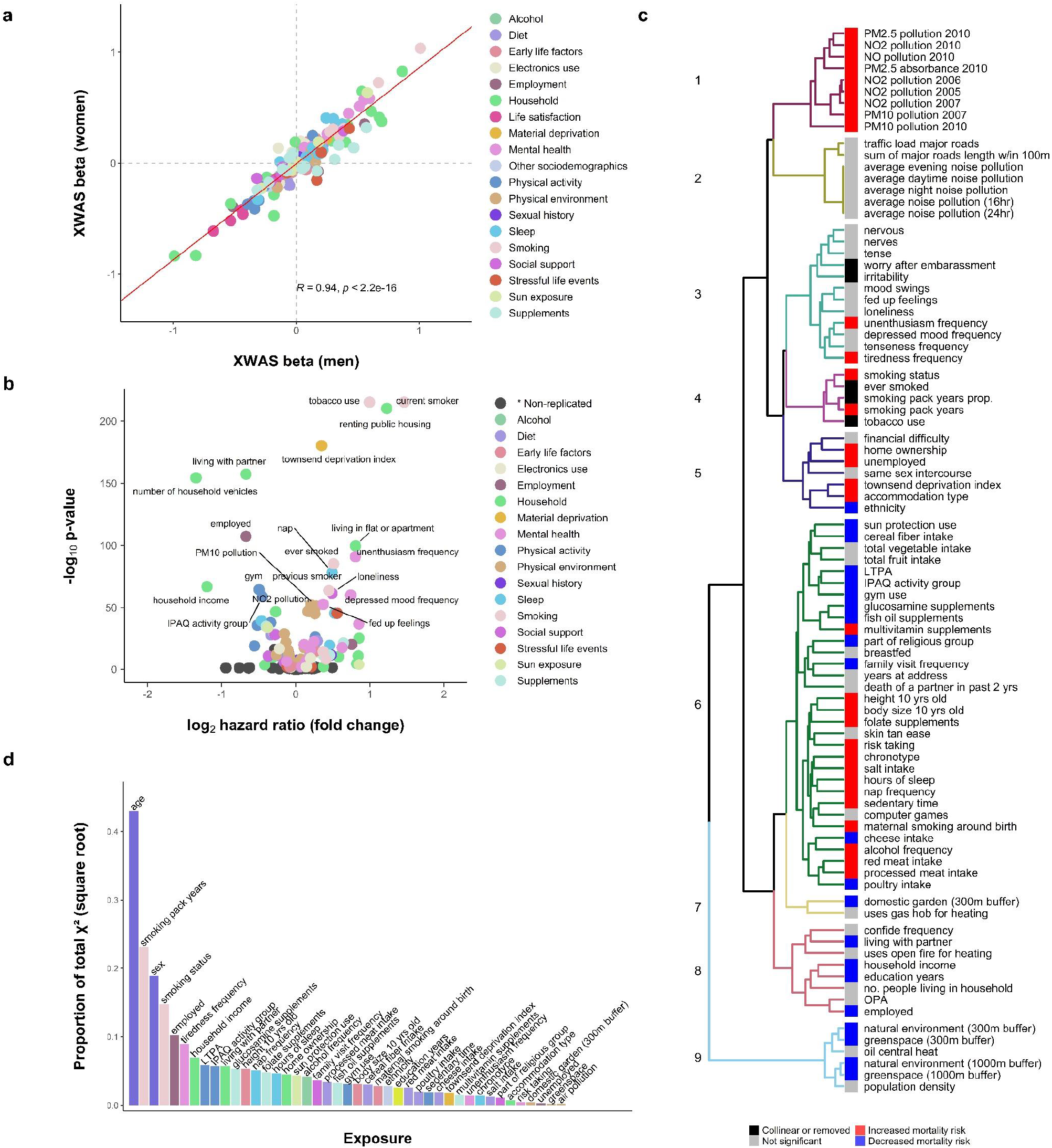
Environmental architecture of mortality in the UK Biobank. **(a)** Correlation between betas for the association between each exposure and mortality calculated separately in women (n=237,637) and men (n=199,257). **(b)** Volcano plot of log-transformed p-values and fold change (calculated as log_2_ of the hazard ratio) for all mortality XWAS associations in the final pooled analysis. Each point represents the effect and p-value for the association between a single exposure and all-cause mortality from the XWAS discovery analysis (n=218,483). Exposures that were FDR significant in both the discovery and replication stages are colored, whereas associations that were not replicated are colored dark grey and grouped in the category “* Non-replicated.” The top 20 points according to strongest p-value are labelled. **(c)** Cluster structure of exposures that were replicated in the mortality XWAS and not discarded during reverse causation and PheWAS sensitivity analyses. Heatmap along the cluster dendrogram shows the effect direction and significance of each exposure in the final cluster multivariable model, with exposures that were not significant at the level of p<0.05 in the cluster multivariate models colored grey. Note that while individual exposures are shown, exposures in clusters 1, 2, and the greenspace/natural environment exposures in cluster 9 were reduced to principal components for multivariable modeling due to extremely high correlation (> 0.90) among similar exposures. **(d)** Individual exposure importance from a multivariable model including age, sex, and all 41 exposures identified in cluster modeling (n=436,891). Variable importance was calculated using a Wald test from ANOVA, where the importance of each variable is the proportion of that variable’s Wald chi-squared (Χ^2^) relative to the total model Χ^2^. Note: the y-axis values were transformed by taking the square root to improve visualization. IPAQ: International Physical Activity Questionnaires; LTPA: leisure time physical activity; MH: mental health; OPA: occupational physical activity.

### PheWAS of replicated exposures

Each exposure replicated in the mortality XWAS and passing the above sensitivity analyses was checked for possible collinearity with other exposures and mismeasurement by conducting a phenome-wide association study (PheWAS) where the replicated exposure was treated as the outcome variable and regressed against all baseline phenotypes present in the UK Biobank using either logistic or linear regression (Supplementary Information). Using this method, we detected a further 10 exposures that associated extremely strongly with either: (i) disease, frailty, or disability phenotypes; or (ii) another exposure such that it likely does not represent new information. For example, we found that the number of vehicles in a participant’s household was very strongly associated with greater household income (Beta: 1.1, p < 8.1×10^−12^), while inversely associated with living in council housing vs. home ownership (Beta: -0.98, p < 5×10^−56^) and being unemployed due to a disability (Beta: -0.62, p < 1.4×10^−245^). These findings indicate that this exposure is mostly capturing socioeconomic and disability status (Fig. S5). Exposures showing mismeasurement from PheWAS were discarded and not carried forward to further analyses, leaving 86 remaining exposures. Summary statistics from all PheWAS are given in Supplementary Files SF62-SF177.

### Cluster architecture of the exposome

We observed high degrees of correlation between exposures replicated in the XWAS (90% of variable pairs had a significant Bonferroni-corrected correlation p-value below 0.001), indicating that some mortality associations observed in the XWAS may be confounded due to this correlation structure. To address this, we used hierarchical clustering to organize replicated exposures that passed all sensitivity analyses into 9 unique clusters (Fig. 3c). We first conducted multivariable mortality models within each cluster by adding all exposures from the cluster into a single Cox model (Fig. S11a). We discarded exposures that did not pass multicollinearity tests or were not significant in this within-cluster model. We then grouped the remaining significant cluster exposures into two large superclusters (clusters 1-5 and 6-9) and conducted a Cox mortality model within each of these superclusters to also account for long-range correlation confounding. Using this method, we identified 41 exposures that remained significant in these cluster multivariable models (Fig. S11b).

### Patterns of age-related multimorbidity and biological mechanisms

To test whether the 41 identified exposures were associated with not just mortality but also multiple stages of the aging process, we tested each exposure individually in relation to incidence of 25 age-related diseases via Cox proportional hazards models (8-15 years of follow-up), as well as cross-sectional patterns of 25 age-related biomarkers and 3 cardiometabolic risk factors (hypertension, obesity, dyslipidemia) via linear and logistic regression, respectively. Each of the 41 exposures was associated with a wide range of aging biomarkers that span diverse organ systems and mechanisms (Fig. 4a). On average, each exposure was associated with 21.9 biomarkers (out of 25). Four exposures were associated with all 25 biomarkers (Townsend deprivation index, smoking status, hours of sleep, ethnicity) and nine with 24/25 biomarkers in total (unenthusiasm frequency, total sedentary time, home ownership, taking multivitamin supplements, leisure time physical activity [LTPA], household income, years of education, relative body size at 10 years old, alcohol intake frequency). Cardiometabolic risk factors studied were cross-sectionally associated with nearly every exposure studied (Fig. 4b). Notably, hypertension was cross-sectionally associated with all exposures tested.

**Fig. 4.**
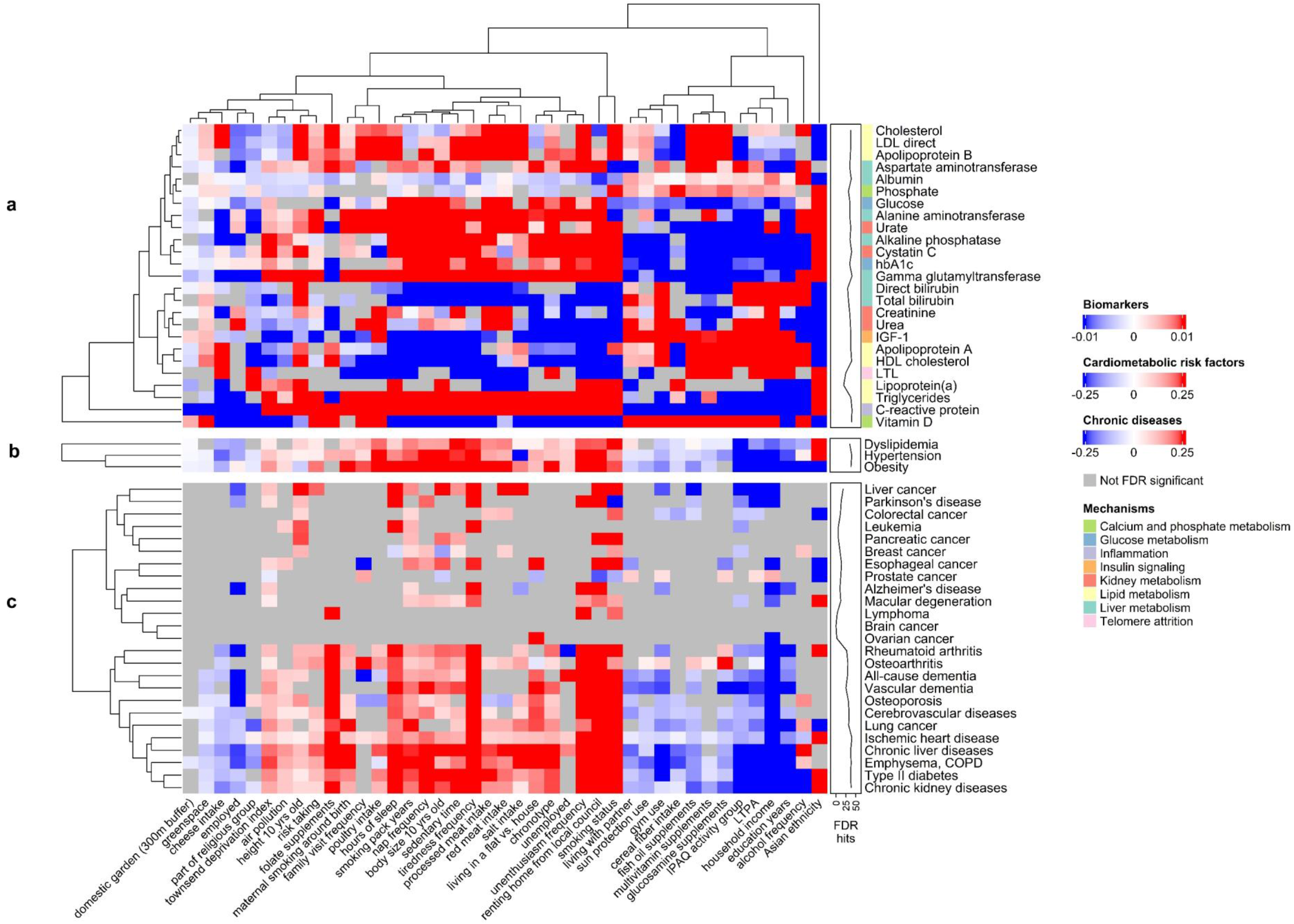
Environmental architectures of age-related biological mechanisms and diseases in the UK Biobank. **(a)** Associations between each mortality-associated exposure and aging biomarkers. **(b)** Associations between each mortality-associated exposure and cardiometabolic risk factors. **(c)** Associations between each mortality-associated exposure and age-related chronic diseases. Colors in the heatmaps represent betas for associations between exposures and biomarkers/diseases. A line annotation track is shown that counts the total number of FDR significant associations for each outcome. For heatmap **(a)**, an addition annotation track shows the primary biological mechanism associated with each aging biomarker. Air pollution and greenspace are the first principal components combining all air pollution and greenspace variables, respectively. For nominal categorical variables with more than one response level, the association for the level with the strongest p-value is reported in this figure and the exposure’s label reflects the response category shown. COPD: chronic obstructive pulmonary diseases; IGF-1: insulin-like growth factor 1; LTL: leucocyte telomere length; LTPA: leisure time physical activity.

Each of the 41 exposures was also prospectively associated with concurrent incidence of multiple age-related diseases (Fig. 4c), indicating that each exposure is a likely catalyst of disease multimorbidity. On average, each exposure was associated with 11.7 age-related diseases (out of 25). Smoking was associated with 21 (current smoking status) and 20 diseases (pack years). Household income and home ownership were associated with 19/25 diseases, followed by tiredness frequency and Townsend deprivation index (18/25), and IPAQ physical activity group (17/25). Of note, we found no associations between any exposure and incidence of brain cancer.

Approximately 75% of exposures were associated with a consistent biological signature and multimorbidity pattern. For example, exposures associated with decreased mortality risk (employment, household income, education, living with a partner, IPAQ, LTPA, gym use, fish oil supplements, cereal fiber intake) were associated with decreased risk of nearly two thirds of all age-related diseases studied; decreased risk of obesity, dyslipidemia, and hypertension; increased levels of biomarkers indicating better health (vitamin D, HDL cholesterol, phosphate, albumin) or slower aging (LTL); and decreased levels of detrimental biomarkers (C-reactive protein, blood glucose, hbA1c, cystatin C, triglycerides). Exposures associated with increased mortality risk (smoking, lack of home ownership, being unemployed, deprivation, air pollution, maternal smoking around birth, sedentary lifestyle, poor mental health, poor sleep, processed and red meat intake) were generally associated with opposite patterns of these same diseases, risk factors, and biological mechanisms. Summary statistics from all biomarker, age-related disease, and cardiometabolic risk factor analyses are given in Supplementary Files SF8-SF60.

### Environmental and genetic architectures of mortality and age-related diseases

To determine the contribution of age and sex, exposome, and genetic risk in describing variation in mortality and each of the studied age-related diseases, we calculated stepwise multivariable Cox models beginning with just age and sex (model 1), then adding either polygenic risk scores (PRS) for the outcome (model 2) or exposome (i.e., all exposures associated with the outcome; model 3), and finally adding both the exposome and PRS together (model 4). Models were first calculated among participants recruited in England (n=436,891) and then validated in participants recruited in Scotland/Wales (n=55,676).

Compared with a model containing age and sex, we found that adding PRS for 22 diseases that are either major causes of death or aging phenotypes only increased the total mortality model R^2^ by 2% (Fig. 5a; Tables S15-S16). By contrast, we found that adding all 41 exposures associated with mortality (i.e., exposome) to age and sex increased the total mortality model R^2^ by 17-19% (model 3 vs. 1). Adding the mortality exposome to the model with age, sex, and all PRS increased the total mortality model R^2^ by 16-17% (model 4 vs. 2). While the combined effect of the exposome explained a large proportion of mortality variation, we found that individually most exposures only explained a small proportion of total mortality variation (Fig. 3d). Effect estimates from model 3 for all exposures shown in Fig. S11c for mortality and Fig. S20-S44 for all age-related diseases studied. Exposure importance plots for all diseases studied are shown in Fig. S11-S19.

**Fig. 5.**
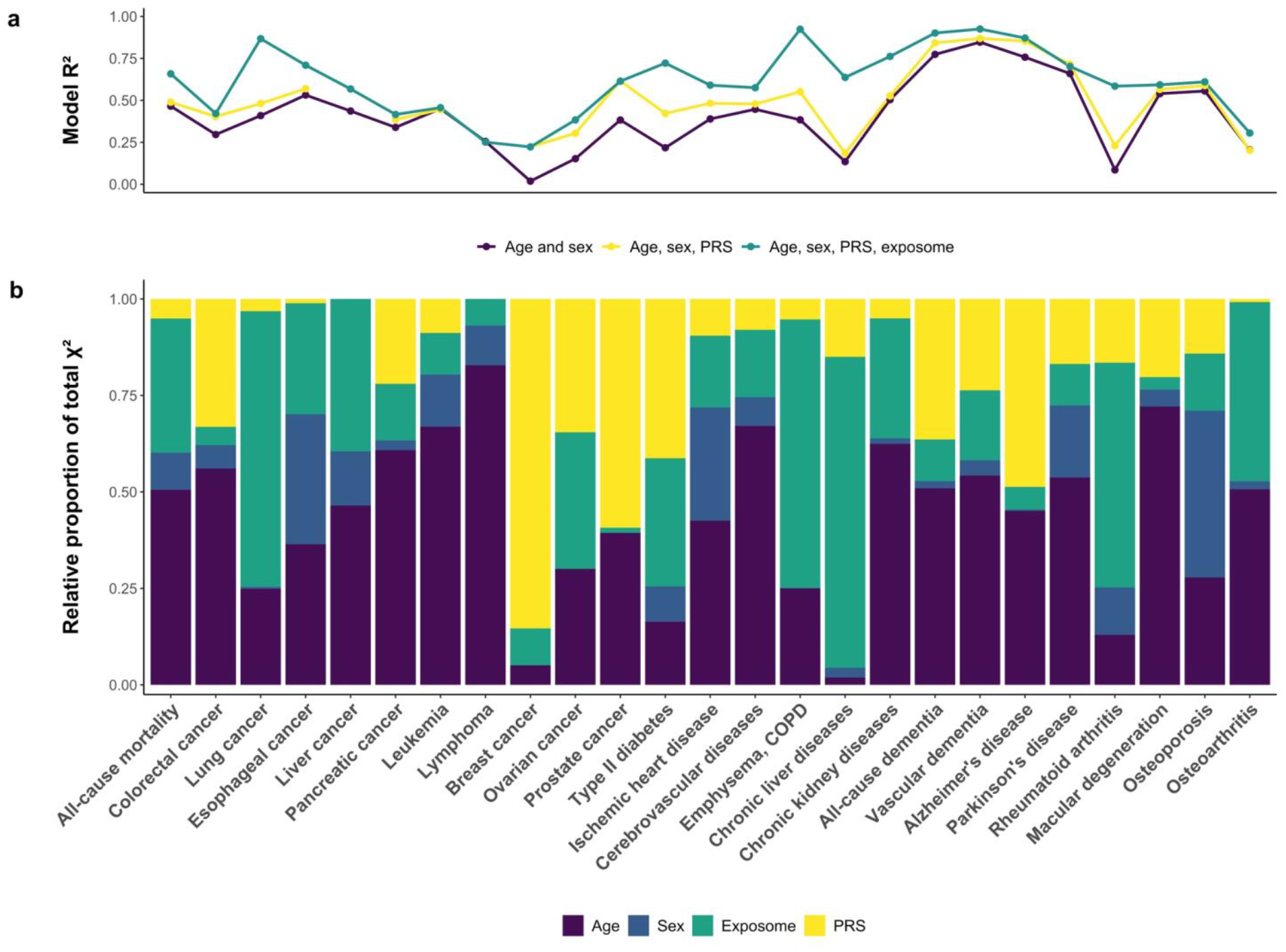
Combined environmental and genetic architectures of mortality and age-related diseases. **(a)** R^2^ calculated across studied outcomes for several sequential multivariable models: model 1 containing age and sex (purple); model 2 containing age, sex, and polygenic risk scores (PRS; yellow); and model 4 containing age, sex, PRS, and exposome (green). If a PRS was not available for a particular outcome, then the green R^2^ shows the results from model 3 (age, sex, exposome). R^2^ values are shown from the validation analyses (n=55,676). **(b)** Variable importance for age, sex, polygenic risk, and exposome for all outcomes studied in model 4 conducted among UK Biobank participants recruited in England (n=436,891). Variable importance is calculated as the proportional of the total model chi-squared (Χ^2^) that each variable category explains and is plotted as the relative contribution to the total model Χ^2^ for each category so that they sum to 1. In all analyses, PRS includes polygenic risk scores, as well as genetic principal components and genotyping batch. PRS used for mortality models includes PRS for all other diseases and phenotypes shown in this figure (22 total). Note: PRS information was not available for liver cancer or lymphoma and is not included in models. Ovarian, breast, and prostate cancer models were sex-specific and sex was not included in model 4 for these outcomes.

Models including age and sex, exposome, and PRS (model 4) captured >50% of variation in most outcomes studied, with the exception of colorectal cancer, pancreatic cancer, leukemia, breast and ovarian cancers, and osteoarthritis (Fig. 5a). For all-cause mortality and all age-related diseases studied, the relative importance of age, sex, exposome, and PRS are shown in Fig. 5b according to the relative proportions of the total model chi-squared (Χ^2^) that each variable category explained in model 4. The exposome explained the most amount of model variation for lung cancer, emphysema/COPD, chronic liver diseases, and rheumatoid arthritis. Certain outcomes seem to be more influenced by polygenic risk than the exposome, such as breast and prostate cancers, Alzheimer’s disease (AD), all-cause dementia, macular degeneration, and colorectal cancer. Of note, ovarian cancer and type 2 diabetes showed a smaller contribution of age and sex, with the exposome and PRS explaining the majority of variation in equal parts. Lastly, a number of outcomes showed age and sex as the most influential determinants, but also showed the exposome explaining the majority of the residual variation not explained by age and sex. These include all-cause mortality, esophageal cancer, ischemic heart disease, and cerebrovascular diseases.

## Discussion

This study provides the first comprehensive assessment of the relative contributions of environmental and genetic influences to aging and mortality. We show that the exposome explains a large percentage of mortality variation beyond the contribution of age, sex, and polygenic disease risk. We further demonstrate that the exposome shapes mortality risk through influencing a common signature of aging biological mechanisms and disease multimorbidity. For most age-related disorders studied, we find that the effect of the exposome exceeds that of the genome. We also find that exposome risk is composed of many interrelated factors that, although individually may have small effects, when combined additively explain a substantial amount of variation for mortality and certain age-related diseases such as emphysema/COPD, lung cancer, chronic liver and kidney diseases, and rheumatoid arthritis.

Our results demonstrate that many age-related diseases share a common environmental etiology that drives disease multimorbidity and ultimately premature mortality. We found that generally diseases fell into two large blocks. The first included cardiometabolic diseases, lung and kidney disease, dementia, vascular dementia, osteoporosis, osteoarthritis, and rheumatoid arthritis, all of which showed a shared environmental etiology involving most exposures associated with mortality. The second block included most common cancers and neurodegenerative disorders (AD, Parkinson’s disease, macular degeneration), which were associated with fewer exposures but still showed similar directions of effect as the first block.

The group of diseases with a small observed impact of the exposome relative to the genome in our study were neurodegenerative disorders and some cancers (breast, prostate, and colorectal). Of note, although we saw associations for known determinants of colorectal cancer such as physical activity, smoking, and intakes of cereal fiber, processed meat, and red meat, we found that the combined contribution of these exposures to explaining colorectal cancer variation was limited compared to polygenic risk.

Throughout our analyses, measures of physical activity, smoking, and individual socioeconomic status (household income and home ownership) showed the strongest effects on mortality and were associated with the greatest number of age-related diseases and aging biomarkers. While numerous previous studies have documented the significant roles of these exposures in shaping mortality risk ^18-21^, we provide a more complete picture of the myriad biological mechanisms and disease pathways associated with each.

Several findings ask for further research. We observed that frequency of alcohol intake was unexpectedly associated with decreased risk of diabetes, ischemic heart disease, and chronic kidney disease. However, a previous Mendelian randomization (MR) study shows an association of alcohol intake with increased risk of cardiometabolic diseases in the UK Biobank ^22^. Further, while glucosamine supplementation was associated in our study with decreased risk of cardiometabolic diseases and with cardiometabolic disease mechanisms (lower glucose, hbA1c, and triglycerides; higher HDL and total cholesterol), previous clinical trials have reported no effect of oral glucosamine on blood pressure, glucose, and lipids ^23^. Further mechanistic or MR studies will be needed to disentangle whether glucosamine intake has a causal effect on these outcomes or instead whether our results reflect the fact that those who take glucosamine are also likely to have higher socioeconomic status and less likely to be sick or disabled at baseline, as shown in the glucosamine PheWAS (Supplementary File SF98).

There are several limitations to note for our analysis. First, despite our prospective study design, we are unable to make causal conclusions based on our findings. Although consistent association patterns observed across different layers of outcomes (mortality, biomarkers, cardiometabolic risk factors, incident diseases) suggest that many exposures we identified may play early roles in shaping age-related disease and mortality risk, causality will need to be verified with further methods such as MR. Second, exposome influences are dynamic over time and our study design cannot capture this dynamic aspect of the exposome since all exposures were only measured at one time point in the full cohort. We also have not captured all exposome influences, as we were limited to the exposures available in the UK Biobank. Conspicuously absent from our analysis are chemical and toxicological exposures beyond air pollution. Furthermore, many exposures we tested come from self-reported questionnaire data, which introduces potential recall bias, mismeasurement, and uncertainty into the reliability and accuracy of the responses. Finally, the use of existing PRS as proxies for the inherited genetic component of each disease is somewhat preliminary, as these are still being updated and improved and do not include the component of rare variation in single genes such as BRCA1/2 or the genes for familial hypercholesterolemia.

Despite these limitations, we believe that our approach offers many advantages over traditional single exposure approaches in epidemiology. Through the use of: (i) independent discovery, replication, and validation stages; (ii) exposome-wide significance thresholds; (iii) sensitivity analyses to test for reverse causation bias; (iv) a comprehensively phenotyped cohort in which we could systematically conduct PheWAS to test for exposure mismeasurement; and (v) within-cluster multivariable models to test for correlation bias, our approach greatly increases the reproducibility and positive predictive value of findings. This study design improves substantially upon XWAS and exposome analyses published to date (see our systematic review in the Supplementary Information). When compared with the only previously published “environment-wide” analysis of mortality ^24^ that focused on a narrower range of chemical and lifestyle exposures in a small sample (n=6,008), our study identified approximately 17x more factors associated with all-cause mortality and improved the final mortality variance explained (R^2^) by 31x from 2.1% in this previous study to 66%. This demonstrates the importance of using large datasets and testing as broad a range of exposome influences as possible.

Overall, our results indicate that environment-focused interventions are likely to have the highest impact on ameliorating premature mortality and most age-related morbidity. We argue that greater use of exposome study designs will significantly accelerate identification of high-priority population health targets for age-related morbidity and premature mortality. Our study also opens the door for further targeted proteomic, metabolomic, or other ‘omics studies to explore the biological effects of the exposures that we identified.

## Methods

### Study design and participants

The UK Biobank is a prospective cohort study with extensive genetic and phenotype data available for 502,505 individuals resident in the United Kingdom ^25^. The full [UK Biobank protocol](https://www.ukbiobank.ac.uk/media/gnkeyh2q/study-rationale.pdf) is available online.

### Exposures

We considered as potential XWAS exposures all non-genetic variables available as of July 24, 2020 that were collected at baseline and were available for participants recruited across all assessment centers. After all exclusions, recoding, and quality control (Supplementary Information), 176 unique exposures remained that were available in the full cohort that were common to both women and men. All continuous exposure variables were centered and standardized before analysis, except for age at recruitment. All ordinal categorical variables were recoded to only test linear associations and other polynomial contrasts (e.g., quadratic or cubic associations) were not assessed. All nominal categorical exposures were analyzed with the most common category set as the reference. Detailed data dictionaries including all exposures used in imputation and XWAS steps are included in Supplementary Files SF1-SF2.

### Outcomes

Detailed information about the [linkage procedure](http://biobank.ctsu.ox.ac.uk/crystal/refer.cgi?id=115559) with national registries for mortality and cause of death information is available online. Mortality data were accessed from the UK Biobank data portal on May 4, 2022, with a censoring date of September 30, 2021 or October 31, 2021 for participants recruited in England/Scotland or Wales, respectively (11-15 years of follow-up).

Aging biomarkers (Table S8) were measured using baseline non-fasting blood serum samples as previously described ^26^. Data on leukocyte telomere length (LTL) was only available in a slightly smaller sample (n=472,506) than other biomarkers and was not imputed. Biomarkers were previously adjusted for technical variation by the UK Biobank, with [sample processing](https://biobank.ndph.ox.ac.uk/showcase/showcase/docs/serum_biochemistry.pdf) and [quality control](https://biobank.ndph.ox.ac.uk/showcase/ukb/docs/biomarker_issues.pdf) procedures described on the UK Biobank website.

Data used to define chronic diseases and cardiometabolic risk factors are outlined in Table S9. Incident chronic disease diagnoses were ascertained using ICD diagnosis codes and corresponding dates of diagnosis taken from linked hospital inpatient records and death register data. ICD data were accessed from the UK Biobank data portal on May 30, 2022, with a censoring date of September 30, 2021; July 31, 2021; or February 28, 2018 for participants recruited in England, Scotland, or Wales, respectively (8-15 years of follow-up).

### Missing data imputation

UK Biobank participants recruited from England were randomly assigned to a discovery (n=218,446) or replication set (n=218,445) while maintaining the same proportion of mortality cases in each. We performed missing data imputation separately in the discovery, replication, and Scottish/Welsh validation (n=55,676) datasets using the R package missRanger ^27^, which combines random forest imputation with predictive mean matching (Supplementary Information). We imputed 5 datasets, with a maximum of 10 iterations for each imputation. All subsequent study analyses were run independently in each of the five imputed datasets, and results were pooled using Rubin’s rule ^28^.

### Exposome-wide association study

XWAS of all-cause mortality were initially carried out separately in women and men, and then a final XWAS was calculated in the pooled dataset with both women and men to increase power. Exposures in the final pooled XWAS were limited to those asked to both women and men, omitting sex-specific reproductive factors. In each XWAS, we serially assessed associations of each individual exposure with all-cause mortality using Cox proportional hazards models with age as the time scale; stratified by 5-year birth cohorts and sex (in the pooled analysis only); and adjusted for assessment center, years of education (7 years, 10 years, 13 years, 15 years, 19 years, 20 years), and ethnicity (White, Asian, Black, Mixed, Other). Since it has been shown that UK Biobank participants are likely to misreport alcohol consumption as a function of higher disease burden ^29^, self-reported overall health status was added as an additional XWAS covariate for the self-reported alcohol intake exposure only. P-values in the discovery and replication analyses were corrected using the false discovery rate (FDR; Benjamini-Hochberg method^30^) with a significance threshold of FDR p < 0.05. After completing the mortality XWAS, discovery and replication sets were recombined into the full English sample (n= 436,891) to complete further sensitivity analyses (Supplementary Information).

### Correlation and cluster analyses

Correlation between all variables was calculated in the full English sample using the R package polycor ^31^ to create a heterogenous correlation matrix for each imputed dataset (Supplementary Information). We used hierarchical clustering via Euclidean distance to identify the cluster structure of exposures replicated in the pooled XWAS and not susceptible to reverse causation bias (plus education and ethnicity). Within-cluster Cox multivariable mortality models included all remaining variables in the cluster after removing collinear variables (Supplementary Information), with additional adjustment for assessment center, household income (Less than 18,000, 18,000 to 30,999, 31,000 to 51,999, 52,000 to 100,000, Greater than 100,000), Townsend deprivation index, years of education, and ethnicity (if those variables were not already in the cluster). The significance threshold used in cluster multivariable analyses was a nominal p < 0.05.

### Aging mechanisms and incident chronic disease analyses

Aging biomarker variables were log transformed and then were age-adjusted by regressing each onto age at recruitment separately in women and men. Across exposures replicated in the XWAS and passing all sensitivity tests, we serially assessed associations between each exposure and age-adjusted biomarker using cross-sectional linear regression models with covariates for sex, 5-year birth cohort, assessment center, years of education, ethnicity, number of medications, smoking status (current, previous, never), and IPAQ physical activity level (low, moderate, high). IGF-1, LTL, and vitamin D models included additional covariates for standing height (in cm), leukocyte count (10^9^ cells/Liter), and month of biomarker assessment (to control for seasonality of sun exposure), respectively.

For chronic disease analyses, we serially assessed associations between each exposure and incident disease using the Cox proportional hazards model, with all XWAS covariates plus household income, smoking status, and IPAQ physical activity group. Sex-specific reproductive exposures (e.g., menopause) replicated in the female- and male-only XWAS analyses were also tested as exposures in analyses of sex-specific chronic disease outcomes (breast, ovarian, and prostate cancer).

For cardiometabolic risk factors (obesity, hypertension, dyslipidemia), we serially assessed each exposure and risk factor pair using cross-sectional logistic regression models adjusted for age, sex, assessment center, household income, years of education, ethnicity, smoking status, and IPAQ physical activity level.

Across all biomarker, chronic disease, and cardiometabolic risk factor analyses, p-values were corrected separately for each outcome using FDR.

### Exposome and polygenic risk multivariable models

For each outcome, five multivariable models were calculated. The first only includes age (scaled) and sex in the model (model 1). Model 2 includes age, sex, and the polygenic risk score (PRS) for the outcome, if available (see below for more detail). Model 3 includes age, sex, and all exposures associated with the outcome (exposome). Model 4 includes age, sex, exposome, and PRS. If a PRS was not available for a particular outcome, then models 2 and 4 were not calculated for that outcome. Each model was validated in the independent Scottish/Welsh dataset (n=55,676) by obtaining the linear predicted values from the models in the English dataset and measuring the C-index and R^2^ for these values in relation to the outcome rates in the Scottish/Welsh population.

In all multivariable Cox models, the proportional hazards assumption was tested by examining the Schoenfeld residuals, and an interaction with time was added to any variable with non-proportional hazards (Supplementary Information). Relative importance for each variable and category of variables within the multivariable models was calculated using Wald chi-squared (Χ^2^) statistics via ANOVA, where the relative importance of each is the proportion of the variable/group Χ^2^ relative to the total model Χ^2^.

## Data Availability

UK Biobank data are available through a procedure described at https://www.ukbiobank.ac.uk/enable-your-research.

## Acknowledgements

We thank Robert Clarke, Roel Vermeulen, Gary Miller, and Timothy Key for critical review of the analyses presented in this manuscript. This research has been conducted using the UK Biobank Resource under Application Number 61054.

## Author contributions

M.A.A., C.M.vD., A.A., and N.A. conceptualized the study. M.A.A. performed all data curation, formal analysis, and data visualization. Data curation and formal analyses were supervised by C.M.vD., A.A., and N.A. Analytical input was provided by W.S. and A.N-H. for clustering analyses. Analytic input for physical activity variables was provided by A.D. Guidance on calculating polygenic risk scores (PRS) was provided by J.C. for diseases not included in the UK Biobank PRS release (pancreatic, lung, esophageal, leukemia, COPD, chronic liver disease, chronic kidney disease, osteoarthritis), under the supervision of D.H. The systematic review was performed by M.A.A. and S.M.K., including independent screening of abstracts/papers. M.A.A. prepared the manuscript, figures, tables, and supplementary files, with edits and revisions provided by all other co-authors. The GitHub code repository was created and is maintained by M.A.A.

## Data and materials availability

UK Biobank data are available through a procedure described at https://www.ukbiobank.ac.uk/enable-your-research. R and PLINK code needed to reproduce all analyses, figures, and tables will be made publicly available on GitHub before journal publication. Summary statistics from all analysis stages are included in Supplementary Files SF3-SF177.

## List of Supplementary Materials

Supplementary Information

Materials and Methods

Sensitivity Analyses

Supplementary discussion of findings in relation to existing research

Supplementary discussion on study limitations and strengths

Fig. S1 – S58

Tables S1 – S16

References (1 – 87)

Supplementary File titles and summaries

Supplementary File

This single Excel file contains Supplementary Files SF1-SF177 (each file is its own tab in the document); see main Supplementary Information PDF for table titles and summaries.

