## Supplementary Information for "Integrating the environmental and genetic architectures of mortality and aging"

Argentieri et al. 2023

### SUPPLEMENTARY INFORMATION

**Supplementary methods**

### 1.1 Study design and participants

The UK Biobank is a prospective cohort study with extensive genetic and phenotype data available for 502,505 individuals resident in the United Kingdom [1]. The full UK Biobank protocol is available online.

### 1.2 Variable selection

We considered for study inclusion all non-genetic variables that were collected at baseline in the UK Biobank via the touchscreen questionnaire, verbal interviews, physical measures, cognitive function assessment, blood sample assays, and urine assays that were available as of July 24, 2020. This included measures of the exposome, as well as variables on disease status, physical and biological measures (e.g., BMI, hand grip strength, blood/urine biomarkers), disease treatment (e.g., medications), and disability/frailty (e.g., disability allowances, walking pace). These non-exposures are referred to collectively as disease morbidity and aging phenotype variables. Exposures were included for use in our exposome-wide association study (XWAS) of all-cause mortality, whereas prevalent and incident diseases, morbidities, and aging phenotypes were included in our datasets both to improve the precision of the multiple imputation and to use later as covariates for analysis.

For certain physical measures, such as vision and hearing tests and heel bone mineral density, only non-specialist summary variables were considered and the rest were excluded. Similarly, the “best measure” variable for both forced vital capacity (FVC) and forced expiratory volume in 1-second (FEV1) were used and the raw readings for each individual blow attempt were not analyzed. Baseline ECG during fitness variables were excluded because no summary measures are currently available. Responses on hormone replacement therapy or oral contraceptives from field ID 6153 were not considered, as more detailed information on these was available in the female-specific reproduction variables. Where multiple variables exist for the same trait, the most complete variable with the least amount of missing data was selected for analysis (e.g., standard measures of weight and BMI were used instead of impedance measures). Variables marked as pilot variables, or related to metrics for collecting other variables, device IDs, or other procedural metrics were also excluded.

Further deprivation variables derived from participants' post codes at baseline were also considered for inclusion. Townsend deprivation index is calculated for each participant based on the national census output areas from the national census immediately preceding their recruitment date into the UK Biobank.

Baseline air pollution measures are available at several different time points preceding or concurrent with the baseline recruitment in the UK Biobank. Air pollution estimates for nitrogen oxides (NO), nitrogen dioxide ($NO_2$), and particulate matter ($PM_{2.5}$, $PM_{10}$) in the years 2005-2007 were derived from EU-wide air pollution maps (resolution 100m x 100m) [2]. Pollution concentration values were mapped to UK Biobank participants by overlaying the x-y geographical coordinates of UK Biobank participants with these maps (projected to British National Grid). The same air pollution measures were also estimated in 2010 for each participant's address using a Land Use Regression (LUR) model developed as part of the European Study of Cohorts for Air Pollution Effects (ESCAPE) [3].

Additionally, several other derived physical environment measures are available at baseline for UK Biobank participants. Traffic variables (e.g., distance to nearest major road, traffic intensity on nearest road) were calculated using LUR based on Eurostreets (vs 3.1) digital road network for the year 2008. Noise pollution for the year 2009 was modelled using a version of the CNOSSOS-EU noise model [4]. Further information on derivation of the air pollution, noise pollution, and traffic variables is available on the UK Biobank website. Greenspace exposure and distance to the coast was mapped to participants based on home location grid references. Greenspace measures were created using the 2005 Generalised Land Use Database for England [5]. Coastal proximity was calculated for participants as a Euclidean distance raster from the coastline for a small grid cell size (50m), which was then mapped to UK Biobank participant coordinate locations. Further information is available on the UK Biobank website. Finally, urban and rural home area classification were mapped to UK Biobank participants by post code using the classifications created by the Office for National Statistics (ONS) as part of the ONS Postcode Directory (ONSPD).

We also considered for inclusion five items administered in the UK Biobank from the Childhood Trauma Screener (CTS), which was developed as a short form of the Childhood Trauma Questionnaire for large epidemiological populations [6]. CTS variables were collected in 2016-17 through a mental health questionnaire administered to UK Biobank participants via online follow-up. All UK Biobank participants with a valid email address on record were invited to complete the questionnaire, of whom 158,835 completed the questionnaire. Valid responses for CTS questions were received for 157,348 participants. These variables were initially considered for inclusion in our analysis as baseline exposures because they were asked retrospective to childhood, however sensitivity analyses showed significant survival bias evident in associations with all-cause mortality using these variables in the general cohort. Although childhood abuse exposures were associated with increased risk of all-cause mortality in just the sub-population administered the mental health questionnaire, analyses in the entire cohort study population (after imputing missing values for childhood trauma for the whole cohort) showed these same exposures to be associated with significantly decreased risk of mortality. This is because these variables were only asked to a subset of the cohort that survived all the way from baseline until 2017, and thus represent responses from a healthier and longer living sub-population compared with the full baseline cohort.

After creating binary dummy variables from any "mark all that apply" questions (this process is described below in the "Variable coding" section), we arrived at a total of 769 potential variables to include in our dataset for imputation. From this list of 769 potential variables, we first excluded all variables missing in more than 80% of our final male and female datasets. Furthermore, we also excluded all nested variables (e.g., a question asking respondents to indicate the number of years since diagnosis that was only asked to participants who responded yes to having a previous cancer diagnosis) except for those that could be recoded to avoid improper missing data imputation (see "Derived variables", below).

Finally, we excluded the questionnaire item on never eating eggs, dairy, wheat, sugar from analyses, as the binary dummy variables derived from these were not reliable in our early testing and suffered from mismeasurement issues. For example, we found that never eating sugar was very strongly associated with having a diabetes diagnosis (OR: 4.2, $p < 2.9 \times 10^{-155}$) and insulin medication intake (OR: 5.2, $p < 1.5 \times 10^{-82}$), indicating that this exposure is mostly capturing diabetes and disability-related dietary restriction (Fig. S4).

After all variable exclusions, including further variable deletions during the pre-XWAS quality control stage (see section "Variable quality control", below), we were left with a final total of 174 and 168 exposures in women and men, respectively, to be included in the XWAS analysis. We were also left with a total of 239 variables in both women and men representing disease morbidity and aging phenotypes. Exposures common to both men and women were

164 in total. Two variables (accommodation type [field ID 670] and diet change over the past 5 years [field ID 1538]) are included in both domains of analysis, as some response levels of these variables were considered indicative of an exposome association (and only included in the XWAS), whereas other response levels were indicative of illness or disability (e.g., diet change in the past 5 years due to an illness, or reporting living in sheltered accommodation). All continuous exposure variables were centered and standardized before analysis, except for age at recruitment. All ordinal categorical variables were recoded to only test linear associations and other polynomial contrasts (e.g., quadratic or cubic associations) were not assessed. All nominal categorical exposures were analyzed with the most common category set as the reference. Detailed data dictionaries including all exposures used in imputation and XWAS steps are included in Supplementary Files SF1-SF2.

#### 1.3 Outcomes

Detailed information about the linkage procedure with national registries for mortality and cause of death information is available online. Mortality data were accessed from the UK Biobank data portal on May 4, 2022, with a censoring date of September 30, 2021 or October 31, 2021 for participants recruited in England/Scotland or Wales, respectively (11-15 years of follow-up).

Aging biomarkers (Table S8) were measured using baseline non-fasting blood serum samples as previously described [7]. Data on leukocyte telomere length (LTL) was only available in a slightly smaller sample (n=472,506) than other biomarkers and was not imputed. Biomarkers were previously adjusted for technical variation by the UK Biobank, with sample processing and quality control procedures described on the UK Biobank website.

Incidence of 25 chronic diseases and prevalence of three cardiometabolic risk factors (hypertension, obesity, dyslipidemia) was assessed in the full population studied in the mortality XWAS analyses (n=436,891). ICD codes used to define each disease are given in Table S9, below. For all incident chronic disease and clinical risk factor analyses, participant follow-up started at recruitment into the UK Biobank study and ended on the censoring date or date of first disease diagnosis, for all participants, leaving a total of 8-15 years of total follow-up. We also searched death register data for participants with one of the diseases listed as a primary or contributory cause of death but no hospital inpatient diagnosis. In this case, the participant was listed as a case and the date of diagnosis used was the date of death. If any participants with no relevant diagnosis died before the censoring date, then date of death was used as the censoring date. ICD data were accessed from the UK Biobank data portal on May 30, 2022, with a censoring date of September 30, 2021; July 31, 2021; or February 28, 2018 for participants recruited in England, Scotland, or Wales, respectively (8-15 years of follow-up).

For each disease, prevalent cases at baseline were identified using the baseline touchscreen data and baseline verbal interview codes listed in Table S9. We also used the ICD hospital diagnosis data to further identify any prevalent cases who had a corresponding first date of diagnosis before or on their date of recruitment into the UK Biobank. Prevalent cases were excluded from analysis of each disease in order to identify true incident cases. The incidence and prevalence rates of each disease and endophenotype studied are shown in Table S6. For any participant with multiple dates of diagnosis for a single disease, the first date of diagnosis was used. Breast, ovarian, and prostate cancer analyses were carried out as sex-specific analyses in female (breast, ovarian) or male (prostate) participants.

### 1.4 Variable coding

The following variable recoding was carried out before multiple imputation. All variable responses of “Prefer not to answer”, “Do not know”, and “Not applicable” were recoded to NA. Following multiple imputation, any variable values where the participant responded “Prefer not to answer” in the original dataset were recoded to NA. All nominal categorical variables were coded as unordered factors, with the reference level set as the most frequent response reported in the UK Biobank dataset. Ordered categorical variables were coded with the reference level set as the lowest response (e.g., “Never”, “Rarely”, or “None”). Dichotomous categorical variables were coded with the reference level set as “No.” All variables with “mark all that apply” response categories were converted into multiple dummy variables, with each unique response option used to create a yes/no dichotomous variable.

For all numeric diet intake variables, participants who responded “less than one” were recoded as 0.5. For variables on number of hours spent watching TV, on the computer, and driving, participants who responded “less than one” were recoded as 0.5.

To harmonize responses to the home area population density variable (field ID 20118) across different countries, this variable was recoded with a simpler classification of urban or rural home area population density. Participants who were classified in the original variable as "England/Wales - Urban – sparse," "England/Wales - Urban - less sparse," "Scotland - Large Urban Area," or "Scotland - Other Urban Area" were recoded to “Urban.” All other responses were recoded to “Rural,” with the exception of “Postcode not linkable.” Those with values of “Postcode not linkable” were set as NA after imputation.

All nested variables were excluded from analysis, except for several variables where we could recode to account for nesting. Two variables were recoded in this way before imputation and then imputed normally. For smoking pack years (field ID 20161) and smoking pack years as a proportion of life span exposed to smoking (field ID 20162), missing values were recoded as 0 if the respondent was coded as “No” in response to the derived ever smoked variable available in the UK Biobank (field ID 20160), but otherwise left as NA if respondents had ever smoked.

We also recoded several nested variables after multiple imputation. For bread type, cereal type, and coffee type variables, all participants with a response of 0 for the bread, cereal, and coffee intake questions were coded to “Never eat bread”, “Never eat cereal,” and “Never drink coffee,” respectively. After imputation, we also recoded the number of people living in your household variable to NA if a respondent reported living in a care home or sheltered accommodation, since that question was not asked to these participants and there were very few of them. We then recoded dummy variables created from field ID 6141 (how are you related to people in your household). Specifically, if the participant reported being the only person living in their household, then each dummy variable was set to “No.” Additionally, if the participant reported living in a care home or sheltered accommodation, then each dummy variable was set to NA. For example, for the yes/no dummy variable indicating whether the participant lives at home with parents, the response was recoded to “No” if that same participant reported living as the only person in their household and was set to NA if that same participant reported living in a care home or sheltered accommodation.

*Non-exposome response level recoding.* For two nominal categorical variables, we removed a specific level as we deemed that it was capturing disability and not the exposome. The first was Type of accommodation lived in (field ID 670), where we recoded anyone reported living in “sheltered accommodation” to NA and removed that response level from the variable. The second was Major dietary changes in the last 5 years (field ID 1538), where we recoded anyone reported “Yes, because of illness” to NA and removed that response level from the variable.

*Diet variable recoding*. Following previous research on diet in the UK Biobank [8], we recoded a number of diet variables. For processed meat (field ID 1349), poultry (field ID 1359), oily fish (field ID 1329), and non-oily fish (field ID 1339), we combined the top three frequencies to get four categories: never, < 1.0 time per week, 1.0 time per week, and ≥ 2.0 times per week. For cheese intake (field ID 1408), we combined the bottom two frequencies and the top two frequencies to get four categories: < 1.0 time per week, 1.0 time per week, 2.0-4.9 times per week, and ≥ 5.0 times per week. For daily tea intake (field ID 1488), we grouped participants into the following categories: < 2.0 cups per day, 2.0-3.9 cups per day, 4.0-5.9 cups per day, and ≥ 6.0 cups per day. For daily coffee intake (field ID 1498), we grouped participants into the following categories: 0 cups/day, 0.5-1.9 cups per day, 2.0-2.9 cups per day, and ≥ 3.0 cups per day. Cereal (field ID 1458), bread (field ID 1448), and water (field ID 1528) intake were categorized into quartiles based on participants’ responses. Cereal was coded as < 2 bowls/week, 2-4.9 bowls/week, 5-6.9 bowls/week, ≥ 7 bowls/week. Bread was coded as <8 slices/week, 8-13.9 slices/week, 14-19.9 slices/week, ≥ 20 slices/week. Water was coded as <1 glass/day, 1-1.9 glass/day, 2-2.9 glass/day, ≥ 3 glass/day.

Finally, in line with previous large-scale research on alcohol consumption [9], we restricted analyses of alcohol intake frequency (field ID 1558) to current drinkers only. Previous research has shown that including previous drinkers and never drinkers together in measures of current alcohol intake frequency leads to substantial confounding in mortality analyses due to the fact that many previous drinkers are those that stopped drinking due to some health issue or disability [10,11]. For alcohol intake, all participants who responded as “Never” or “Previous” drinkers to the alcohol status variable (field ID 20117) were coded as NA. In addition, we coded participants who responded as drinking on “Special occasions only” as NA. The final variable was coded as a nominal variable with responses for “One to three times a month”, “Once or twice a week”, “Three or four times a week”, and “Daily or almost daily”, with “One to three times a month” set as the reference.

### 1.5 Derived variables

Several derived variables were calculated in our UK Biobank dataset. All derived variables were calculated after imputation, and the original variables used to create each of the derived variables were then excluded from the XWAS analysis (with the exception of diet variables used to construct the partial fiber score).

*Ethnicity*. Responses to the baseline UK Biobank self-reported ethnicity question (field ID 21000) were condensed into Black, Asian, white, mixed, and other response categories. This condensation largely follows previous research in the UK Biobank [12], although further collapses all reported Asian ethnic backgrounds into a single response category. Response categories were re-coded as follows: Black = "Black or Black British", "Caribbean”, "African", "Any other Black background"; White = "White", "British", "Irish”, "Any other white background"; Mixed = "Mixed”, "White and Black Caribbean", "White and Black African”, "White and Asian", "Any other mixed background"; Asian="Asian or Asian British", "Chinese", "Indian", "Pakistani, "Bangladeshi", "Any other Asian background"; Other = "Other ethnic group”.

*Sleep*. We created a categorical variable for hours of sleep using the UK Biobank hours of sleep continuous measure (field ID 1160). In line with recommendations from the American Academy of Sleep Medicine and Sleep Research Society [13], category levels used for hours of sleep were <7 hours, 7-9 hours, and >9 hours, with the reference set as 7-9 hours.

*Education*. Following previous research on education in the UK Biobank [14-16], an education years variable was created by converting the responses from the education qualifications variable (field ID 6138) into the equivalent years of education. Response categories were mapped onto years of education using the International Standard Classification of Education (ISCED) scale as follows: 7 years = "none of the above (no qualifications)"; 10 years = "CSEs or equivalent" or "O levels/GCSEs or equivalent"; 13 years = "A levels/AS levels or equivalent"; 15 years = "other professional qualification"; 19 years = "NVQ or HNC or equivalent"; 20 years = "college or university degree." Where participants marked multiple educational qualifications, the qualification with the highest corresponding years of education was used for that participant.

*Standardized lung function.* Standardized FEV1 and FVC variables were created by dividing the FEV1 (field ID 20150) and FVC (field ID 20151) best measure variables by standing height squared (field ID 50). This was done to ensure that values for these variables were not strongly determined by body size [17,18].

*Normalized hand grip strength.* Hand grip strength variables (field IDs 46-47) were normalized to body mass by dividing by weight [19,20].

*Bread and cereal fiber scores.* Following a method previously reported in the UK Biobank [21], we created bread and cereal fiber scores using baseline self-report data on intake of bread type and bread intake, and breakfast cereal type and breakfast cereal intake. Bread and cereal intake were measured as a numeric response corresponding to portions consumed per week. Bread and cereal intake were divided by 7 to get an estimate of daily intake, and this daily intake was multiplied by the estimated fiber content for the specific type of bread and cereal that each participant reported to mainly eat (Table S10). Participants who indicated "less than one" in response to the weekly bread and cereal intake questions were recoded as 0.5.

We converted our continuous fiber scores into quintiles and analyzed bread and cereal fiber as an ordinal variables. After creating the bread and cereal fiber scores, consumption of the individual food components of the score were not retained for further analyses.

*Total red meat consumption*. We created a new total red meat consumption variable by summing the frequencies for beef (field ID 1369), pork (field ID 1389), and lamb/mutton (field ID 1379), using the following coding: ‘Never’ = 0, ‘Less than once a week’ = 0.5, ‘Once a week’ = 1, ‘2-4 times a week’ = 3, ‘5-6 times a week’ = 5.5, ‘Once or more daily’ = 7. These values were then summed across the three variables and participants were coded into 4 categories of total red meat consumption: <1 time per week, 1.0-1.9 times per week, 2.0-2.9 times per week, and ≥3.0 times per week.

*Total fruit consumption*. Participants were asked to enter the number of pieces of fresh fruit (field ID 1309) and dried fruit (field ID 1319) they eat per day. One piece of fresh fruit, and two ‘pieces’ of dried fruit were counted as a serving. We then summed the number of total servings consumed per day and grouped participants into the following categories: < 2.0 servings per day, 2.0-2.9 servings per day, 3.0-3.9 servings per day, and ≥ 4.0 servings per day.

*Total vegetable consumption*. Participants were asked to enter the number of heaped tablespoons of cooked vegetables (field ID 1289) and salad/raw vegetables (field ID 1299) they eat per day. Two heaped tablespoons of either type of vegetables were counted as a serving. We then summed the number of total servings consumed per day and grouped participants into the following categories: < 2.0 servings per day, 2.0-2.9 servings per day, 3.0-3.9 servings per day, and ≥ 4.0 servings per day.

*Total dairy milk intake*. We derived an estimate of total dairy milk intake using the questions on type of milk (field ID 1418), bowls of breakfast cereal (field ID 1458), cups of tea (field ID 1488), and cups of coffee (field ID 1498). For participants who selected one of ‘Full cream’, ‘Semi-skimmed’ or ‘Skimmed’ milk intake, we calculated their total daily dairy milk consumption by summing 100 mL of milk for each bowl of breakfast cereal, 35 mL of milk for each cup of tea, and 25 mL of milk for each cup of coffee. Participants were then divided into three categories: those that consumed < 150 mL of milk, 150-299 mL of milk, and ≥ 300 mL of milk daily. Participants who answered ‘Never/rarely have milk’ to the question on type of milk consumed were assigned to the first category. This derivation of totally daily dairy milk intake was previously shown in the UK Biobank to discriminate well between those who had low and high dairy milk intakes according to the more detailed 24-hour dietary assessment variables [8].

*Leisure time physical activity.* As has been done previously [22], we used responses from a mark all that apply question (field ID 6164) asking participants about types of physical activity they have undertaken in the past 4 weeks to create a summary leisure time physical activity (LTPA) score. For each type of activity, if the participant reported undertaking that activity in the past 4 weeks then they were asked how many times in the past 4 weeks and the how long they spent on the activity each time. Those responses were used to determine the number of days/week for each activity and the number of minutes for each activity. These values were multiplied and divided by 7 to get the mins/day for each activity, which was multiplied by the metabolic equivalent of task (MET) for each activity (Table S11). We then summed these values across all variables to get a total LTPA MET value for each participant. Participants were categorized into three groups of LTPA using the IPAQ scoring system (https://www.physio-pedia.com/images/c/c7/Quidelines_for_interpreting_the_IPAQ.pdf): (1) high activity: 3,000 MET-mins per week or greater; (2) moderate: 600-3,000 MET-mins per week; and (3) low: less than 600 MET-mins per week.

*Occupational physical activity.* As has been done previously [22], we used responses on whether participants’ work involves heavy manual or physical work (field ID 816) or involves mainly walking or standing (field ID 806) to create a summary occupational physical activity (OPA) score. For both variables, participants reported how often they performed each type of work by choosing from the following categories: “Never/rarely”, “Sometimes,” “Usually,” or “Always.” To estimate minutes spent per week in each of these types of work, we first multiplied the hours of employment per week (field ID 767) by 60 to get the total minutes per week that each participant spent at work. Values of mins of employment per week were set to 0 for all participants who did not indicate that they were in paid employment or self-employed (field ID 6142). We then calculated the number of minutes spent specifically in each type of heavy manual and walking/standing work by adjusting the total minutes of work per week according to participants responses to how frequently they engage in heavy manual or walking/standing work. Specifically, for both variables we multiplied the minutes spent working per week by: 0 if the participant replied “Never/rarely”; 1/3 if the participant replied “Sometimes”; 2/3 if the participant replied “Usually”; and 1 if the participant replied “Always.” In this way, we calculated the approximate minutes per week that each participant spent in both heavy manual and walking/standing work. The mins per week for both types of work were then multiplied by the metabolic equivalent of task (MET) for each activity (Table S11) and these values were summed across both variables to get a total OPA MET value for each participant. We used the same MET thresholds per week as the LTPA variable to categorize participants into low, moderate, and high OPA.

*Total sedentary time.* Total sedentary time was measured according to a previously reported method in the UK Biobank [23], using measures on self-reported hours spent on a typical day watching television (field ID 1070), using the computer (field ID 1080), and driving (field ID 1090). Values for each variable greater than 24 hours per day were excluded, and those reporting over 16 hours were re-coded to 16 hours. Tertiles were used to categorize sedentary time into low (0-4 hours), medium (5-6 hours), and high (>6 hours) levels of sedentary behavior, with 0-4 hours set as the reference and the variable was classed as an unordered factor.

### 1.6 Variable quality control

As a final quality control (QC) step conducted after imputation but before running the sex-specific XWAS, we systematically examined the crosstabs between each potential categorical exposure variable and the mortality binary indicator in both women and men. Any categorical variable with less than 10 mortality cases for a single response level was then flagged for further inspection. Three possible quality control actions were then undertaken:

(1) Binary categorical variables with less than 10 mortality cases for one of the response levels were completely excluded from the XWAS.

(2) We collapsed the “All of the time” and “Often” response levels for the narcolepsy variable, and we also collapsed the “Yes” and “I am completely deaf” response levels for the variable “Do you have any difficulty with your hearing?”

(3) For nominal categorical variables, any response levels with too few mortality cases (n=5) were set to NA and therefore not analyzed in the XWAS. The only variable that this affected was the “What type of accommodation do you live in?” variable, where the response level indicating that participants were living in a care home was set to NA and not analyzed in the XWAS because too few participants endorsed that response.

In order to improve the interpretability of our results, all responses of “none of the above” or “other” for nominal variables were set to NA and not analyzed. The response level of “None of the above” was also set to NA for the usual walking pace variable, and the resulting response levels of "Brisk pace", "Steady average pace", and "Slow pace" were recoded to be an ordinal variable with slow pace as the highest response level. Usual walking pace was only used as an exposure in phenome-wide association study analyses and not in the XWAS.

#### 1.7 Multiple imputation

The average percentages of missing data across all final variables included in our UK Biobank analysis datasets were 11% in women (range: 0-79%) and 10.9% in men (range: 0-77%). As shown in Fig. S3, the rates of missing data for the subset of 41 exposures identified as independently associated with premature mortality was lower, with the majority of variables missing < 5% of responses and all but one variable missing < 20% of responses. Because of the large heterogeneity in number of missing responses across exposure variables included in our analyses, we created multiple imputed datasets instead of producing a single imputed dataset. Creating a single imputed dataset does not account for the uncertainty caused by the missing data and tends to inflate the significance of p-values, artificially narrow confidence intervals, and represent variables as more strongly associated than they really are [24]. Studies using simulation data alongside ALSPAC cohort data have shown that multiple imputation, even when used on datasets with large proportions of missing data (up to 90%), can still produce unbiased regression results so long as the imputation model is properly specified and the data are missing at random [25]. Indeed, it has been shown that multiple imputation is often more efficient than complete case analyses for estimating the coefficient of a regression variable when other model covariates are incomplete (i.e., have missing data) [26,27].

Missing data were imputed separately for the discovery, replication, and validation datasets (Fig. 1). We used the R package missRanger [28] to perform multiple imputation, which combines random forest imputation with predictive mean matching. Together, these methods offer a robust approach to imputed missing data in heterogeneous datasets (i.e., datasets containing both continuous and categorical data). Random forest imputation is a non-parametric method that is particularly appropriate when a normal distribution for all variables cannot be assumed, as it will preserve any skewness in the original data, and has also been shown to have the least imputation error and greatest accuracy when compared with other leading methods such as multiple imputation by chained equations (MICE) [29,30]. Furthermore, random forest imputation is able to handle collinearity between variables well during imputation, whereas multiple imputation carried out from predictive mean matching alone does not perform well in the presence of collinearity. In fact, early attempts at performing multiple imputation via predictive mean matching using the *mice* [31] package in R with our dataset would not even run successfully because of the high collinearity present between various index of multiple deprivation variables, as well as among air pollution variables.

We imputed 5 datasets, with a maximum of 10 iterations specified for each imputation. Each imputation was also weighted by degree of missing data for each participant, such that the contribution of data from participants with higher proportions of missingness were weighted down in the imputation. We set the maximum number of trees for the random forest to 200, but left all other random forest hyperparameters at their default. The imputation datasets included all exposures, in addition to all baseline disease and aging phenotype variables to be used in later analyses. The imputation procedure considered all variables as predictors to impute missing values in all other variables. In addition, the Nelson-Aalen estimate of cumulative hazard and the all-cause mortality event indicator were also used to predict imputation values, as it has been shown that imputation incorporating the Nelson-Aalen estimate of cumulative hazard to the survival time $H_0(T)$ and the event indicator as predictors improves the imputations for survival analyses [27]. The average out-of-bag (OOB) error rate for multiple imputation across all imputed datasets was 0.077 in women (range: 0 – 0.75) and 0.076 in men (range: 0 – 0.68).

All subsequent study analyses were run independently in each of the five imputed datasets, and results were pooled using Rubin's rule [32].

### 1.8 Exposome-wide association study (XWAS) modeling

XWAS of all-cause mortality were initially carried out separately in women and men, and then a final XWAS was calculated in the pooled dataset with both women and men to increase power. For both the discovery and replication XWAS, we serially assessed associations of each individual exposure variable with all-cause mortality using Cox proportional hazards models. A discussion and rationale for each of the covariates in the final XWAS model are given below.

Mortality risk varies significantly with age. To account for this, Cox models in both the discovery and replication XWAS were calculated using age-at-risk as the time scale for the survival analysis instead of follow-up time in the study (i.e., number of years between date of recruitment into the study and date of censoring). It has been previously shown that using follow-up time in study as the time scale, while merely adding age as a covariate, does not adequately reduce age-related bias in survival analyses for outcomes that are age-related [33,34]. Using age-at-risk as the time scale has been shown to adequately mitigate this bias in survival analysis because it puts similar subjects in a risk set together and allows for a non-parametric age effect. Using age-at-risk as the time scale in a survival model is performed using three variables to calculate the survival outcome: age at recruitment, age at censoring, and the event indicator. The difference between these two approaches is demonstrated in the two R formulas below:

$$coxph(Surv(f,d) \sim e + x, data) \quad (1)$$

$$coxph(Surv(e,a,d) \sim x, data) \quad (2)$$

where formula 1 is a more common survival formula using follow up time in study (*f*) and the event indicator (*d*) to calculate the survival outcome, with age at recruitment (*e*) entered as a covariate (ideally centered to the mean) alongside other exposures and covariates of interest (*x*). Formula 2 is an age-at-risk survival formula using age at recruitment (*e*), age at censoring (*a*), and the event indicator (*d*) to calculate the survival outcome. Note that age at recruitment and age at censoring are not centered or standardized in the age-at-risk analysis, and a separate covariate for age is not needed in the model.

Geographic location has been shown to significantly influence mortality and life expectancy in the UK, even after accounting for differences in deprivation between areas [35]. Accordingly, UK Biobank assessment center (as a proxy for geographic location) was added to our XWAS model as a covariate. In a meta-analysis of 48 prospective cohort studies (n=1,700,000), socioeconomic status was shown to associate significantly with premature mortality, contributing the third highest population attributable fraction for mortality compared with conventional risk factors (behind smoking and physical activity) [36]. We therefore considered education years as potential covariates, as measures of socioeconomic status. Ethnicity was also considered as a covariate, as this has also been used in multivariable adjustment from a previous XWAS of all-cause mortality [37].

Each XWAS model was also stratified by two variables: 5-year birth cohort and sex. Adding strata for 5-year birth cohorts adjusts for cohort effects, as cohort effect patterns have been demonstrated for many age-related diseases and in mortality patterns across multiple European countries (including England) for both sexes and almost all causes of death [38,39]. Participants were assigned to one of seven birth cohort categories (in 5-year intervals from 1935 until 1970) using their year of birth. XWAS models were also stratified by sex since mortality rates were much lower in women compared to men in our dataset (5% vs. 8.7%), and thus we could not assume a common baseline mortality hazard between men and women. When conducting preliminary model selection analyses in our UKB dataset, a model using a strata term for sex resulted in a much better model fit according to AIC compared with an identical model using a fixed effect covariate for sex instead of a strata term.

The Cox proportional hazards model used in the final XWAS was composed with age-at-risk as the time scale; included covariates for UK Biobank assessment center, years of education (7 years, 10 years, 13 years, 15 years, 19 years, 20 years), and ethnicity (White, Asian, Black, Mixed, Other).; and was stratified by 5-year birth cohorts and sex. XWAS Cox models were analyzed with the survival [40] package in R using the following formula:

$$coxph(Surv(e,a,d) \sim strata(b,s) + t + g + r + x, data) \tag{3}$$

where *e* is age at recruitment, *a* is age at censoring, *d* is the event indicator (mortality = 1, no mortality or censored = 0), *b* is 5-year birth cohort, *s* is sex (female/male), *t* is UKB assessment center, *g* is years of education, *r* is ethnicity, and *x* is the individual exposure being tested. Adding a stratification variable in a Cox proportional hazards model allows for separate baseline hazard functions to be fitted within each of the strata, while keeping other covariates constant. In our model, baseline hazards were thus calculated separately within each combination of sex and 5-year birth cohorts (14 total strata), and the resultant regression coefficients returned in the model results are optimized across all strata. Early sex-specific XWAS runs used an identical model, but omitted the strata term for sex.

Since it has been shown that UK Biobank participants are likely to misreport alcohol consumption as a function of higher disease burden [41], self-reported overall health status was added as an additional XWAS covariate for self-reported alcohol intake. Indeed, when testing the association between self-reported alcohol intake mortality without adjustment for self-reported overall health status, we observed that all levels of alcohol intake showed decreased risk of mortality compared with the lowest level of intake, which is counter to well established results from previous large meta-analyses [9,11] and a Mendelian randomization study in the UK Biobank [42].

Exposures in the final pooled XWAS were limited to those asked to both women and men, omitting sex-specific reproductive factors. Sex-specific XWAS variables showing significant associations with all-cause mortality included having had a cervical smear test, having gone through menopause, and number of children in women; and puberty timing (relative age of voice breaking and gaining facial hair) in men. These results are shown in Tables SF3-SF4. These variables were not tested in further disease sensitivity or correlation-based multivariable analysis, however replicated sex-specific factors from the sex-specific XWAS analyses were added to analyses of sex-specific disease outcomes (breast, ovarian, and prostate cancer).

The covariates for education and ethnicity were assumed to be related to mortality based on previous literature and were not tested independently as exposures in the XWAS. Replication XWAS analyses only tested those variables significant in the discovery. P-values in the discovery and replication analyses were corrected using the false discovery rate (FDR; Benjamini-Hochberg method [43]) with a significance threshold of FDR $p < 0.05$.

### 1.9 Prevalent disease sensitivity

A sensitivity analysis was conducted to test whether exposures replicated in the full cohort XWAS were susceptible to reverse causation due to prevalent disease at baseline. We conducted two analyses. The first was conducted in the full sample of participants recruited in England (n=436,891) where we individually tested every exposure replicated in the pooled mortality XWAS again in relation to mortality using the pooled XWAS formula and covariates, but now adding an interaction term between each exposure and an indicator of baseline disease or poor health (see definition below):

$$coxph(Surv(e,a,d) \sim strata(b,s) + t + g + p + r + x*i, data) \qquad (4)$$

where *e* is age at recruitment, *a* is age at censoring, *d* is the event indicator (mortality = 1, no mortality or censored = 0), *b* is 5-year birth cohort, *s* is sex (female/male), *t* is UK Biobank assessment center, *g* is years of education, *r* is ethnicity, *x* is the individual exposure being tested, and *i* is the baseline poor health indicator. The interaction terms used returned estimates for the direct effect of the exposure *x* and poor health indicator *i*, as well as the interaction *x*i*. We flagged any exposure that no longer had a significant direct effect in this model ($p < 0.05$) but its interaction with the baseline poor health indicator was significant ($p < 0.05$). This retrieved 12 variables whose associations with premature mortality are likely completely explained by poor health status at baseline in this dataset (labelled in Fig. S1), and these variables were removed from any further analysis.

The baseline disease/poor health indicator was created for all participants, in which participants were coded as having disease or poor health at baseline if they: (1) had a linked hospital inpatient ICD diagnosis for any of the chronic illnesses or cardiometabolic risk factors studied in our analysis (including hypertension, dyslipidemia, and obesity) with a diagnosis date before or on their date of recruitment to the UK Biobank; (2) were assigned a diagnosis code for any of the chronic diseases or cardiometabolic risk factors studied in our analysis during the baseline clinical interview (field IDs 20001, 20002; Table S9); (3) self-reported a physician diagnosis of heart attack (field ID 6150), angina (field ID 6150), stroke (field ID 6150), high blood pressure (field ID 6150), bronchitis/emphysema (field ID 6152), diabetes (field ID 2443), or cancer (field ID 2453); (4) self-reported a number of cancer diagnoses ≥ 1 (field ID 134); (5) self-reported taking insulin medication (field ID 6153, 6177), cholesterol lowering medication (field ID 6153, 6177), or blood pressure medication (field ID 6153, 6177); or (6) self-reported their overall health status as “poor” (field ID 2178).

To further confirm the robustness of our pooled mortality XWAS results to reverse causation, we also performed the pooled XWAS a second time in the subset of participants with no disease at baseline (testing only those exposures that were replicated in the pooled XWAS in the full sample). Betas for each exposure from each of these two XWAS were then plotted against each and the correlation between them was calculated. As shown in Fig. S2, the correlation between the mortality betas in these two populations is extremely strong (Pearson’s R = 0.96), indicating that the effect estimates from the pooled XWAS in the full sample are largely not biased due to reverse causation.

#### 1.10 PheWAS of replicated exposures

For all exposures replicated in the XWAS and not removed during the above-described disease sensitivity analyses, a phenome-wide association study (PheWAS) was conducted. In each PheWAS, the exposure was used as the outcome variable (hereafter referred to in this section as the exposure-outcome) and was tested against the full set of baseline phenotypes available in the UK Biobank (see Supplementary File SF61 for full list of phenotypes tested). Each PheWAS was conducted as a linear or logistic regression, depending on whether the exposure-outcome was continuous or categorical, with covariates for age at recruitment and sex. All ordinal exposures exposure-outcomes were tested as continuous variables. Nominal categorical exposures were recoded into dummy variables for each response category vs. the reference. All continuous phenotype exposures were scaled and centered to the mean before running the PheWAS. Volcano plots were created from PheWAS results and were inspected visually for phenotypes with strong effect estimates for their association with the exposure-outcome. Example volcano plots are shown in Fig. S4-S5. Interactive volcano plots from all PheWAS are shown in the Online Materials. Summary statistics from all PheWAS are available in Supplementary Files SF62-SF177. Using this method, a further 10 variables were excluded that likely suffer from mismeasurement error and either capture disease/disability status or seem to be capturing redundant information to another exposure: living at home with parents, living at home with siblings, using a mobile phone, length of mobile phone use, difference in mobile phone use now vs. 2 years ago, frequency of driving fast on the motorway, number of vehicles in household, facial aging, number of sexual partners, and age when first had sexual intercourse.

### 1.11 Correlation and cluster analyses

Correlation between all variables was calculated among all UK Biobank participants recruited in England (n=436,891) using the R package polycor [44] to create a heterogenous correlation matrix for each imputed dataset, consisting of Pearson correlations between continuous variables, polyserial correlations between continuous and ordinal variables, and polychoric correlations between ordinal variables. All nominal variables were recoded to k-1 dummy variables before correlation analyses were conducted. Correlation coefficients were first calculated within each imputed dataset, then transformed to a normally distributed z-score via Fisher's z transformation, pooled via Rubin's rule, and then re-transformed back to the original r-scale coefficient after pooling. This was done to create a normally distributed sampling distribution for the correlation coefficients, which is required for pooling via Rubin's rule. P-values for the significance of each pooled correlation coefficient were calculated using the pooled z-transformed correlation coefficient and the standard error based on sample size.

Our exposome dataset exhibited a densely intercorrelated structure among variables replicated in the mortality XWAS. As has been observed previously in environment data [45,46], despite a very low percentage of observed inter-variable correlations with a coefficient of correlation above 0.50 or below -0.50 (4.08%), a very large percentage of inter-variable correlations were significant according to p-values for the correlations. Specifically, 90% of inter-variable correlations between exposures replicated in the mortality XWAS exhibited a significant Bonferroni-corrected correlation p-value below 0.001 (mean absolute value of significant correlation coefficients: 0.09). Only 7.01% of correlation coefficients had an absolute value of 0.30 or above.

Exposures replicated in the mortality XWAS and not confounded by baseline disease status were considered for clustering analysis, using the correlation coefficients between these variables. We used both k-means and hierarchical clustering to determine an adequate cluster structure for the data. To get an initial approximation of the number of clusters that fit the data, we used both AIC and the total within-cluster sum of squares (WSS).

For AIC analyses, we first computed the k-means clustering of the exposures for different numbers of clusters (k) ranging from 1-100. k-means clustering is a machine learning algorithm that determines a pre-specified number of centroids (means) in the correlation coefficient values and then partitions the variables in the dataset into unique clusters based on the distance of their correlation coefficients from the centroids. For each k, we then calculate the AIC of the cluster model as the WSS + 2(k*number of total variables) [47]. We then plotted the AIC as a function of the number of clusters k and examined the plot visually to try to find the elbow in the plot (Fig. S6). An elbow in the plot could not be determined visually, and was thus calculated as the maximum absolute second derivative of the curve of AIC values for k, resulting in 9 clusters.

For WSS analyses, we first computed the hierarchical clustering of exposures for different numbers of clusters (k) ranging from 1-100. For each k, we then calculated the WSS, which is a measure of the variability of the observations within each cluster. A cluster with a small WSS is more compact and has lower variability of observations within a cluster than a cluster that has a large WSS. We plotted the WSS as a function of the number of clusters k, and examined the plot visually to find the elbow in the plot (Fig. S7). This point represents the number of clusters k where adding another cluster doesn't greatly improve (i.e., lower) the total WSS. The WSS plot was created using the fviz_nbclust function in the package factoextra [48].

AIC analysis using k-means clustering initially returned 11 clusters as the optimal solution, however when plotting the k-means clustering according to 11 clusters, this cluster solution did not adequately break up clusters into discrete groupings. We therefore iteratively plotted lower numbers of k-means clusters starting from 10 until the variables were grouped discretely. This brought us to a total of 6 k-means clusters (Fig. S8). We determined however that a 9-cluster solution was a better approximation of the elbow in the WSS curve, preferentially using the WSS plot information since our final cluster structure would be based on hierarchical clustering. When visually inspecting the dendrogram of hierarchical correlation, 9 clusters separate the variables very well in terms of breaking variables into discrete groups with large distances/heights between clusters (Fig. 3c).

### 1.12 Cluster multivariable modeling procedures

As single multivariable model containing all exposures replicated in the XWAS across all clusters was not carried out as the final step in identifying relevant exposures to mortality because certain domains represented in different clusters are mediators of others in the pathway towards mortality. Therefore, adopting this approach would lead to many true determinants of mortality adjusting out of the model due to these other mediating variables in the model. Our own initial analysis using this approach showed this to be the case, and it has also been documented in the literature, where a previous XWAS of all-cause mortality in a small sample from the NHANES cohort put all exposure significant in their XWAS in a single model in which smoking, one of the strongest known factors affecting disease and mortality, adjusted out of the model [37].

We therefore carried out multivariable mortality models within each of the clusters described above as a final step to identify correlation confounding in exposures. As described above, our clustering analysis retrieved 9 independent clusters of exposures replicated in the mortality XWAS. We further conducted multivariable modeling within each of these 9 clusters using the following procedure: (a) All exposures in the cluster were run in a single multivariable mortality Cox model to check for multicollinearity using the variance inflation factor (VIF). VIF analysis was carried out using the car package in R [49]. Exposures with a GVIF^(1/(2*Df)) > 1.6 were flagged for collinearity and removed. GVIF^(1/(2*Df)) is similar to GVIF, but takes into account degrees of freedom for categorical variables. (b) After any collinear variables are removed, all remaining exposures in the cluster were tested together in a single multivariable mortality Cox model using age as the timescale, stratified by 5-year birth cohorts and sex, and adjusted for UK Biobank assessment center, household income (Less than 18,000, 18,000 to 30,999, 31,000 to 51,999, 52,000 to 100,000, Greater than 100,000), education, and ethnicity (if those variables were not already in the cluster). As in the XWAS, additional adjustment for self-reported overall health status was made in the cluster with self-reported alcohol intake. This process led to a number of variables adjusting out that were confounded by short-range correlation between other similar exposures (Fig. S11a).

To account for confounding due to longer-range confounding between exposures, we also combined variables from neighboring clusters. We then took all variables significant in clusters 1-5 and put them in the same multivariable mortality Cox model. Separately, we took all variables significant in clusters 6-9 and put them in the same multivariable mortality Cox model. Variables from clusters 1-5 and 6-9 were not put together into a single model for the reasons described above. These models used the same covariates as the previous stage of cluster modeling. At the end of this process, we were left with 41 exposures that are less likely to be susceptible to confounding from other correlated exposures. Significance in all the cluster multivariable models was determined by a nominal $p < 0.05$.

Air pollution, greenspace, noise pollution, and traffic variables contained multiple, redundant variables that were too correlated and collinear to be used in conjunction with one another in the same model (correlation coefficient > 0.97). This would have required arbitrarily selecting a single air pollution variable to test while discarding the rest, which was undesirable. Instead, we ran separate principal component analyses (PCA) among the nine air pollution variables (NO [2010], NO2 [2005, 2006 ,2007, 2010], PM2.5 [2010], PM2.5 absorbance [2010], and PM10 [2007, 2010]), four greenspace variables (greenspace buffer 1000m, greenspace buffer 300m, natural environment buffer 1000m, natural environment buffer 300m), four noise pollution variables (average daytime noise pollution, average evening noise pollution, average night noise pollution, average 16 hour noise pollution, and average 24 hour noise pollution), and two traffic variables (sum of road length of major roads within 100m, total traffic load on major roads) that were all significant after the pooled XWAS and disease sensitivity analysis. Three

PCs were retrieved that captured >90% of the variation in the air pollution variables, two PCs captured >96% of the variation in greenspace variables, one PC captured >99% of the variation in noise pollution variables, and one PC captured > 91% of the variation in traffic variables. Plots of principal component loadings are shown in Fig. S9 (participants recruited in England) and Fig. S10 (participants recruited in Scottland/Wales). These PCs were used in all cluster models described above instead of the individual variables.

#### 1.13 Aging mechanism and incident disease analysis

All exposures replicated in the mortality XWAS that survived the disease sensitivity and cluster modeling states were tested in relation to 25 age-related blood biomarkers (Table S7-S8). Aging biomarker analyses were cross-sectional, with analyses of leukocyte telomere length (LTL) in a smaller sample since it was only released for a subset of participants (n=410,877). Serum biomarker variables were log transformed and then age-adjusted by regressing each onto recruitment age separately in women and men and then adding the residuals and the intercept to make an age-adjusted variable. All aging mechanisms were tested in linear regression models with covariates for sex, 5-year birth cohort, assessment center, years of education, ethnicity, number of medications, smoking status, and IPAQ physical activity level. IGF-1, LTL, and vitamin D models included additional covariates for standing height, leukocyte count, and month of biomarker assessment (to control for seasonality of vitamin D sun exposure), respectively.

All exposures replicated in the mortality XWAS that survived the disease sensitivity and cluster modeling states were also tested in relation to 8-15 year incidence of 25 chronic disease categories (Table S9), as well as three clinical risk factors (obesity, hypertension, dyslipidemia).

This included all diseases in the top 20 causes of death in the UK Biobank as of July 28, 2022 (except for causes related to accidents/external injury and infectious disease) as well as other conditions that are highly prevalent in aging populations. We serially assessed each exposure and disease pair using Cox models that used age-at-risk as the timescale, were stratified by 5-year birth cohort and sex, and were adjusted for the following covariates: UK Biobank assessment center, years of education, ethnicity, smoking status, and IPAQ physical activity level. Each set of biomarker and disease analyses was corrected separately for multiple testing using FDR. Heatmaps and annotation tracks for biomarker and incident disease analyses shown in Fig. 7 were created using the ComplexHeatmap package in R [50].

For cardiometabolic risk factors (obesity, hypertension, dyslipidemia), we serially assessed each exposure and risk factor pair using cross-sectional logistic regression models adjusted for age, sex, assessment center, household income, years of education, ethnicity, smoking status, and IPAQ physical activity level.

### 1.14 Final multivariable exposome modeling

For each outcome, five multivariable models were calculated. The first only includes age (scaled) and sex in the model (model 1). Model 2 includes age, sex, and the polygenic risk score (PRS) for the outcome, if available (see below for more detail). Model 3 includes age, sex, and all exposures associated with the outcome (exposome). Model 4 includes age, sex, exposome, and PRS. If a PRS was not available for a particular outcome, then models 2 and 4 were not calculated for that outcome. Each model was validated in the independent dataset of participants recruited in Scotland/Wales (n=55,676) by obtaining the linear predicted values from the models in the participants recruited in England and measuring the C-index and $R^2$ for these values in relation to the outcome rates in the validation set. Effect estimates from model 3 for all chronic diseases studied are shown in Figs. S35-58.

For sex-specific outcomes (breast, ovarian, and prostate cancers), we also included in the exposome all sex-specific exposures that were replicated in the female- and male-only mortality XWAS. Across all models for all outcomes, we excluded UK Biobank assessment center, as its responses are location-specific and do not generalize to other populations. Ordinal categorical variables were allowed all polynomial contrasts for these final multivariable models. The Cox proportional hazards models used for these multivariable models different slightly from those used in previous analyses, instead using time in study as the timescale, using recruitment age and sex as fixed covariates, and removing birth cohort from the model given its collinearity with age. This is represented in the below R formula for model 4:

$$coxph(Surv(f,d) \sim e + s + g + x, data) \tag{5}$$

where *f* is survival time for the outcome, *d* is the event indicator, *e* is age at recruitment (scaled), *s* is sex, *g* is the polygenic risk score and associated genetic covariates for the outcome, and *x* is a vector of all exposures associated with the outcome in previous analysis steps.

Because different variables in our dataset had small amounts of NA values after our recoding procedures (see above), adding more and more variables to a single model increases the number of participants that are dropped from regression due to an NA response across any variable. To ensure that model performance would not be affected by this and to avoid overfitting, we systematically calculated crosstabs between each categorical exposure and the event indicator for each outcome. Datasets used for this crosstab procedure included only the subsets of participants with no missing values across any exposures or PRS variables for that outcome, as well as excluding any prevalent outcome cases. Any categorical exposure with less than 10 outcome cases for one of the response levels was completely excluded from all exposome models for that specific outcome. The only exception was the variable on type of accommodation lived in, where instead we recoded all responses of "Mobile or temporary structure (i.e. caravan)" to NA and removed that as a response level from the variable (since only a few hundred people endorsed this response level in the subset of participants in the multivariable models).

For each Cox model, the proportional hazards assumption was tested by examining the Schoenfeld residuals using the cox.zph function in the Survival package. For each model, if any variable that showed a p-value lower than 0.0001 in the cox.zph output, then the Schoenfeld residuals were plotted to inspect the residuals distribution. Variables were deemed to violate the proportional hazards assumption based on visual inspection of the Schoenfeld residuals to detect non-conformity of the residuals over time. For these variables, a new model was calculated with an interaction with time. Survival time splitting to use for time interactions in these models was performed using the timeSplitter function from the Greg R package [51], using two years as the interval for time splitting.

### 1.15 Calculating polygenic risk scores

Where possible, we used multi-ancestry PRS that were previously made available by the UK Biobank (Table S12). Methods for deriving these PRS are described elsewhere [52]. For cancer outcomes where no PRS were provided by the UK Biobank, we identified recent PRS from the PGS Catalog [53], selecting scores derived in predominantly European populations that did not overlap with the UK Biobank cohort (as no multi-ancestry scores were available). We calculated these PRS as weighted sums, ∑(no. risk alleles*effect size) in the UK Biobank v3 imputed genotype data. PGS catalog entries used to calculate PRS were as follows: leukemia

(PGS000077) by Graff et al. (2021) [54], lung cancer (PGS000078) by Graff et al. (2021) [54], pancreas cancer (PGS000083) by Graff et al. (2021) [54], esophageal cancer (PGS002298) by Choi et al. (2020) [55], COPD score (PGS001788) by Wang et al. (2021) [56], chronic kidney disease (PGS000859) by Mansour Aly et al. (2021) [57], non-alcoholic fatty liver disease (PGS002282) by Schnurr et al. (2022) [58], liver cirrhosis (PGS000726) by Emdin et al. (2020) [59], and knee osteoarthritis (PGS002729) by Sedaghati-Khayat et al. (2022) [60]. All variants in these scores met our quality control criteria of imputation information > 0.4 and minor allele frequency (MAF) > 0.005 in the UK Biobank data. Although these new PRS were mostly developed in European populations, we calculated the PRS for our full multi-ancestry sample and accepted the limitation that the PRS may be slightly mis-specified in non-European participants. Nonetheless, initial testing of the PRS showed that they performed well in our full sample including all ethnicities, with a strong, dose-response association across quintiles of genetic risk for all PRS except esophageal cancer, which showed weaker associations between the PRS and cancer outcome. This is consistent with the association results obtained from the original publication on the creation of this PRS [55].

All PRS were coded as quintiles for use in our multivariable models. In all multivariable models including PRS variables, we also added an additional covariate for genotype array (BiLEVE vs. Axiom; field ID 22000) as well as the first four genetic principal components published by the UK Biobank (field ID 22009).

#### 1.16 Pooling $R^2$, C-index, and Chi-squared values across imputed datasets

$R^2$ values for each model were calculated using the CoxR2 package [61] as a measure of explained randomness based on the partial likelihood ratio statistic under the Cox Proportional

Hazard model [62], which has been shown in simulation studies to perform better for survival analysis than traditional or pseudo $R^2$ metrics [63]. Following previous guidance [64], $R^2$ values were first calculated separately within each imputed dataset, converted to r-scale coefficients by taking the square root, and then converted to the z-scale using Fisher's z transformation. Z-transformed $R^2$ values were then averaged across all five imputed datasets. These averaged values were then re-transformed back to the r-scale using inverse z transformation and then squared to return a pooled $R^2$ value. C-index values were also pooled using the same method. In variable importance analyses, Wald chi-squared ($X^2$) values were obtained for each variable exposure by running ANOVA on each multivariable cox model separately in each imputed dataset using the rms package in R [65]. To obtain the pooled proportion of the total model $X^2$ that each variable and variable category explained, the sum of $X^2$ values for the same variable/category across all imputed dataset was divided by the sum of the total model $X^2$ values across all imputed datasets. Exposure importance according to total model $X^2$ for key diseases in exposome multivariable Cox models are shown in Figs. S12-S34.

#### 1.17 Systematic review of exposome-wide health studies

To better situate our research in relation to the landscape of published exposome-wide or environment-wide analyses, we conducted a systematic review to identify all exposome studies published to date on any health outcomes in humans. We searched PubMed on March 28, 2022 using the following search string: Exposom* OR Exposome-wide OR Environment-wide OR "Environment-wide association study" OR "ExWAS" OR "Ex-WAS". Eligibility criteria included peer-reviewed, original research articles containing an exposome- or environment-wide analysis of biological, health, or behavioral outcomes in humans, with no limit for year of publication. Exclusion criteria included duplicates; non-English publications; research not carried out in humans; reviews, commentaries, or methods articles; corrigendum or response articles; and preprints. Paper abstracts were screened independently for eligibility by two separate reviewers, with any discrepancies in inclusion/exclusion decisions reconciled to arrive at the final list of papers to include. Studies were reviewed for inclusion using Rayyan software [66]. Protocol for the systematic review was pre-registered on the Open Science Framework (OSF) (doi: 10.17605/OSF.IO/N3DCV; https://osf.io/n3dcv/).

1,472 studies were retrieved via our search, of which 73 were deemed appropriate to include. After retrieving all published exposome analyses to date, only one paper was identified with an "environment-wide" analysis of mortality [37]. No exposome papers were published with aging as an outcome.

**Sensitivity analyses**

*Accelerometer data and self-reported physical activity*. We also calculated mortality multivariable models using the accelerometer data available in the UK Biobank. The aim of this analysis was to compare the explanatory power of the baseline self-reported physical activity variables versus an objective measure of physical activity. Data collection for accelerometer data occurred from 2013-2015 in a subsample of 103,672 UK Biobank participants. Mortality survival times were re-calculated using the date from the start time of the accelerometer wear period (field ID 90010) as the start date of follow up for each participant. We used the overall acceleration average variable (field ID 90012) as our measure of objective physical activity. Description of the development of this variable has been described previously [67]. Before analysis, we carried out several quality control steps to refine the overall acceleration average variable by excluding participants: (1) whose data could not be calibrated (field ID 90016); (2) who did not wear the device long enough to get a stable measure of their physical activity status (field ID 90015); (3) with >1% clipped values before or after calibration (field IDs 90183, 90185) out of the total data readings (field ID 90187); or (4) who had an unrealistically high overall acceleration average of 100 milli-gravity or greater. This process excluded 6,189 participants, leaving a total sample of n=85,520 for the accelerometer analyses.

Two models were tested: the first included all self-reported baseline physical activity measures (IPAQ, LTPA, OPA, sedentary time) in the full sample of UK Biobank participants recruited in England (n=436,891), the second only included overall acceleration average in the subsample of participants with accelerometer data (n=85,520). Both models were run as Cox proportional hazards models with mortality survival time as the timescale with additional covariates for age at time of activity measurement, sex, UK Biobank assessment center, years of education, household income, ethnicity, smoking status, and Townsend deprivation index. All polynomial contrasts were allowed for all ordinal variables. When putting all self-reported baseline physical activity measures in the same model there was no collinearity according to VIF. The pooled $R^2$ from the all-cause mortality model using overall acceleration average was 0.59, whereas the pooled $R^2$ from the model using self-reported baseline physical activity measures was 0.56 (Table S13). Objective physical activity explained a greater amount of the mortality variation in our UK Biobank sample by 3%, indicating that our overall estimate of the variation of mortality explained by baseline self-reported physical activity measures underrepresents the total influence of the objectively measured physical activity on mortality by only approximately 3%.

*Prostate cancer.* After conducting analyses between all validated exposures and incident prostate cancer, we observed that many significant associations were in the opposite direction as expected. Notably, we observed all smoking variables showed an association with decreased risk of incident prostate cancer. This inverse association has been well documented in previous studies [68-70]. It has been posited that those who do not smoke may be more likely to undergo a prostate specific antigen (PSA) test and receive a diagnosis whereas those who smoke may be less likely to undergo testing and therefore would be undiagnosed or not diagnosed until a much later stage. We attempted to test this by stratifying the sample population by those who had ever received a PSA test at baseline (field ID 2365; n=137,598 for those with no PSA test, n=58,425 for those with a PSA test), as well as conducting a model in the full sample of men recruited in England (n=196,113) with PSA test as a fixed covariate. All Cox models included age as the timescale, were stratified by 5-year birth cohorts, and were adjusted for UK Biobank assessment center, household income, ethnicity, years of education, and IPAQ activity group. All polynomial contrasts were allowed for all ordinal variables.

The inverse association between smoking and incident prostate cancer remained in both those who had and had not received a PSA test (Table S14). Furthermore, adding in ever having taken a PSA test as a covariate to the model in the full sample also did not alter the inverse association observed between smoking and prostate cancer. Together, these results seem to rule out any detection bias explaining the associations between smoking and prostate cancer. Previous research has also demonstrated that increased insulin-like growth factor 1 (IGF-1) levels are associated with increased risk of prostate cancer [71] and it has been posited that smoking may reduce the risk of prostate cancer through decreasing IGF-1 [70]. Our analysis did show a significant association between smoking and decreased IGF-1 levels (Fig. 5), which may support this hypothesis.

**Supplementary discussion of study findings**

Cluster multivariable models returned robust, significant associations for many exposures that are unstudied or understudied in the context of premature mortality, including several aspects of physical activity (components of the leisure time physical activity variable, including: pleasure walks; light and heavy DIY activities like watering the lawn, digging, carpentry; swimming; and strenuous sport activity), type of accommodation lived in (apartment vs. house), and certain measures of household composition such as number of people living within your household and living with parents or siblings. Although frequency of confiding in others has previously been associated with all-cause mortality in the UK Biobank as part of a composite measure including loneliness [72], we also demonstrate that it has explanatory power for mortality on its own.

In line with previous research [9], we observed that frequency of alcohol intake showed a U-shaped association with mortality unless modelled in current drinkers only. When modeling using current drinkers, we found a J-shaped association between alcohol intake and mortality, where only the highest amount of alcohol intake was associated with increased risk of mortality compared with the lowest intake level. Previous large meta-analyses [11] and a Mendelian randomization study [42] in the UK Biobank have demonstrated increasing risk of mortality with each increase in alcohol consumption, however we could not replicate this linear dose-response association in our analyses. Of note, we also only observed an increase in mortality risk for the highest level of alcohol intake in the more parsimonious XWAS model when adding further adjustment for self-reported overall health status. This lends further support to the idea that self-reported alcohol status can be severely biased by health status at recruitment, including recent research from the UK Biobank found that participants are likely to misreport behavioral traits such as alcohol consumption, often as a function of higher disease burden [41]. It may be that the true direction of the association between self-reported alcohol intake and mortality may only be possible to detect via Mendelian Randomization. We also observed the same associations between alcohol intake and mortality in our unimputed data, and are therefore confident that our observed association is not somehow an artefact of our multiple imputation procedure.

Furthermore, our analysis replicates a previous finding from the Million Women Study showing that despite many previously reported associations between happiness and mortality, happiness has no effect on all-cause mortality when proper covariate adjustment is made [73].

Interestingly, our research indicates that risk of premature mortality is lower for Black, Asian, and “Other” ethnicities compared with whites in the UK Biobank, even after adjustment for a large suite of sociodemographic and deprivation factors. This mirrors previous research using national UK census and death registration data showing that life expectancy is lower for whites compared with all other ethnic groups in the UK [74]. However, these same non-white ethnic groups also tend to live in higher deprivation areas, report poorer self-rated health, and report poorer experiences of using health services in the UK [75]. More research is required to understand how these opposite forces interact in different ways to produce lower mortality risk for UK minorities. While the relationship between geographic location, area disadvantage, and mortality is well established in the UK [35], further research is also needed on how this relationship applies across ethnic groups.

Overall, cluster multivariable analyses showed robust associations across multiple, similar exposures that all have a concurrent effect on premature mortality while in the same model (e.g., multiple measures of physical activity: walking for pleasure, swimming and cycling, home DIY, general activity levels). This demonstrates that the environmental architecture of human aging is comprised of a complex network of interrelated factors that cannot be reduced to the effect of a single variable for entire conceptual categories like social integration, physical activity, and diet/lifestyle. Analyses that focus on a single measure for any of these domains may fail to capture the total effect on aging captured by these domains. For example, our results add to the growing body of evidence that multiple dimensions of physical activity have independent, additive influences on mortality risk [76,77], and that even promotion of leisure time physical activities such as walking for pleasure, gardening, and home DIY will have positive benefits in reducing mortality burden. Our results add to this body of literature by further mapping the biological mechanisms and disease pathways through which these dimensions of physical activity influence mortality.

Interestingly, we found that a variable for years of education does not remain significantly associated with mortality when put in a model with all 41 exposures identified in the cluster models. This may indicate that the effect of education on mortality is completely mediated by the other exposures identified in our analysis and does not have an independent effect on mortality outside of these mediators, although this interpretation needs further testing. Other socioeconomic exposures, such as household income, home ownership, and employment all remain significant in this full exposome model and seem to have independent effects on mortality.

Food supplements such glucosamine, folate, fish oil supplement, multivitamins emerge as protective across many age-related diseases, with the exception of folate, which has been associated previously with increased risk of all-cause mortality and cardiovascular disease [78], as well as cancer incidence [79,80], as is found in our study.

**Supplementary discussion on study limitations and strengths**

Principal strengths and limitations to this work are discussed in the main paper manuscript. Further to our discussion of self-report bias from the main article, it is important to note that a great deal of the questionnaire measures used in the UK Biobank for self-reported traits (e.g., diet) often vary from those used in other cohort study populations, which introduces the potential for variability or “vibration” [81] between effects observed in the UK Biobank and those observed in other cohorts and consortia. Another limitation to note is that the UK Biobank population is not perfectly representative of the larger UK population, as it has been shown that the main UK Biobank population is healthier and more affluent than the general population and suffered from a low recruitment rate (5.5%) [12]. Therefore, extrapolations to the entire UK population based on the results of our analyses should not be undertaken. Finally, a limitation of our approach is that we only systematically tested for linear associations in our mortality XWAS, biomarker, and disease analyses. It is likely that certain exposures have non-linear relationships with these outcomes, and we are developing future exposome-wide analysis methods that can more systematically test for non-linear relationships.

Despite these limitations, our study possesses many unique strengths that contribute greatly to the robustness of our findings. First, the scope and diversity of exposures used in our analysis is greatly expanded beyond what has normally been tested in previous XWAS and exposome studies. This has allowed us to identify associations for understudied variables in the context of aging and premature mortality. Furthermore, we are afforded relatively higher confidence in exposures we identified as related to aging in our study based on our many steps of variable filtering, disease sensitivity, and triangulation through biomarker and disease testing. Previous epidemiological studies on environmental variables in relation to all-cause mortality, including large scale cohort studies [82] and previous exposome studies [37], have not taken these precautions. Our study design allows for a more fine-grained approach to systematically addressing bias in exposome-wide studies that greatly improves our ability to minimize risk of these biases.

Furthermore, our systematic review (described in section 1.16, above) revealed that all previous exposome- and environment-wide analyses of age-related diseases and cardiometabolic risk factors (type 2 diabetes [83], cardiometabolic traits [84], lung cancer mortality [85], chronic kidney disease [86], peripheral artery disease [87], hypertension [88], and blood pressure [89]) have only added age as a fixed effect covariate to their model and have either been cross sectional or, if longitudinal, have not used age-at-risk as the timescale for analysis. These existing exposome studies are likely to suffer from residual confounding due to age-varying risk or cohort effects, even where longitudinal methods are used. Our robust approach to modeling age-varying risk with added adjustment for 5-year birth cohorts diminishes the potential of confounding from age that is widespread in existing exposome research and improves confidence in our associations reported.

We believe that exposome-wide studies using robust, multi-stage approaches such as ours not only offers the opportunity to discover new environmental influences on aging, but can help to address the preponderance of false positives and non-replicated associations evident in observational epidemiology in a manner similar to the transition to genome-wide association studies (GWAS) in genomics research [90,91]. Through using large, well-characterized study samples with adequate power; use of stringent exposome-wide significance thresholds; performing concurrent replication analyses in independent populations; and undertaking an agnostic analysis of all exposome features available in a given dataset and documenting results from all tests (both significant and null) without selective reporting, exposome science will be crucial in increasing the reproducibility of significant associations in observational epidemiology.

### Supplementary figures

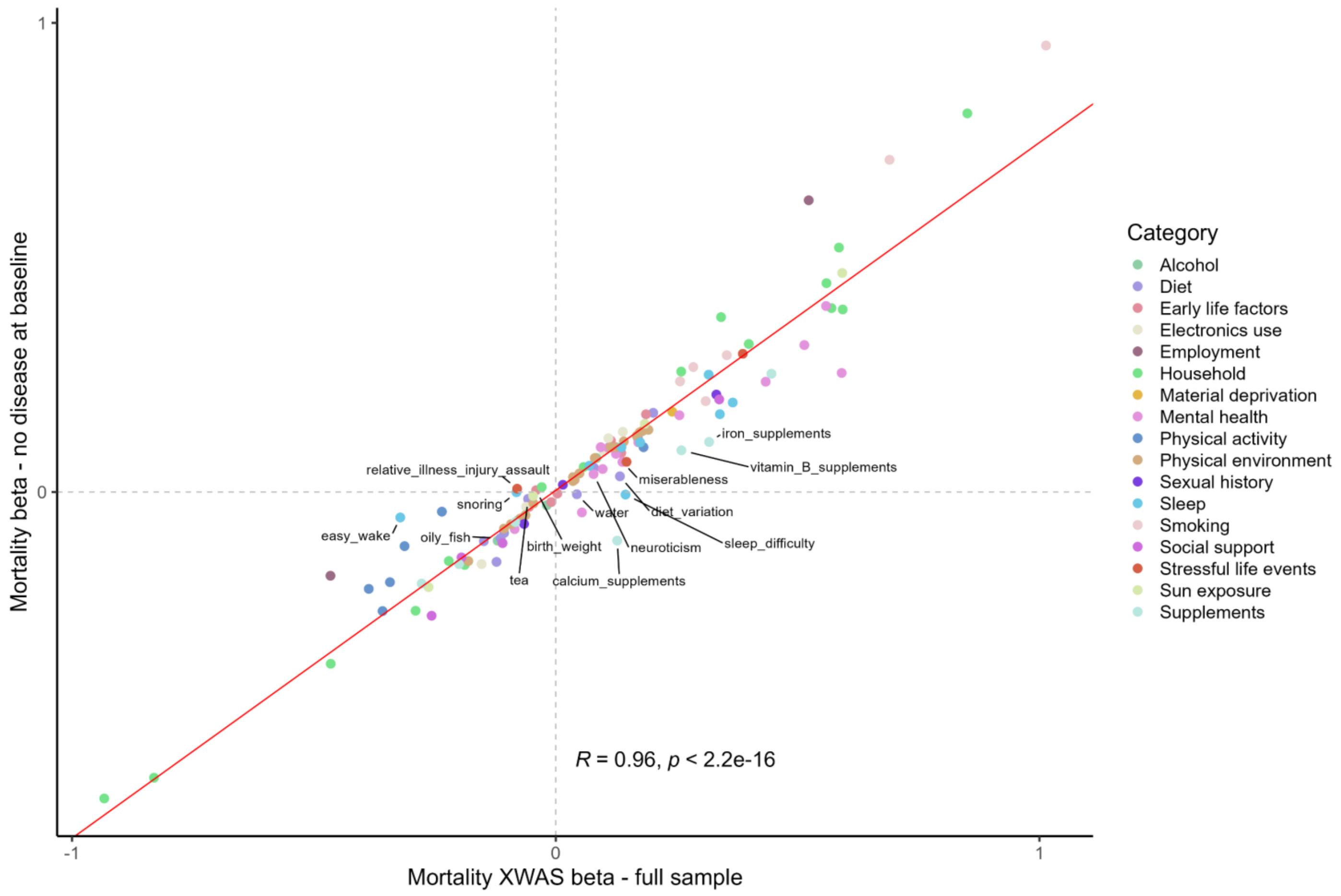

Figure S1. Correlation between mortality XWAS betas calculated in the full pooled sample including women and men (x-axis) and the subset of participants with no disease or poor health (y-axis). Pearson's R for the correlation between betas is shown, as is the p-value for the correlation. A best fit line is fitted by regressing the betas from the y-axis onto the betas from the x-axis. Labelled points are those variables that were flagged during the disease indicator interaction analysis.

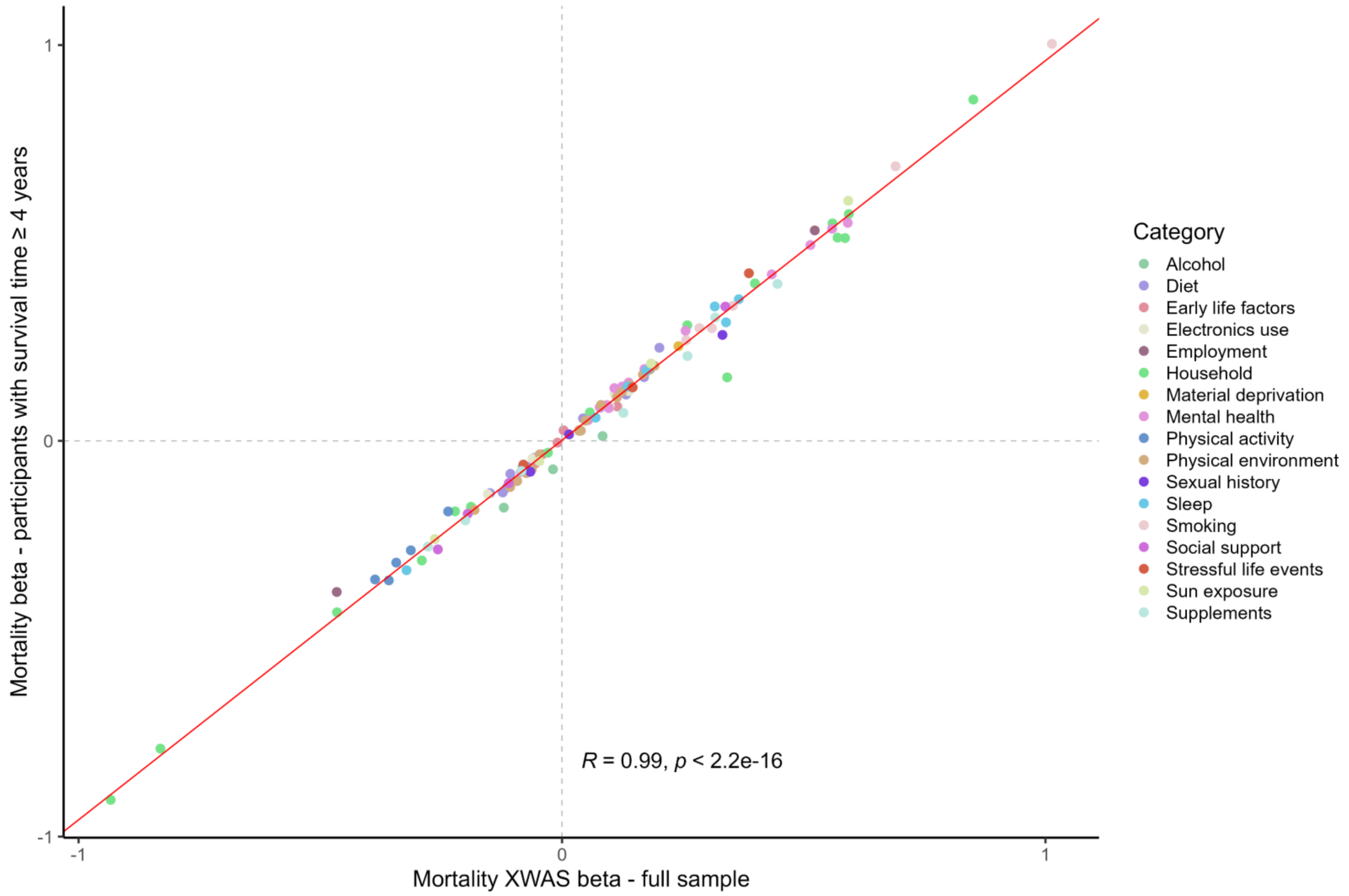

Figure S2. Correlation between mortality XWAS betas calculated in the full pooled sample (x-axis) and the subset of participants excluding those who died within the first 4 years of follow up (y-axis). Pearson's R for the correlation between betas is shown, as is the p-value for the correlation. A best fit line is fitted by regressing the betas from the y-axis onto the betas from the x-axis.

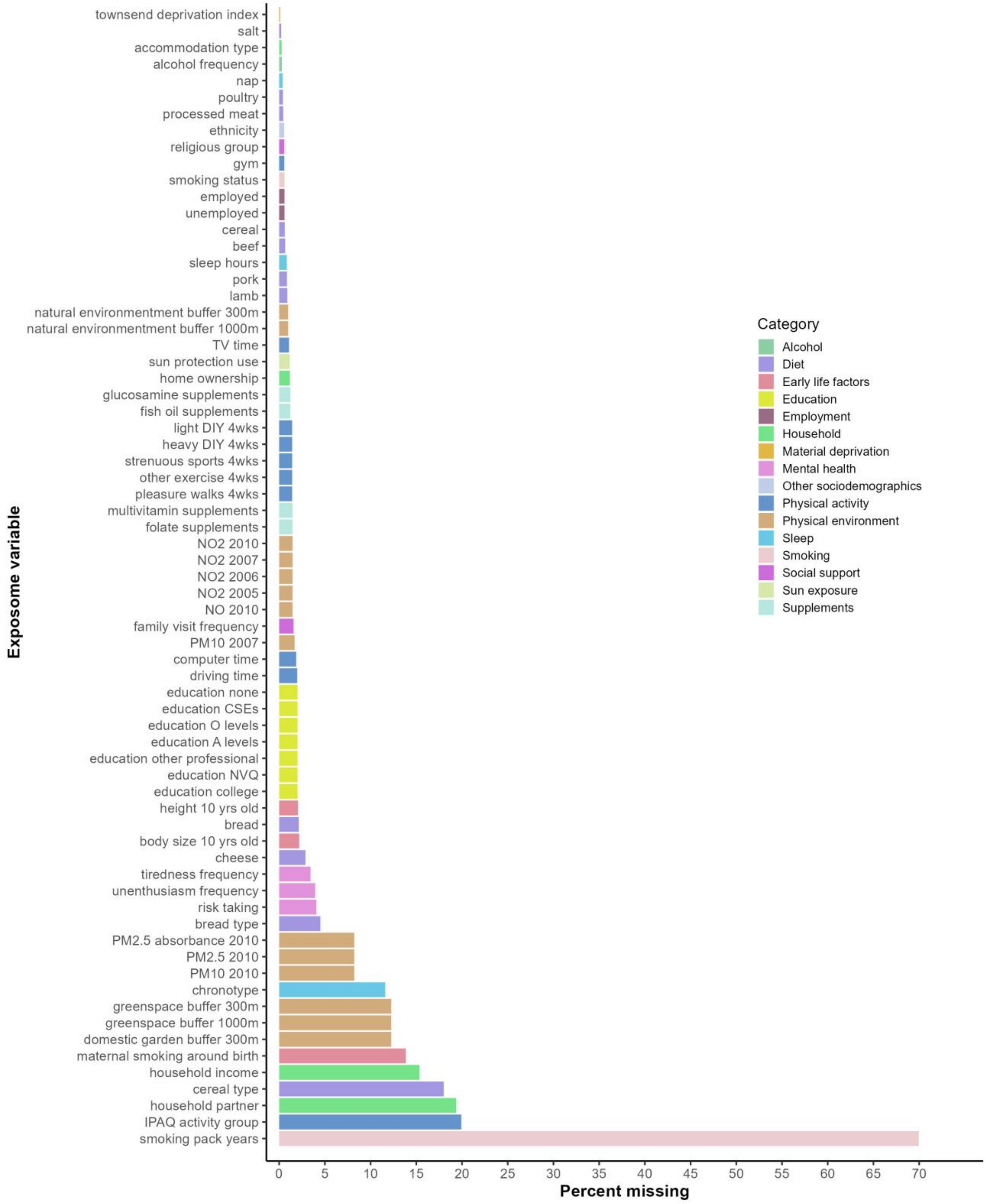

Figure S3. Rates of missing information among the 41 exposures replicated in the mortality XWAS and that survived the disease sensitivity and cluster modeling steps. Derived variables are not shown (they were only created after multiple imputation) and instead all component variables that make up derived variables are shown

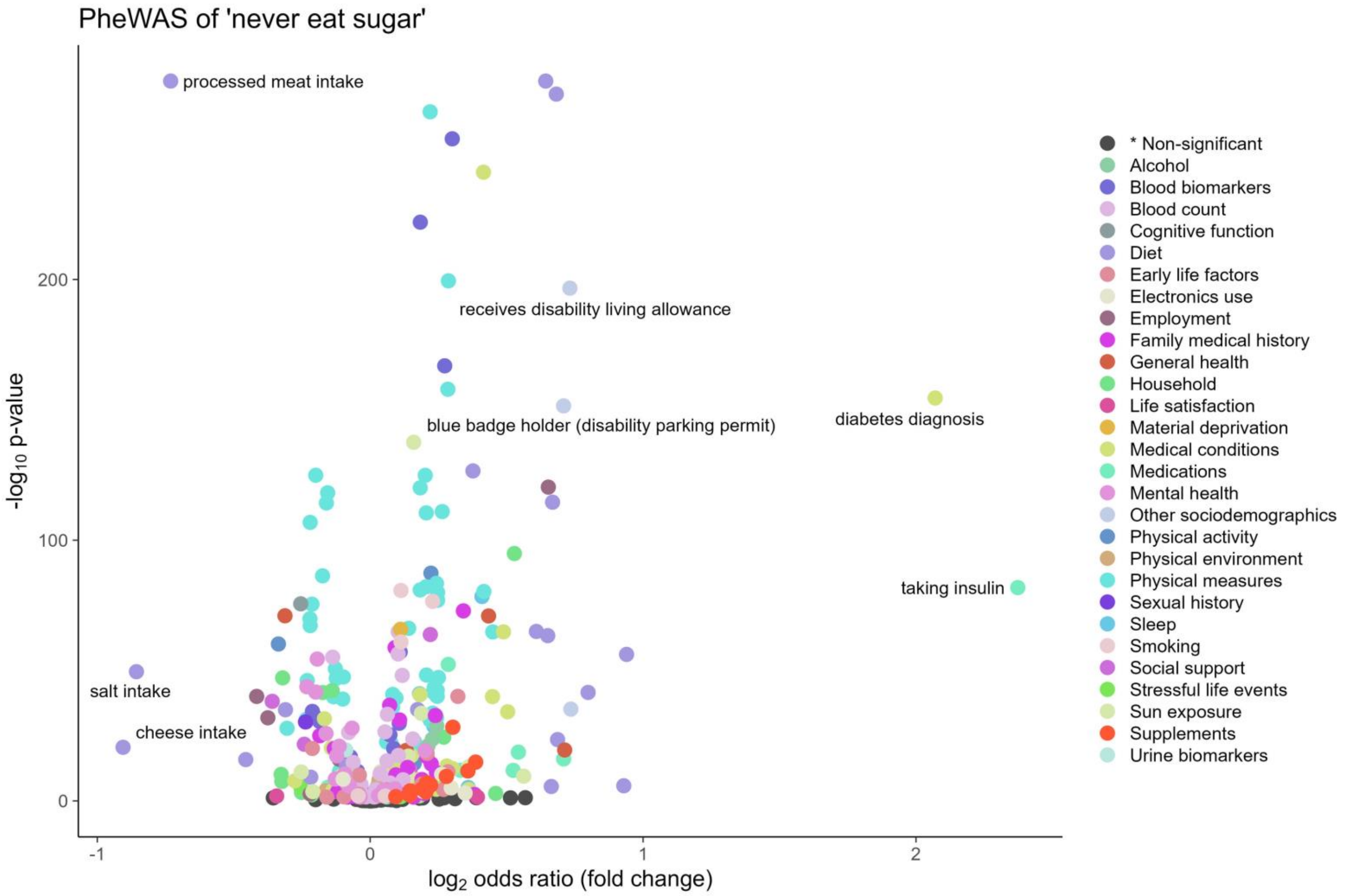

Figure S4. Volcano plot of log-transformed p-values and fold change (calculated as $\log_2$ of the odds ratio) from a PheWAS of never eating sugar in participants recruited in England (n=436,891). Each point represents the effect and p-value for the association between a single exposure and never eating sugar. Exposures that were FDR significant are colored, whereas associations that were not significant are colored dark grey and grouped in the category "* Non-significant."

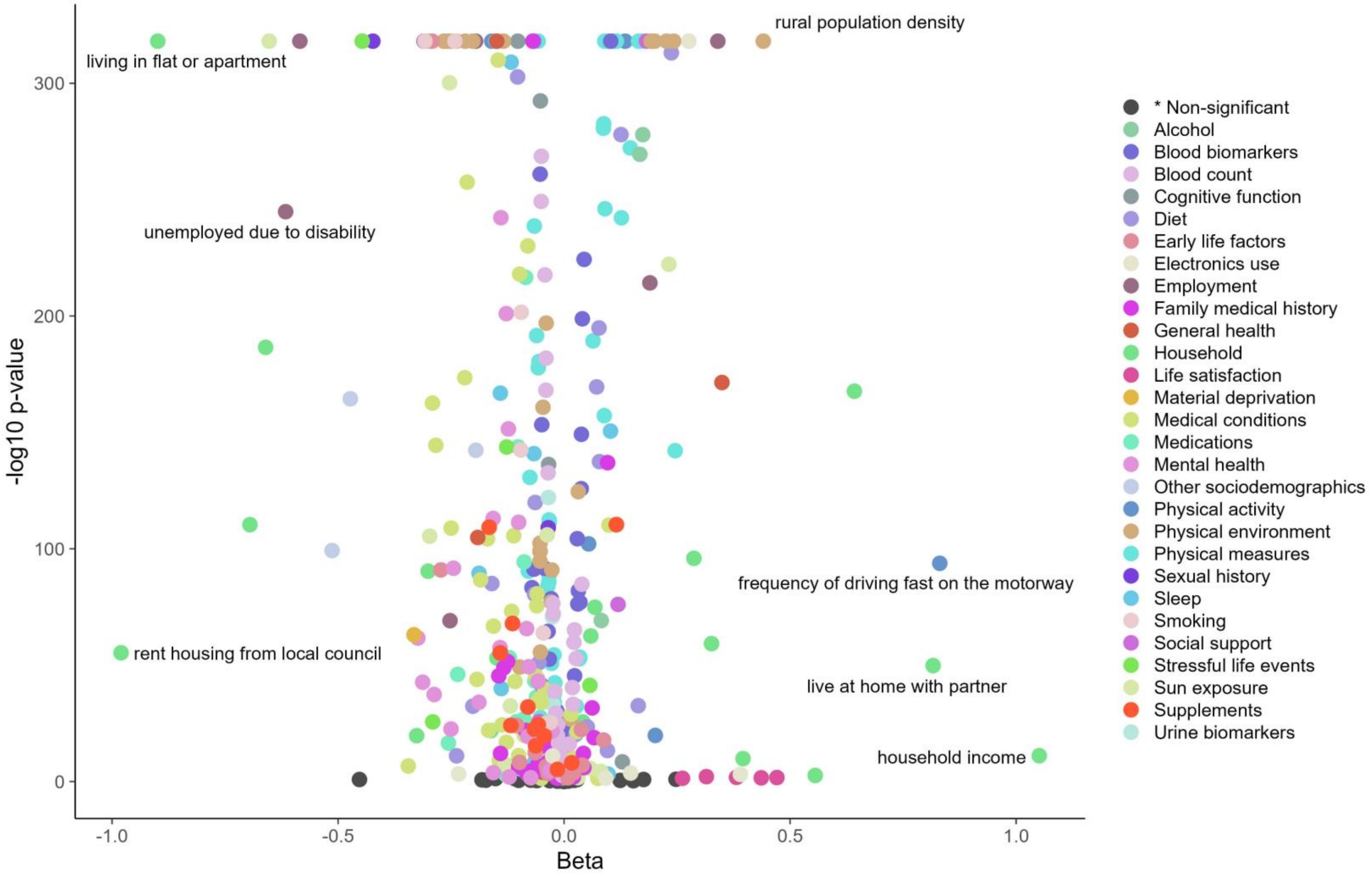

Figure S5. Volcano plot of log-transformed p-values and fold change (calculated as $\log_2$ of the odds ratio) from a PheWAS of number of household vehicles in participants recruited in England (n=436,891). Each point represents the effect and p-value for the association between a single exposure and number of household vehicles. Exposures that were FDR significant are colored, whereas associations that were not significant are colored dark grey and grouped in the category "* Non-significant."

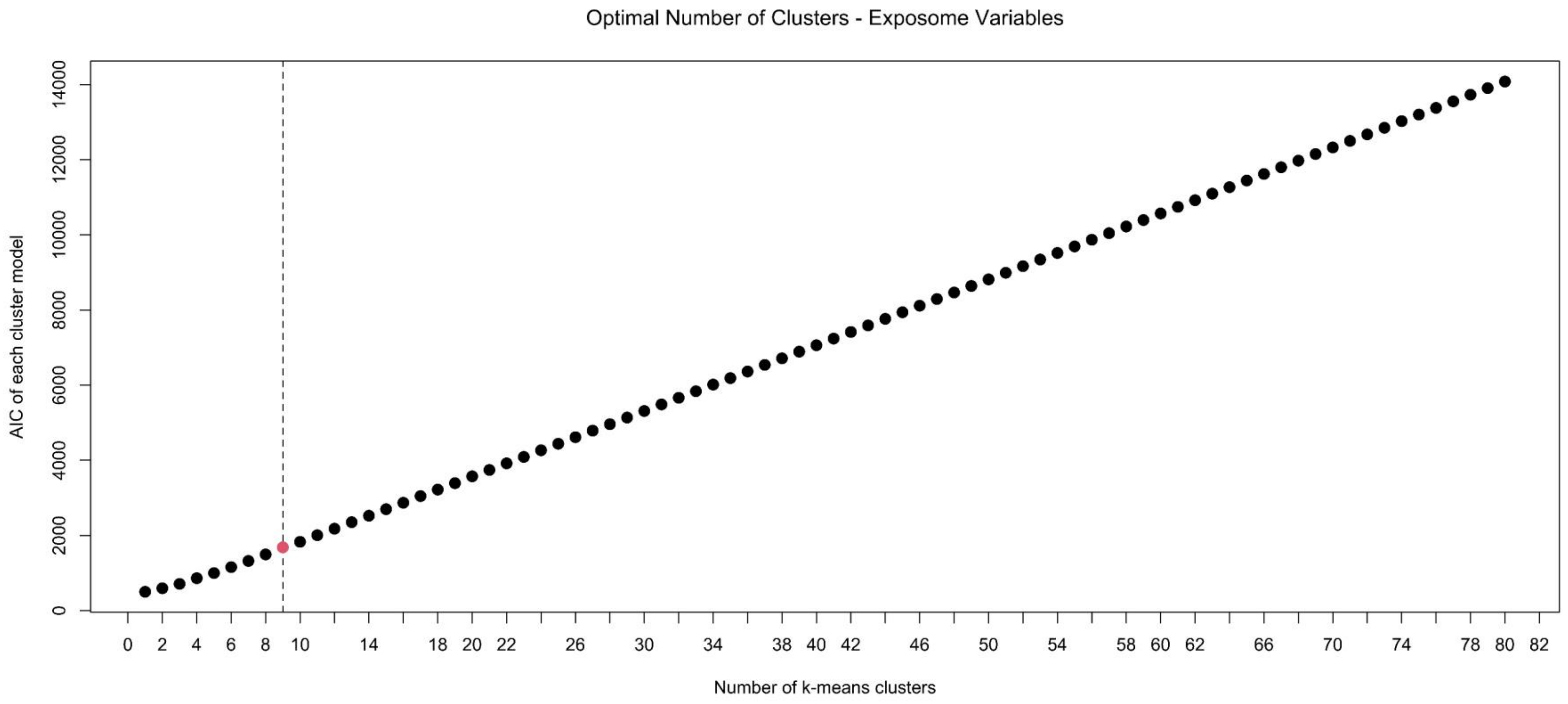

Figure S6. AIC for the cluster model (y-axis) according to the number of k-means clusters (y-axis) for exposures replicated in the pooled mortality XWAS. The black dotted line at 11 shows the number of clusters detected with an optimal AIC according to k-means.

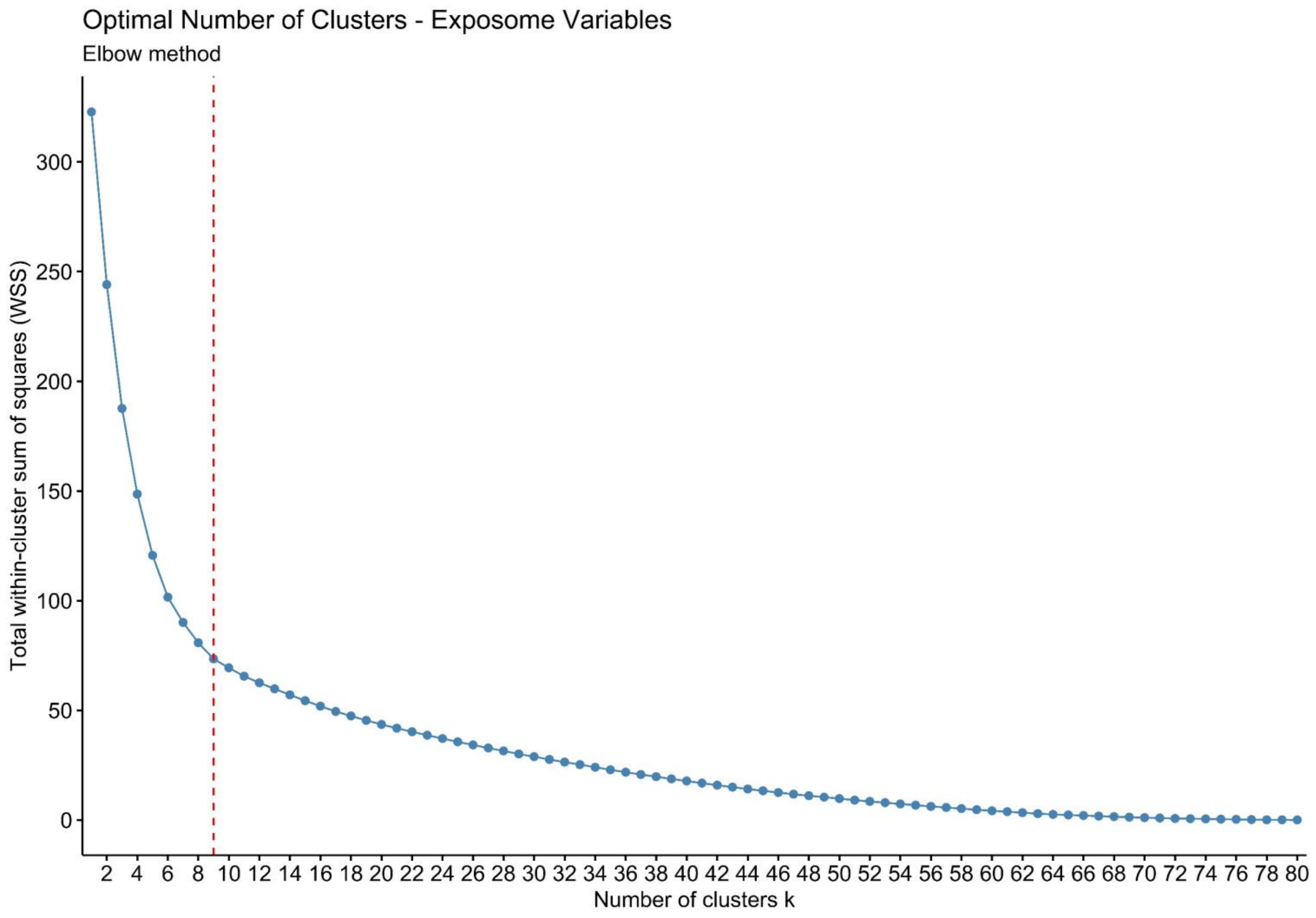

Figure S7. Total within-cluster sum of squares (WSS; y-axis) according to the number of hierarchical clusters (y-axis) for exposures replicated in the pooled mortality XWAS. The red dotted line at 9 shows the number of clusters selected by AIC.

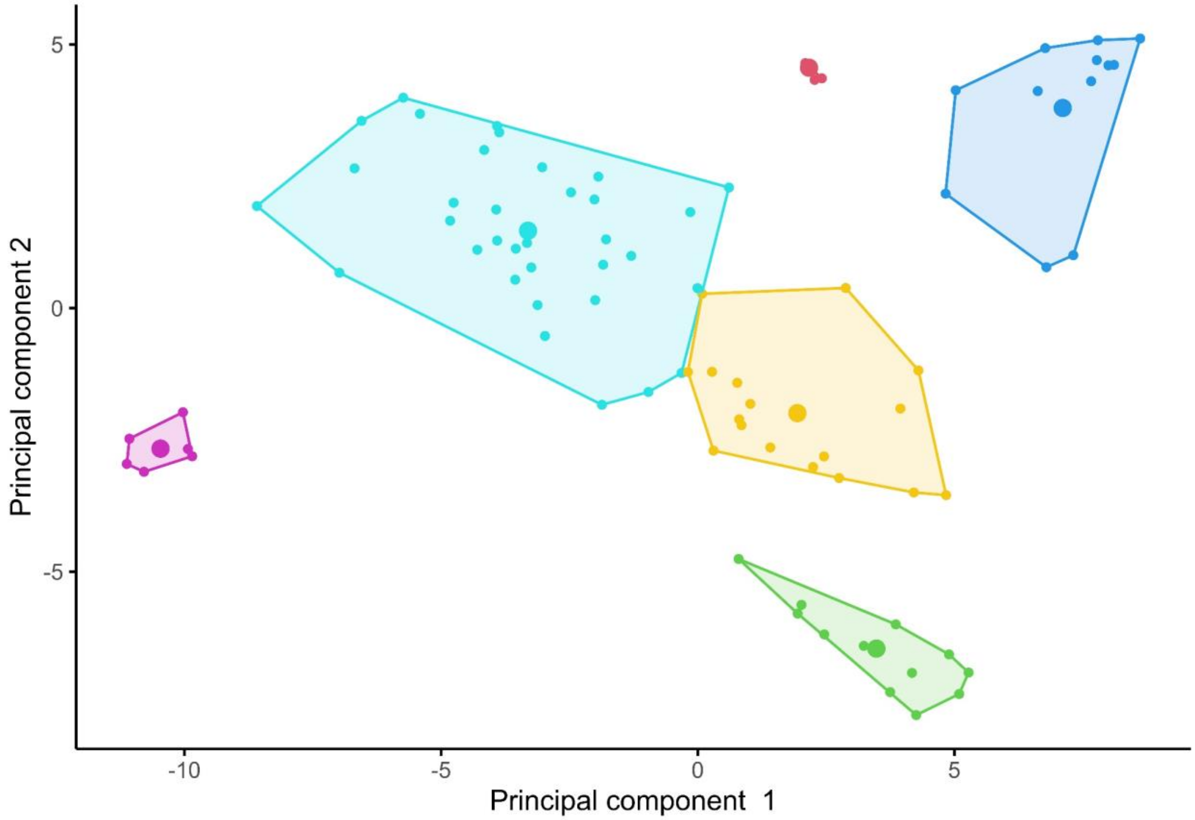

Figure S8. K-means clustering of exposures replicated in the pooled mortality XWAS using a 6-cluster solution. Each dot represents an individual exposure, with the larger dot being the centroid of each k-means cluster. X- and y-axes are the first two principal components taking the correlation matrix of all the exposures as an input.

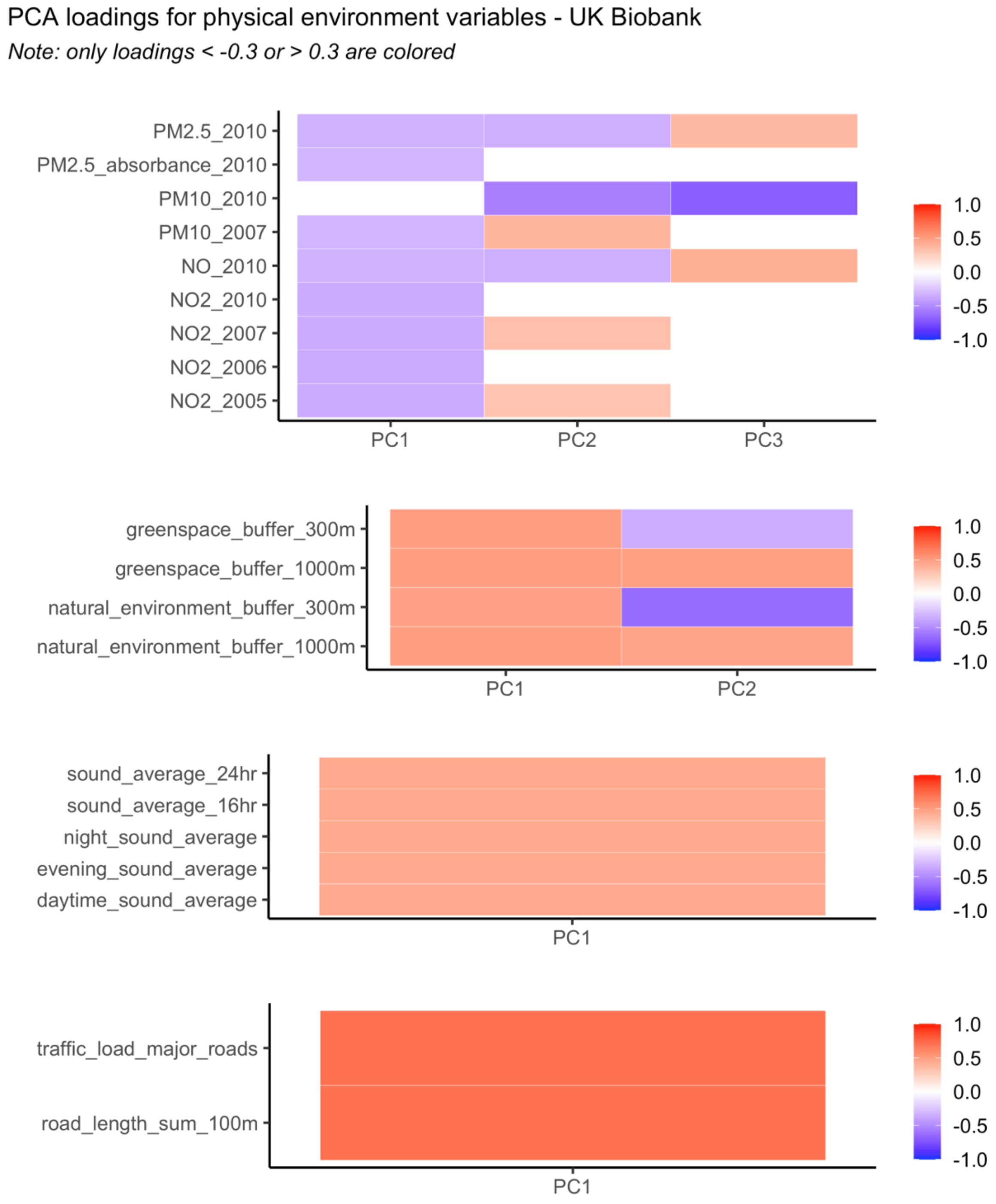

Fig. S9. Principal component (PC) loadings for highly correlated physical environment variables among \ UK Biobank participants recruited in England (n=436,891). Since all air pollution levels loaded inversely onto the first air pollution PC, the first PC was multiplied by -1 before analysis in order to reflect increasing levels of air pollution in line with the original direction of the component variables.

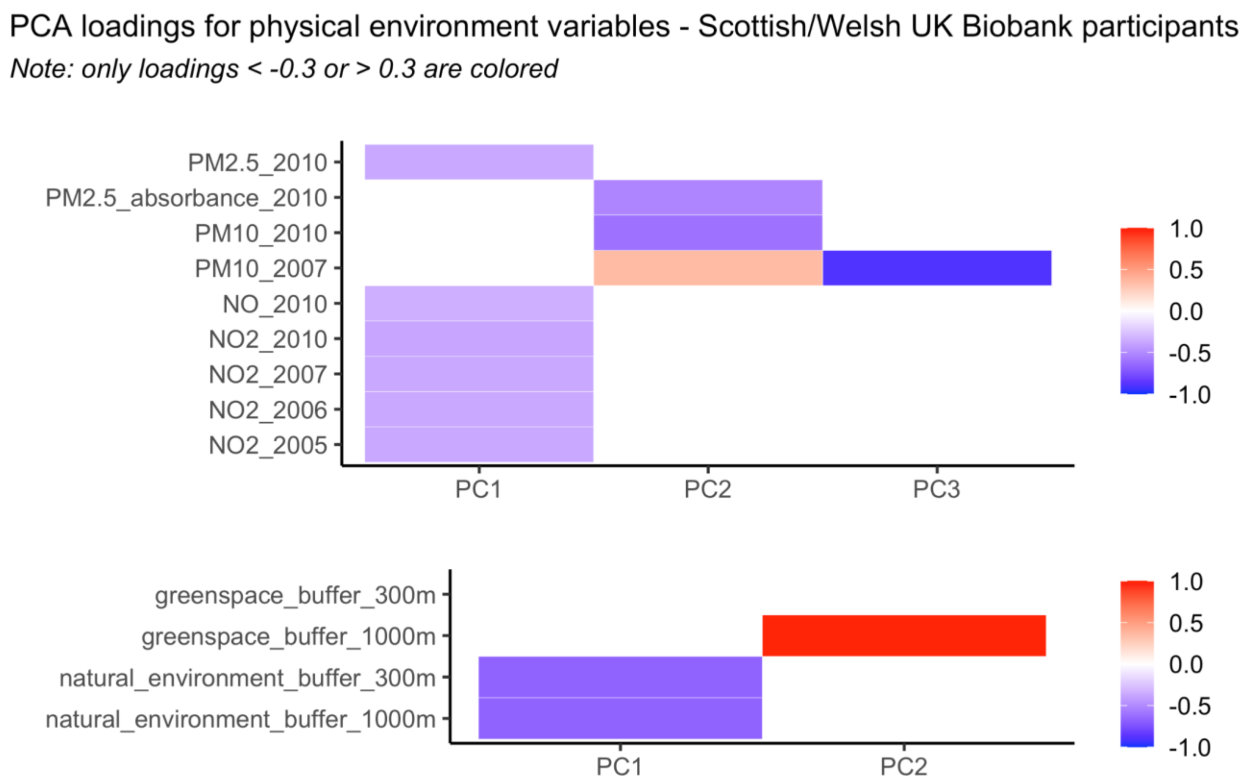

Fig. S10. Principal component loadings for highly correlated physical environment variables among UK Biobank participants recruited in Scotland/Wales (n=55,676). Principal components were only made for categories of variables that were significant in the cluster multivariable modeling carried out in participants recruited in England (n=436,891). Since all air pollution levels loaded inversely onto the first air pollution PC, the first PC was multiplied by -1 before analysis in order to reflect increasing levels of air pollution in line with the original direction of the component variables.

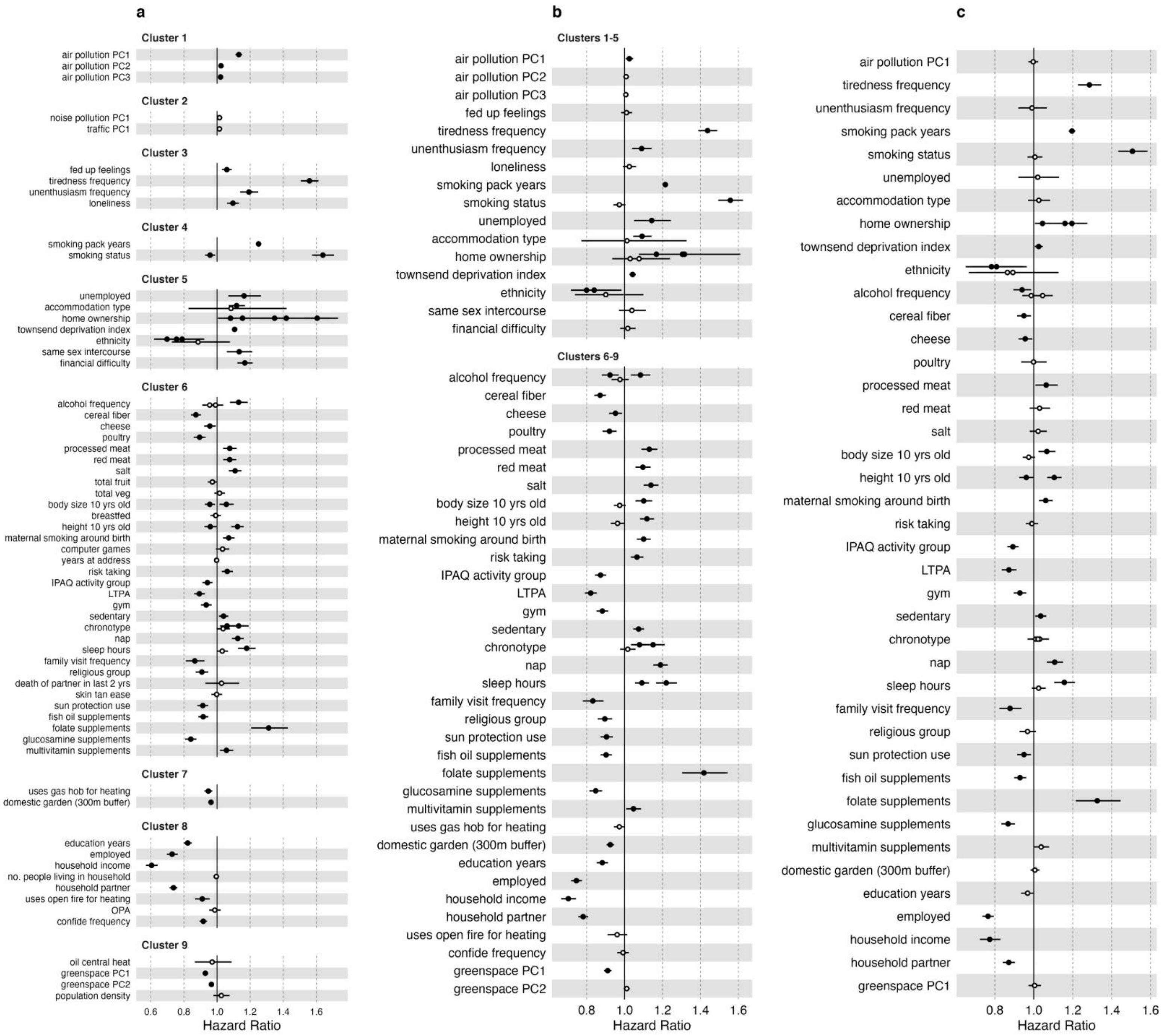

a
Cluster 1
air pollution PC1
air pollution PC2
air pollution PC3
Cluster 2
noise pollution PC1
traffic PC1
Cluster 3
fed up feelings
tiredness frequency
unenthusiasm frequency
loneliness
Cluster 4
smoking pack years
smoking status
Cluster 5
unemployed
accommodation type
home ownership
townsend deprivation index
ethnicity
same sex intercourse
financial difficulty
Cluster 6
alcohol frequency
cereal fiber
cheese
poultry
processed meat
red meat
salt
total fruit
total veg
body size 10 yrs old
breastfed
height 10 yrs old
maternal smoking around birth
computer games
years at address
risk taking
IPAQ activity group
LTPA
gym
sedentary
chronotype
nap
sleep hours
family visit frequency
religious group
death of partner in last 2 yrs
skin tan ease
sun protection use
fish oil supplements
folate supplements
glucosamine supplements
multivitamin supplements
Cluster 7
uses gas hob for heating
domestic garden (300m buffer)
Cluster 8
education years
employed
household income
no. people living in household
household partner
uses open fire for heating
OPA
confide frequency
Cluster 9
oil central heat
greenspace PC1
greenspace PC2
population density
0.6
0.8
1.0
1.2
1.4
1.6
Hazard Ratio
b
Clusters 1-5
air pollution PC1
air pollution PC2
air pollution PC3
fed up feelings
tiredness frequency
unenthusiasm frequency
loneliness
smoking pack years
smoking status
unemployed
accommodation type
home ownership
townsend deprivation index
ethnicity
same sex intercourse
financial difficulty
Clusters 6-9
alcohol frequency
cereal fiber
cheese
poultry
processed meat
red meat
salt
body size 10 yrs old
height 10 yrs old
maternal smoking around birth
risk taking
IPAQ activity group
LTPA
gym
sedentary
chronotype
nap
sleep hours
family visit frequency
religious group
sun protection use
fish oil supplements
folate supplements
glucosamine supplements
multivitamin supplements
uses gas hob for heating
domestic garden (300m buffer)
education years
employed
household income
household partner
uses open fire for heating
confide frequency
greenspace PC1
greenspace PC2
0.8
1.0
1.2
1.4
1.6
Hazard Ratio
c
air pollution PC1
tiredness frequency
unenthusiasm frequency
smoking pack years
smoking status
unemployed
accommodation type
home ownership
townsend deprivation index
ethnicity
alcohol frequency
cereal fiber
cheese
poultry
processed meat
red meat
salt
body size 10 yrs old
height 10 yrs old
maternal smoking around birth
risk taking
IPAQ activity group
LTPA
gym
sedentary
chronotype
nap
sleep hours
family visit frequency
religious group
sun protection use
fish oil supplements
folate supplements
glucosamine supplements
multivitamin supplements
domestic garden (300m buffer)
education years
employed
household income
household partner
greenspace PC1
0.8
1.0
1.2
1.4
1.6
Hazard Ratio

Figure S11. Forest plot of exposome associations with all-cause mortality (n= 436,891) in **(a)** multivariable models within each individual cluster, **(b)** multivariable models within grouped neighboring clusters, and **(c)** final exposome-wide model used for C-index and $R^2$ calculations. Only variables significant at $p < 0.05$ were carried forward to the next model. Models in **(a)** and **(b)** were Cox models calculated using age as the timescale, stratified by 5-year birth cohorts and sex, and adjusted for UK Biobank assessment center, years of education, household income, and ethnicity (only if the covariate was not already in the cluster model). The full exposome $R^2$ model **(c)** uses a single Cox model with mortality survival time as the timescale and adding age (scaled) and sex as fixed covariates (estimates not shown for age and sex in the forest plot). Multiple estimates are shown for nominal categorical variables, with estimate points for each response level in the same row. Estimates not significant at $p < 0.05$ are shown as hollow points. IPAQ: International Physical Activity Questionnaires; LTPA: leisure time physical activity; MH: mental health; OPA: occupational physical activity; PC: principal component

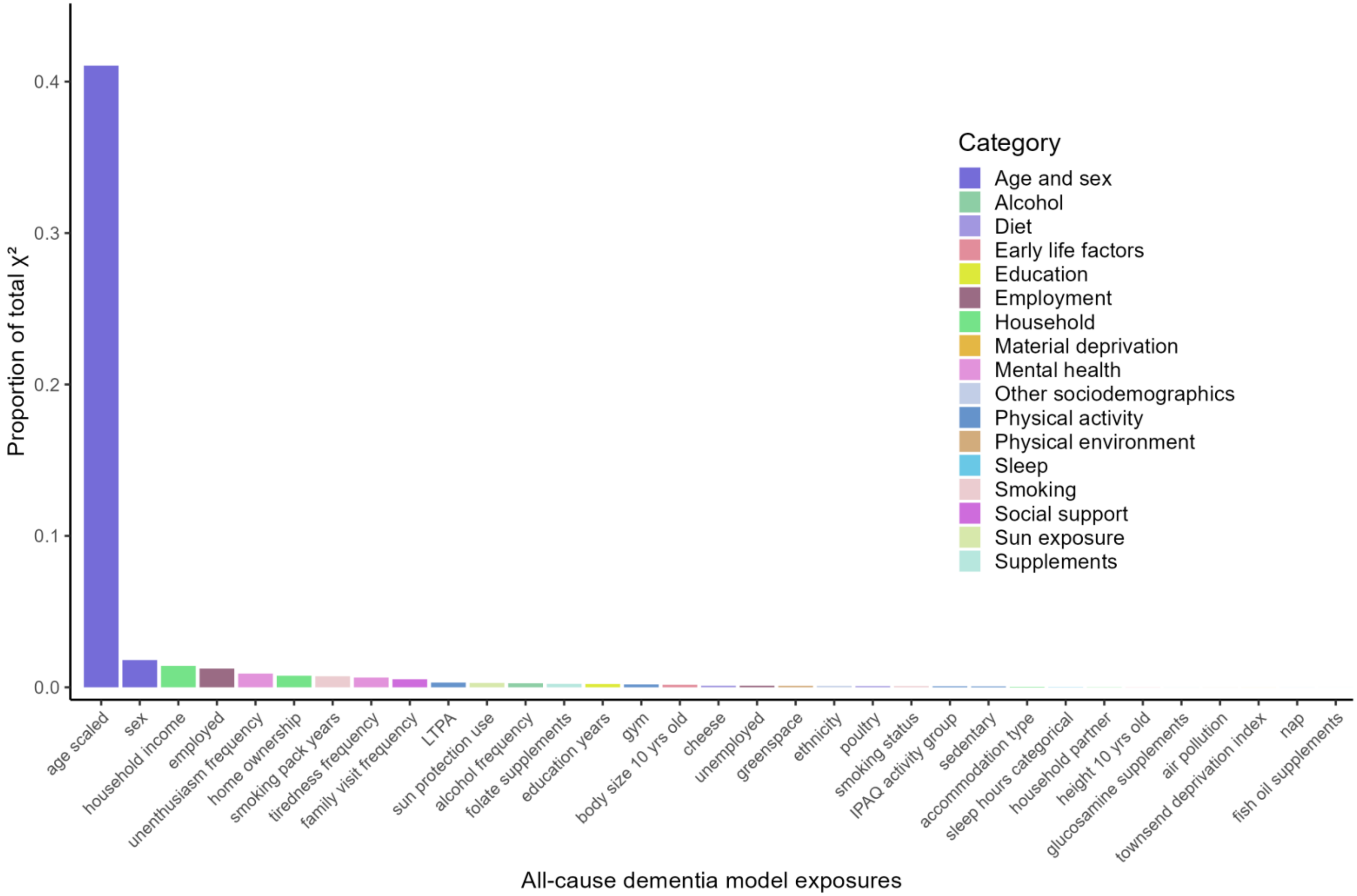

Figure S12. Variable importance according to proportion of total model chi-squared ($X^2$) for each exposure in the final multivariable all-cause dementia exposome Cox model.

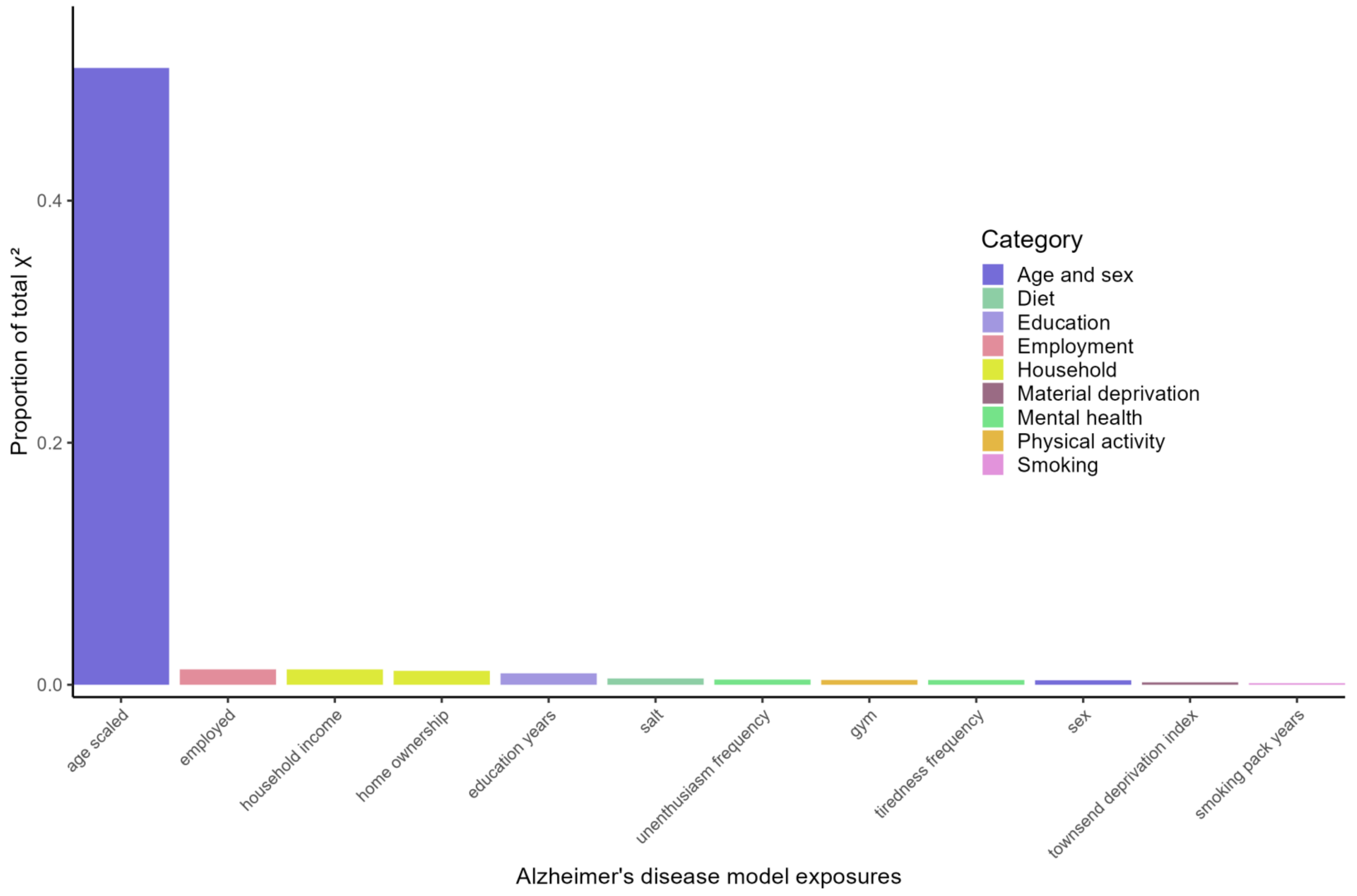

Figure S13. Variable importance according to proportion of total model chi-squared ($X^2$) for each exposure in the final multivariable Alzheimer's disease exposome Cox model.

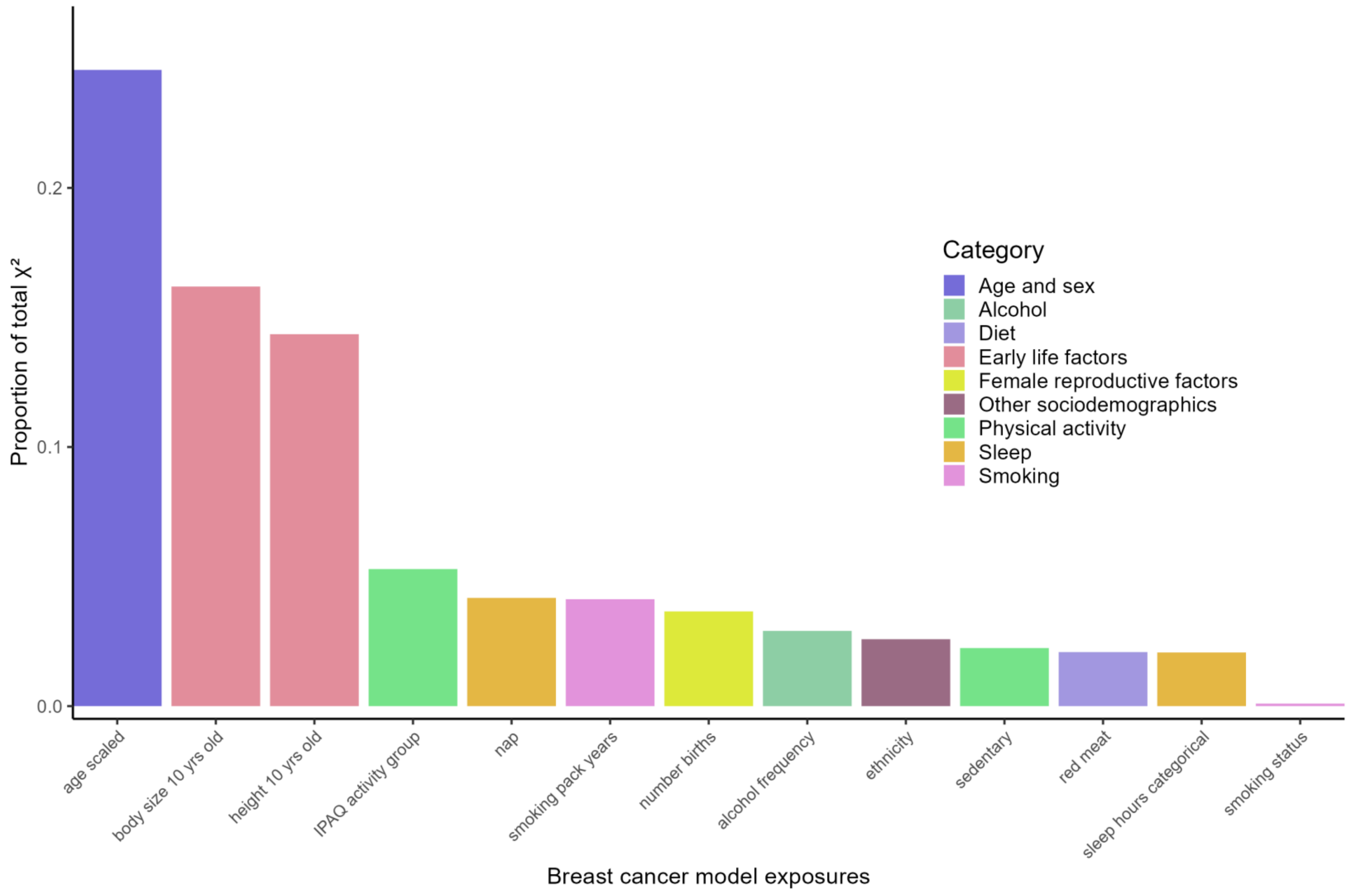

Figure S14. Variable importance according to proportion of total model chi-squared ($X^2$) for each exposure in the final multivariable Breast cancer exposome Cox model.

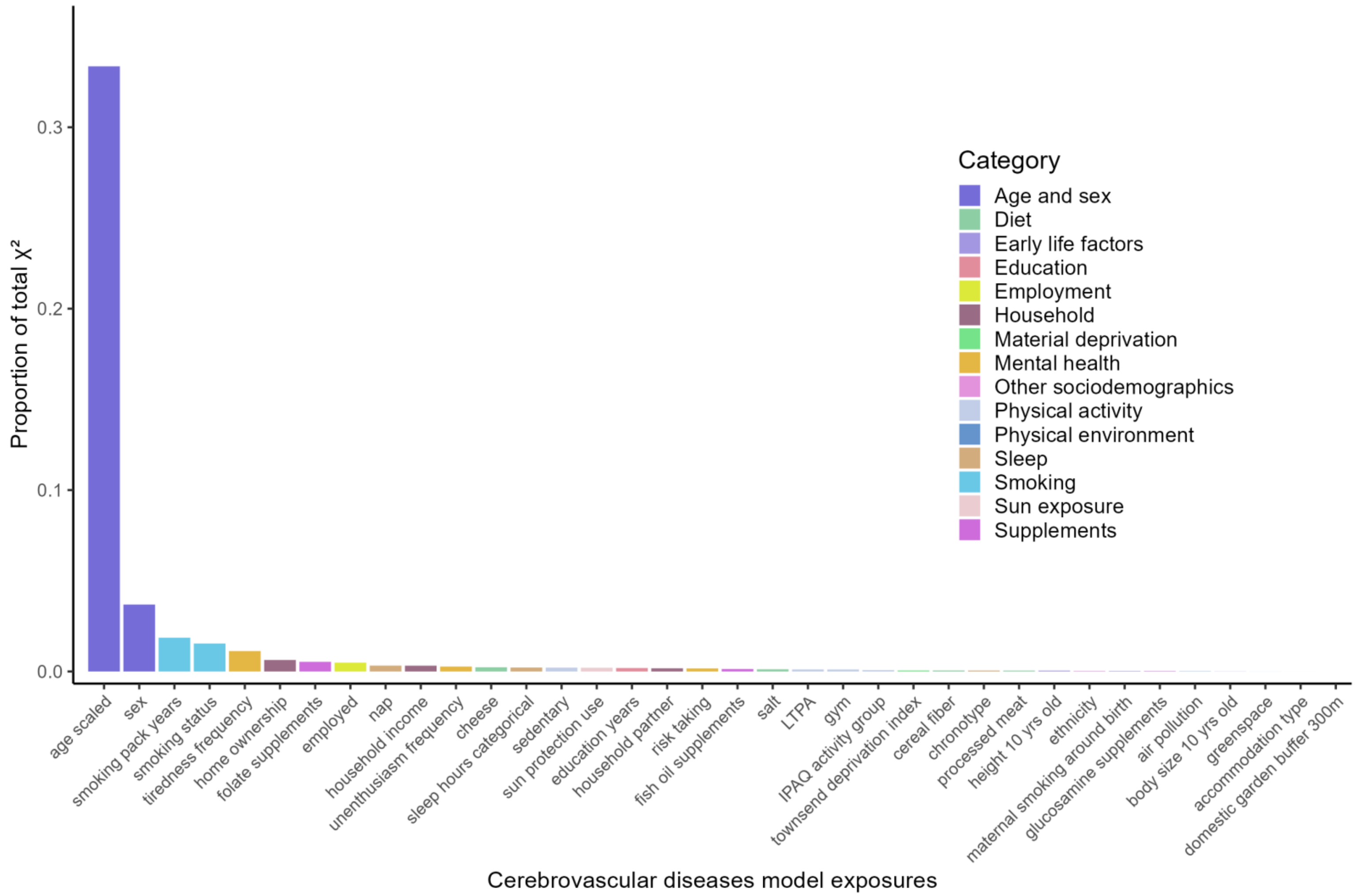

Figure S15. Variable importance according to proportion of total model chi-squared ($X^2$) for each exposure in the final multivariable cerebrovascular diseases exposome Cox model.

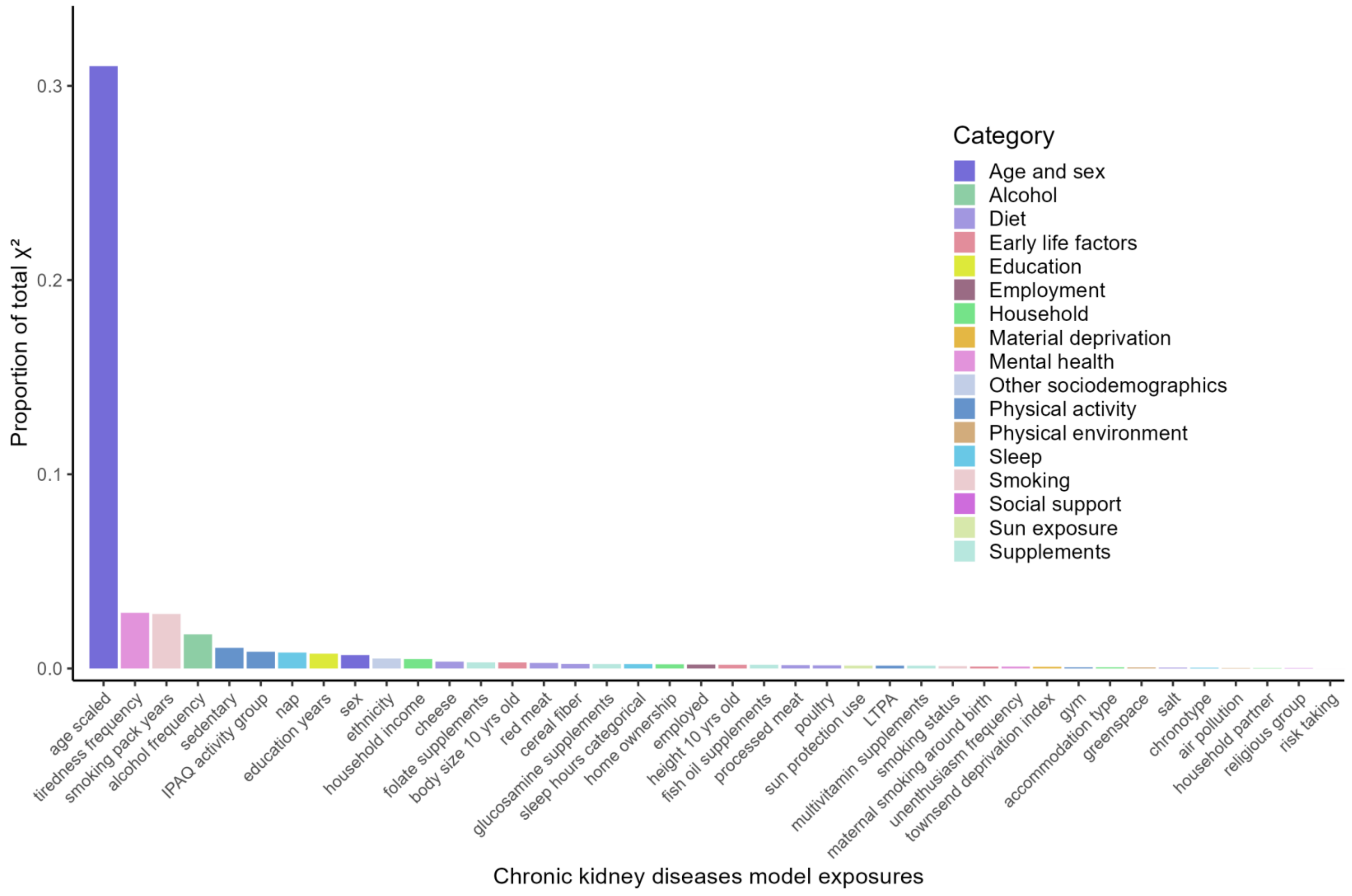

Figure S16. Variable importance according to proportion of total model chi-squared ($X^2$) for each exposure in the final multivariable chronic kidney diseases exposome Cox model.

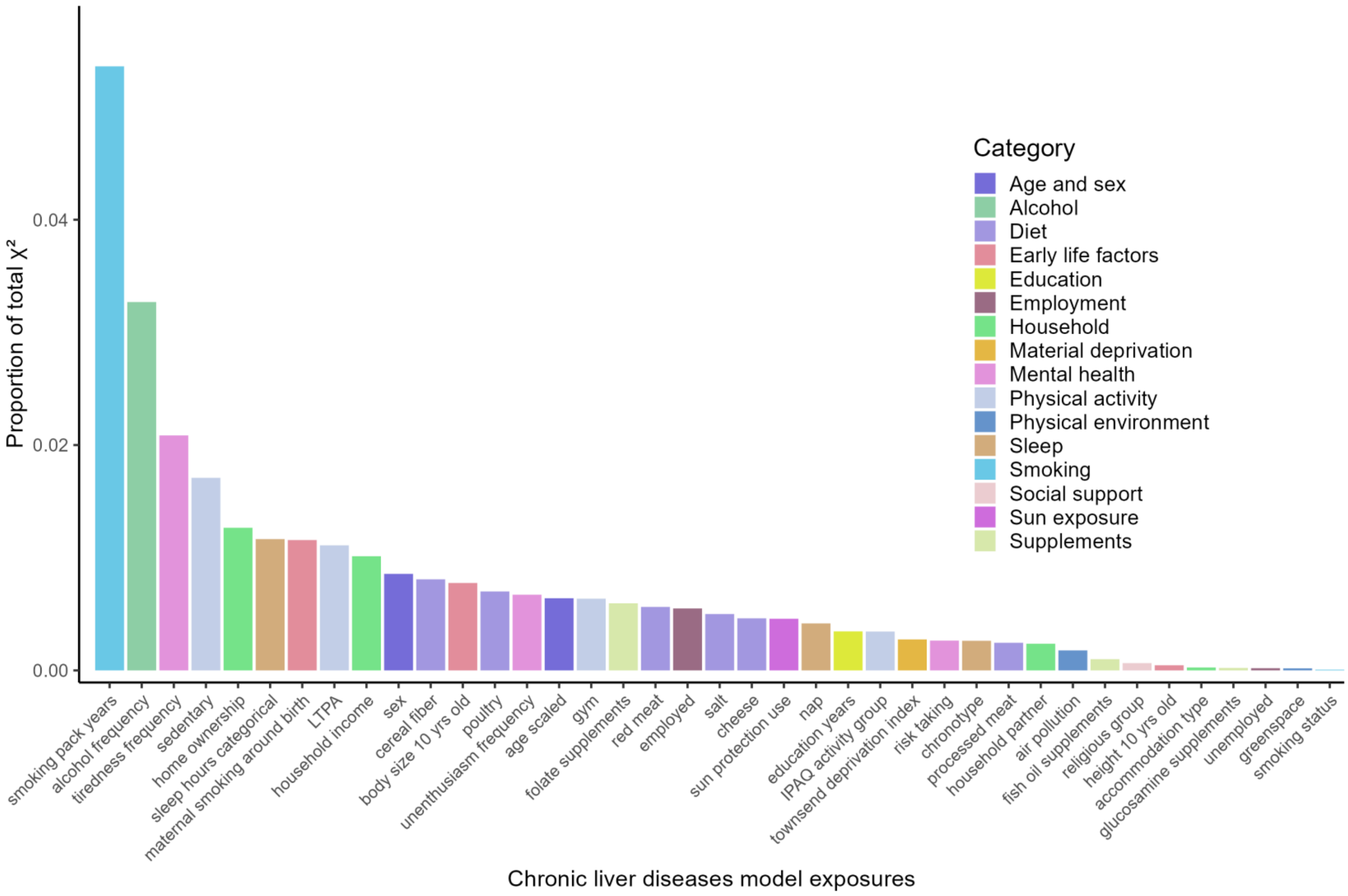

Figure S17. Variable importance according to proportion of total model chi-squared ($X^2$) for each exposure in the final multivariable chronic liver diseases exposome Cox model.

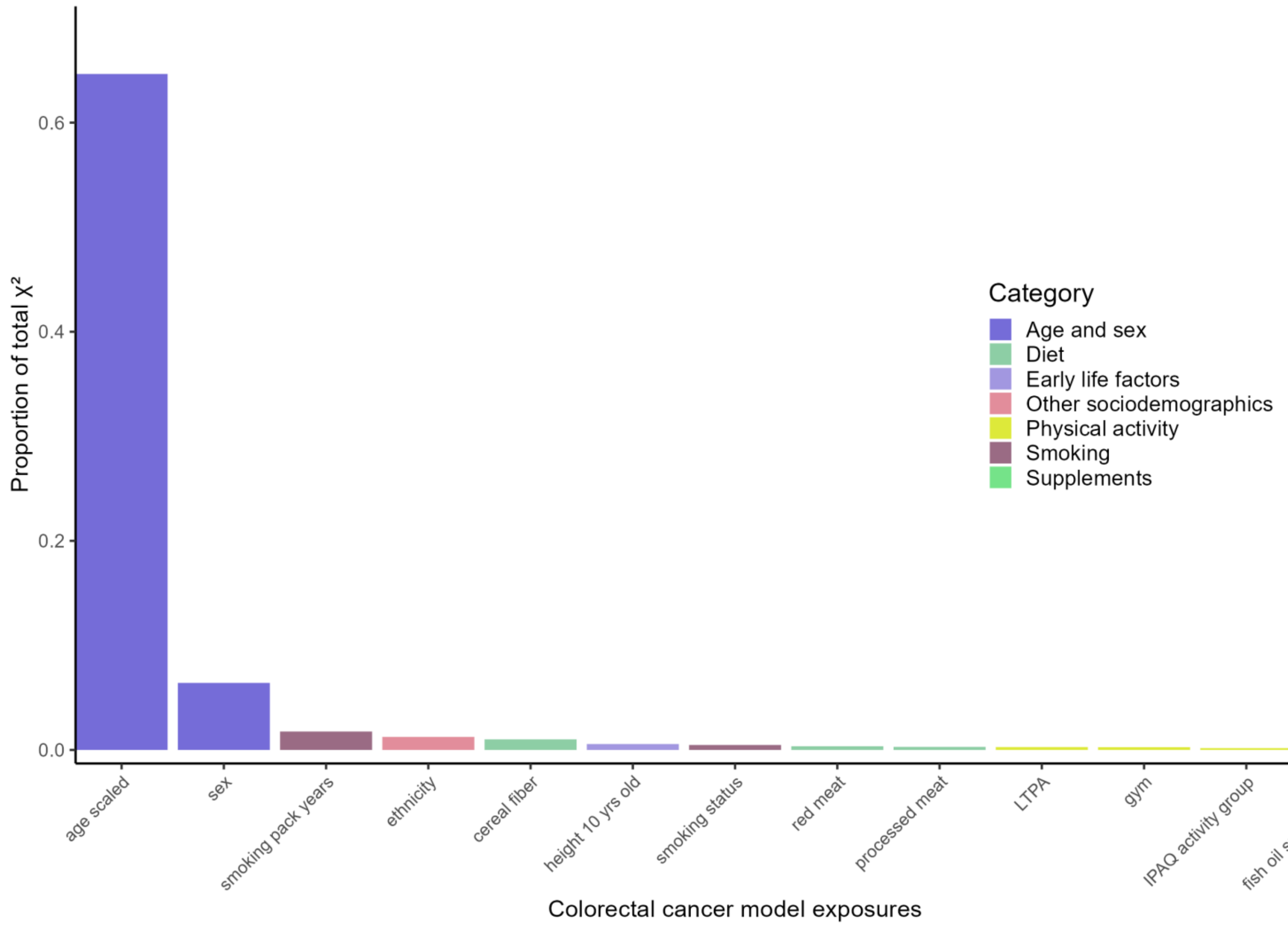

Figure S18. Variable importance according to proportion of total model chi-squared ($X^2$) for each exposure in the final multivariable colorectal cancer exposome Cox model.

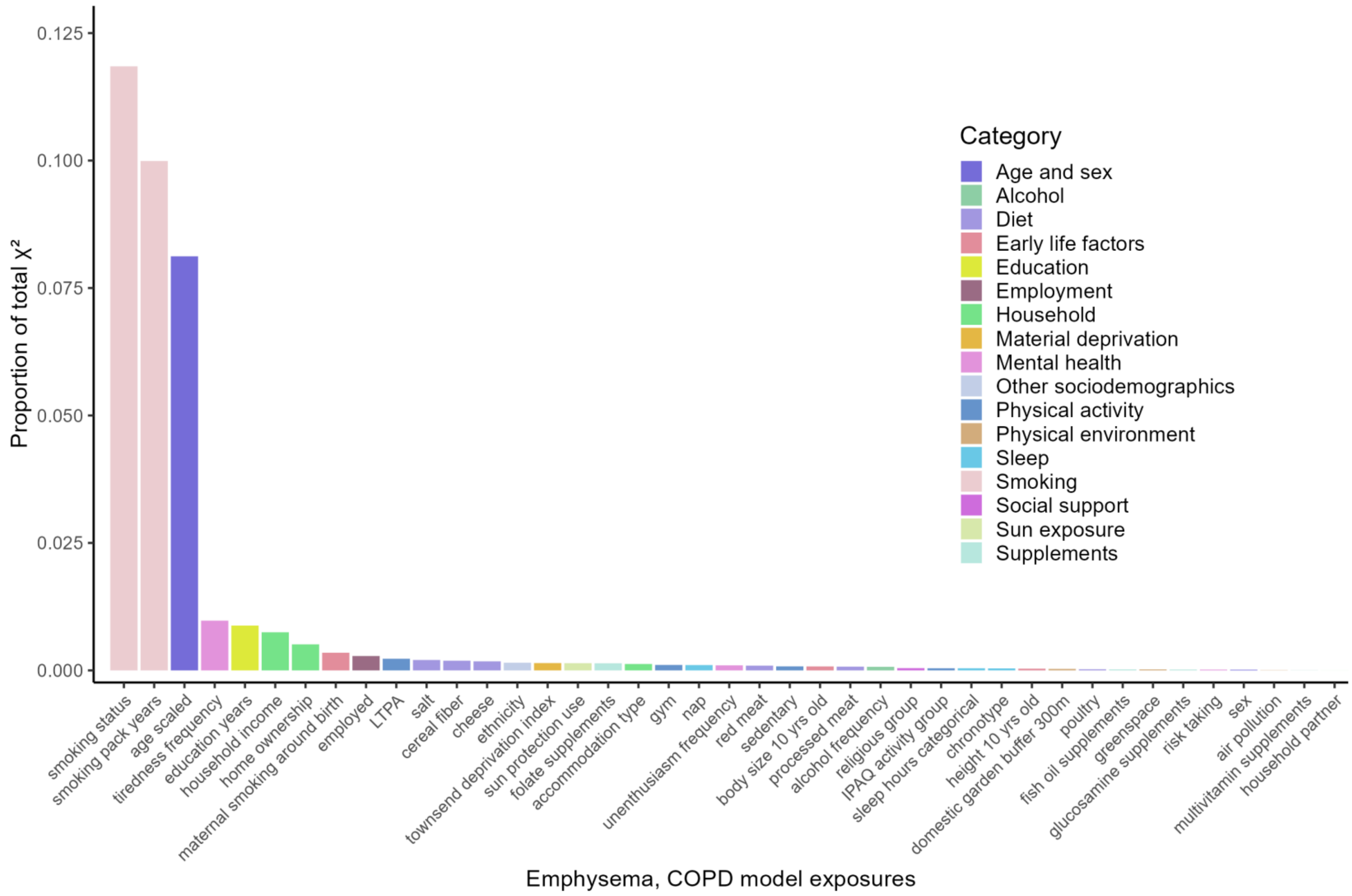

Figure S19. Variable importance according to proportion of total model chi-squared ($X^2$) for each exposure in the final multivariable emphysema, COPD exposome Cox model.

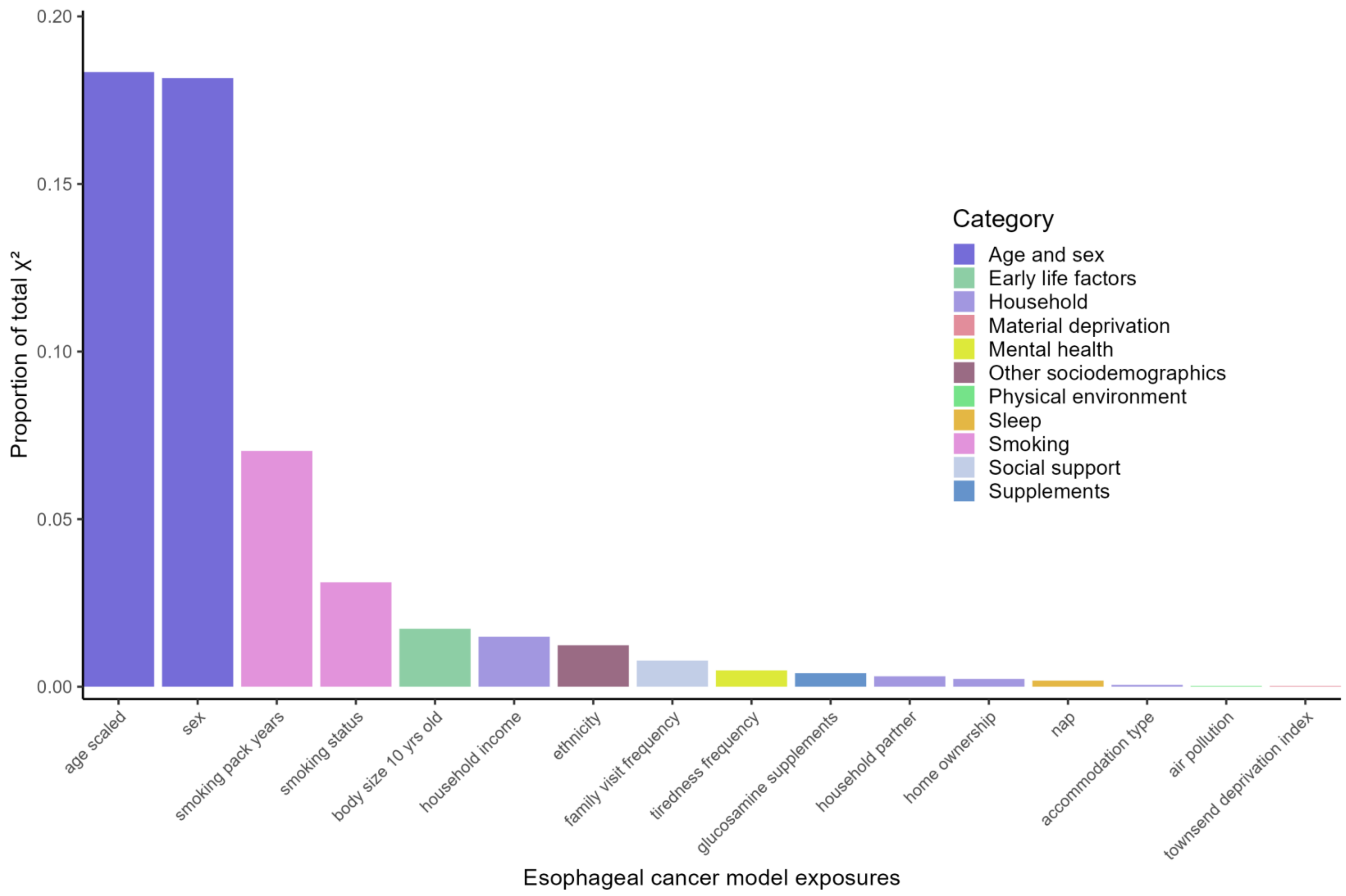

Figure S20. Variable importance according to proportion of total model chi-squared ($X^2$) for each exposure in the final multivariable esophageal cancer exposome Cox model.

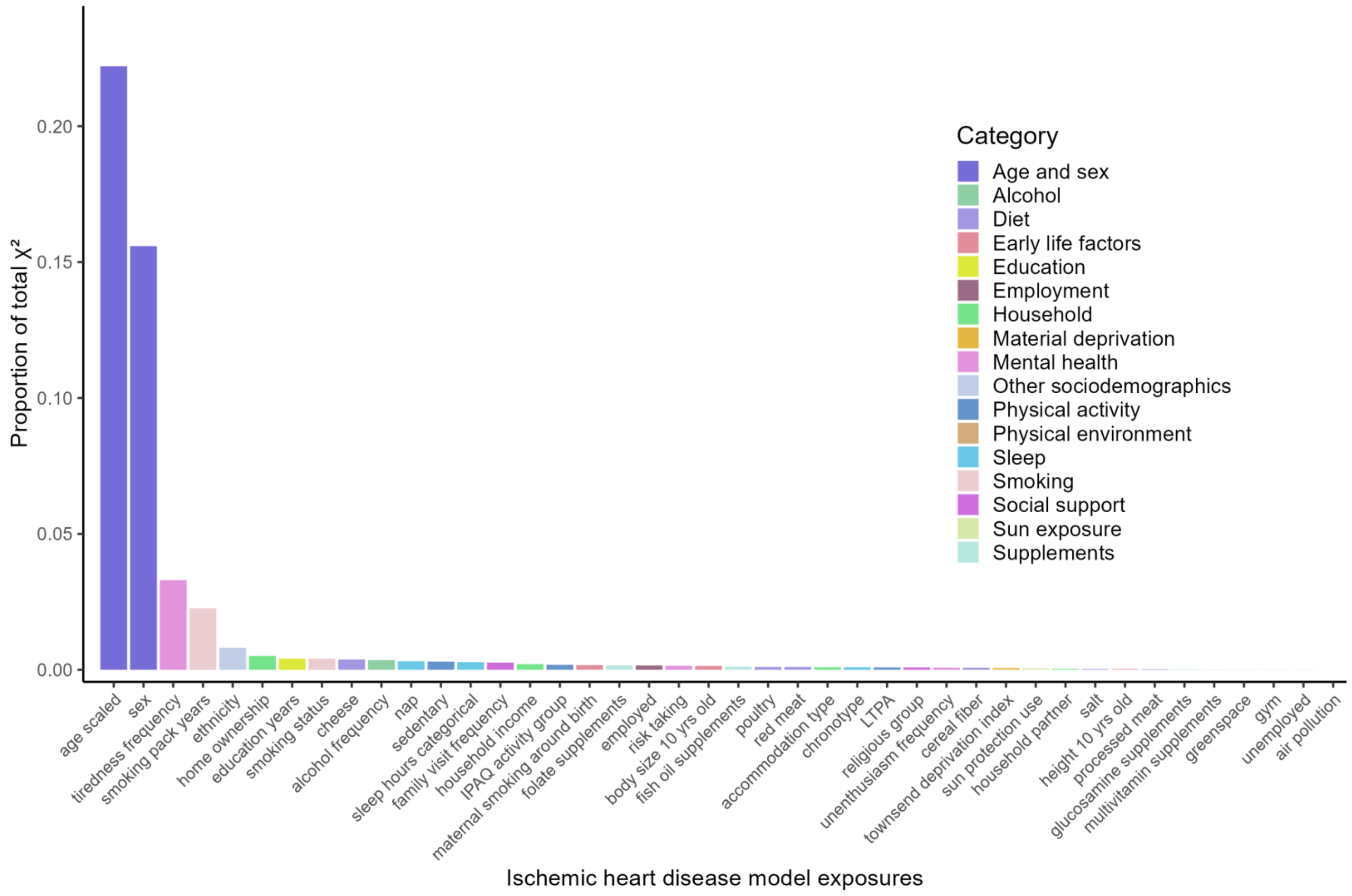

Figure S21. Variable importance according to proportion of total model chi-squared ($X^2$) for each exposure in the final multivariable ischemic heart disease exposome Cox model.

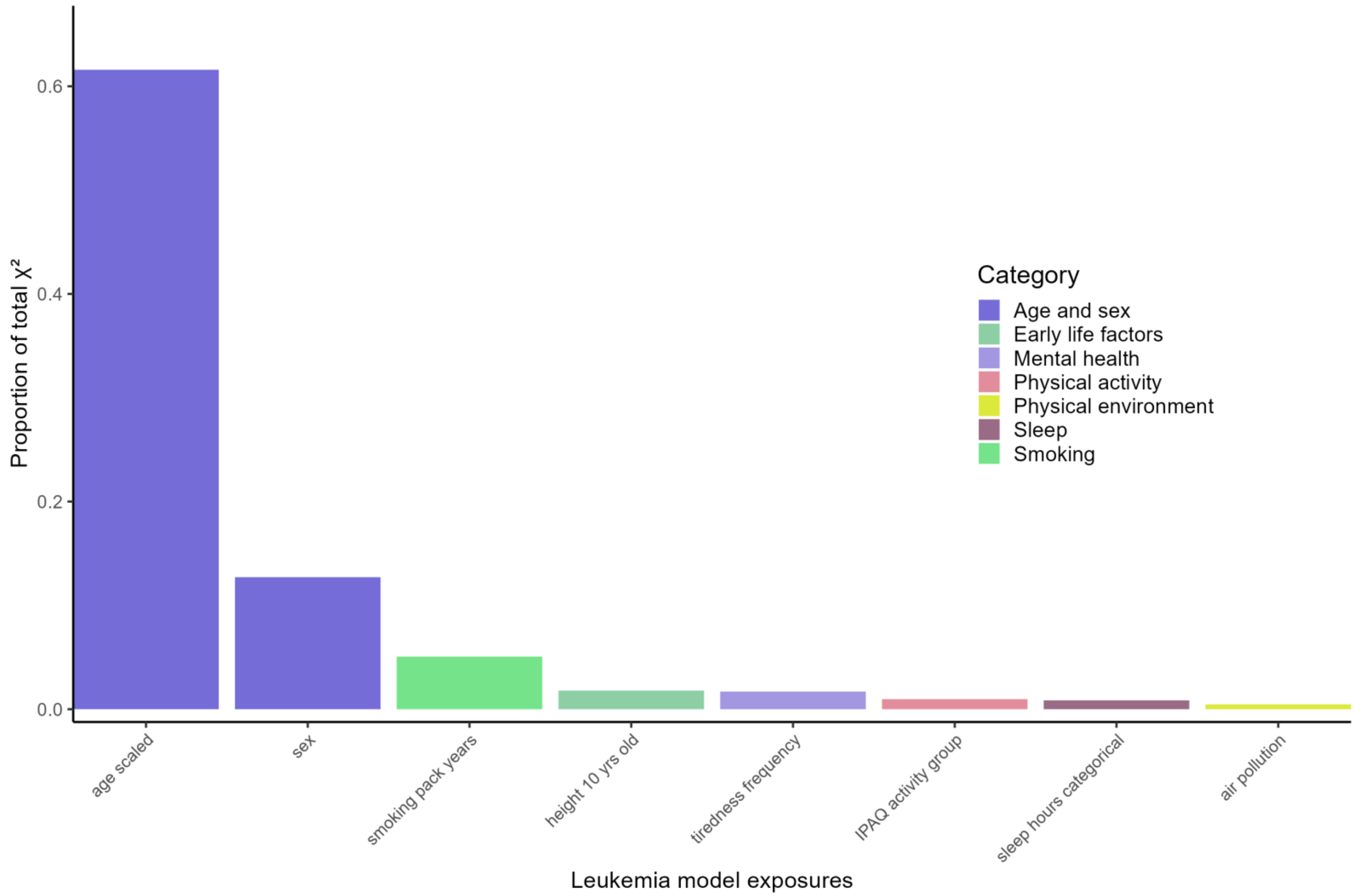

Figure S22. Variable importance according to proportion of total model chi-squared ($X^2$) for each exposure in the final multivariable leukemia exposome Cox model.

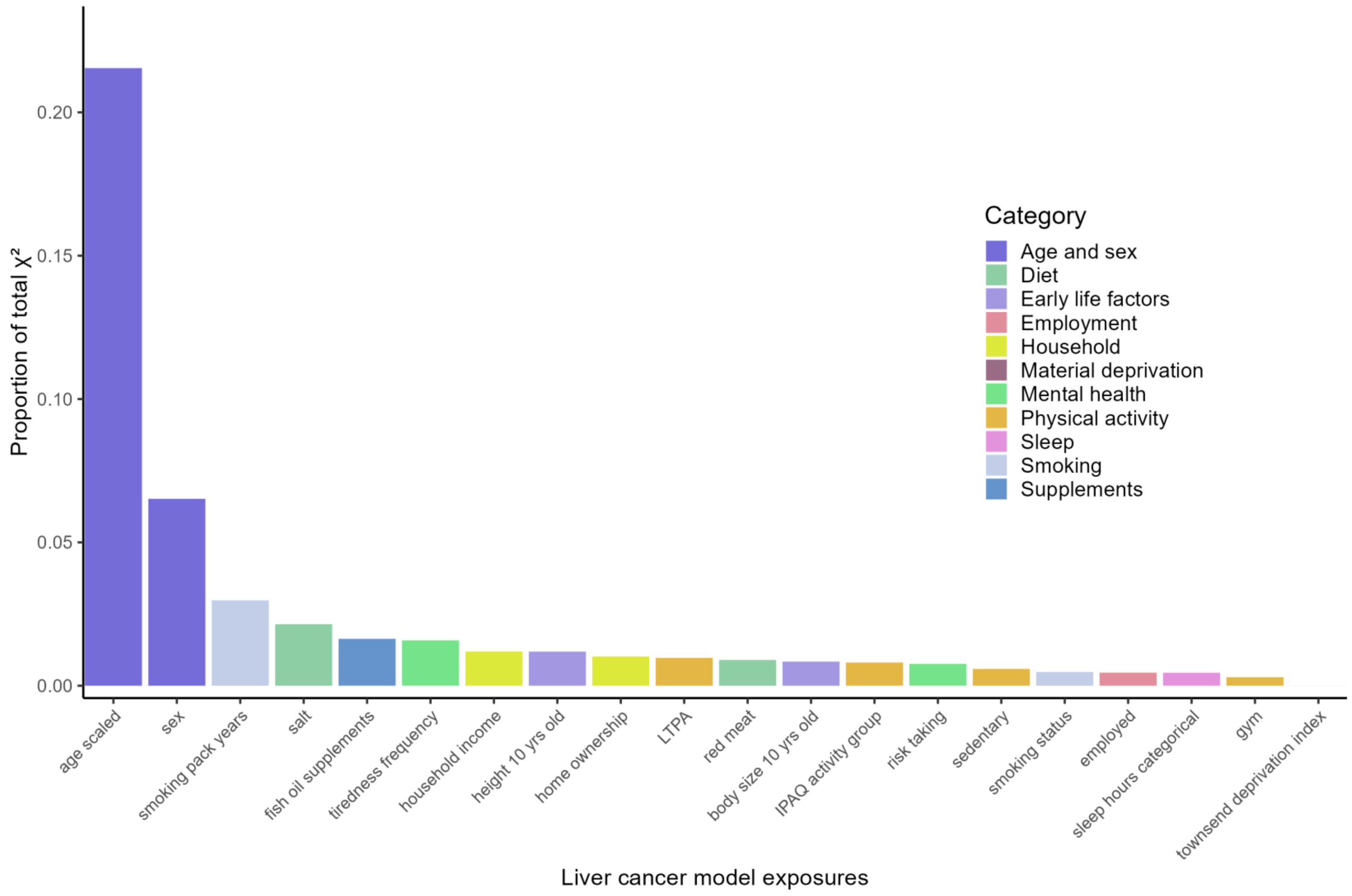

Figure S23. Variable importance according to proportion of total model chi-squared ($X^2$) for each exposure in the final multivariable liver cancer exposome Cox model.

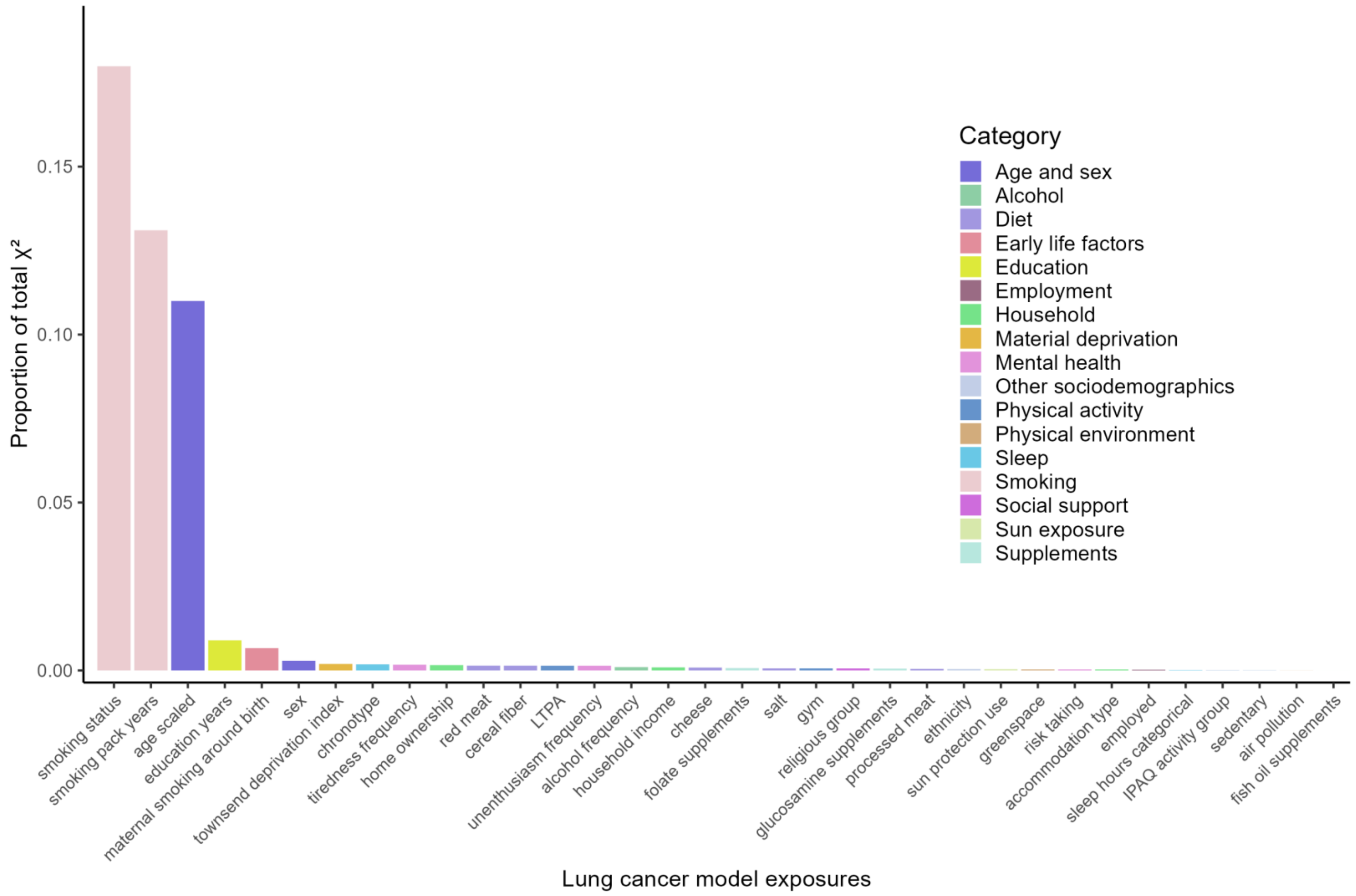

Figure S24. Variable importance according to proportion of total model chi-squared ($X^2$) for each exposure in the final multivariable lung cancer exposome Cox model.

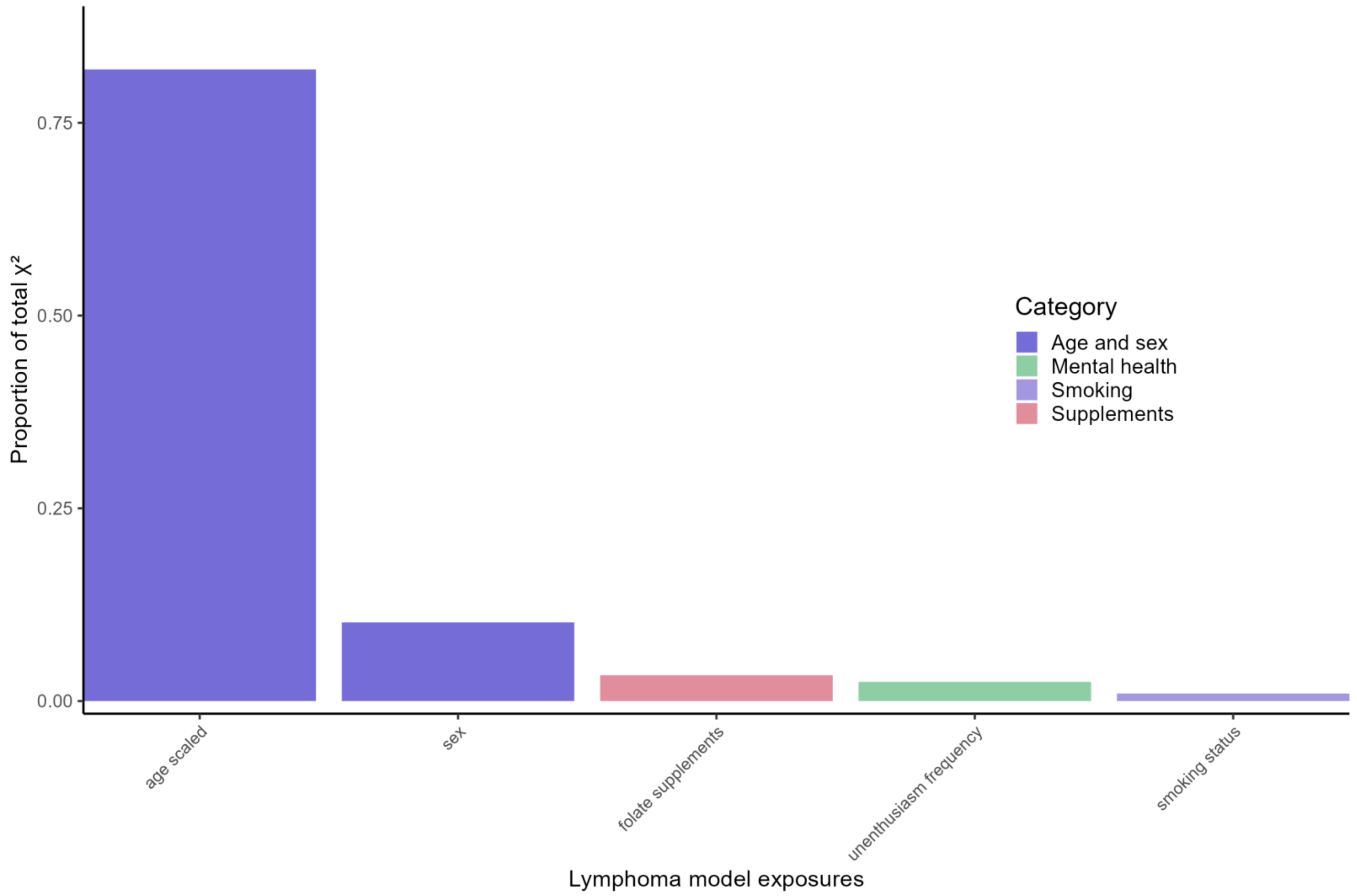

Figure S25. Variable importance according to proportion of total model chi-squared ($X^2$) for each exposure in the final multivariable lymphoma exposome Cox model.

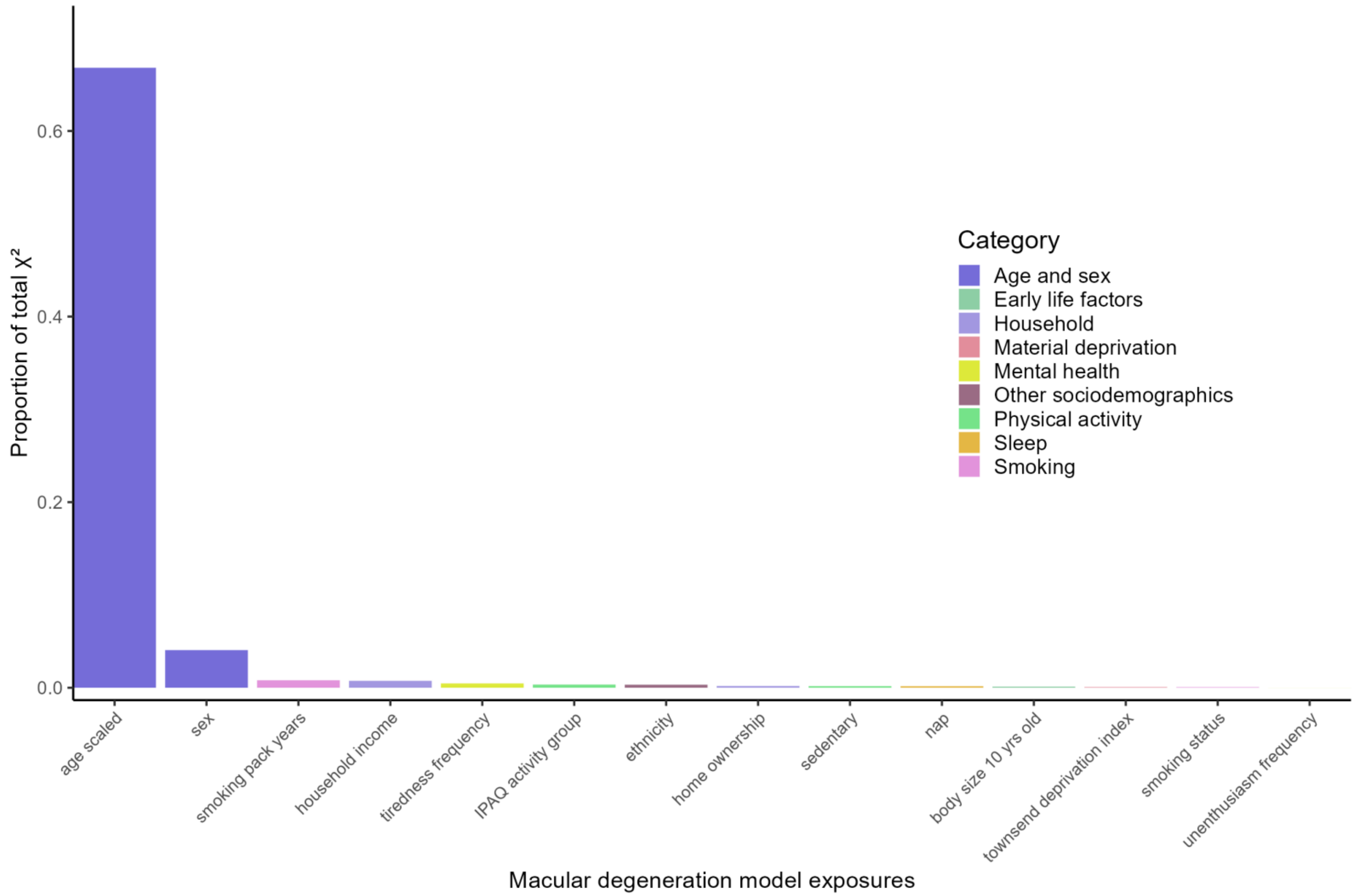

Figure S26. Variable importance according to proportion of total model chi-squared ($X^2$) for each exposure in the final multivariable macular degeneration exposome Cox model.

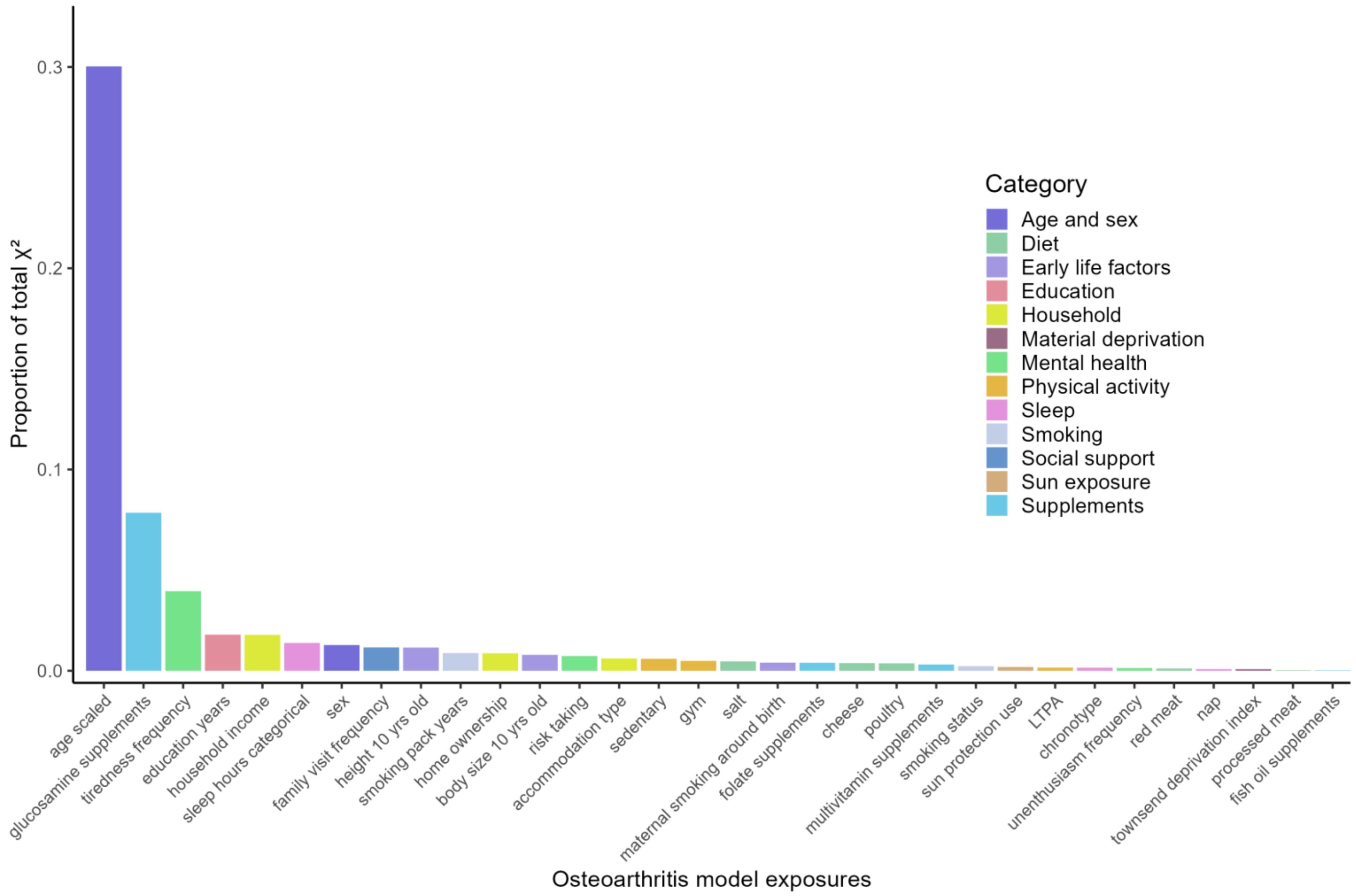

Figure S27. Variable importance according to proportion of total model chi-squared ($X^2$) for each exposure in the final multivariable osteoarthritis exposome Cox model.

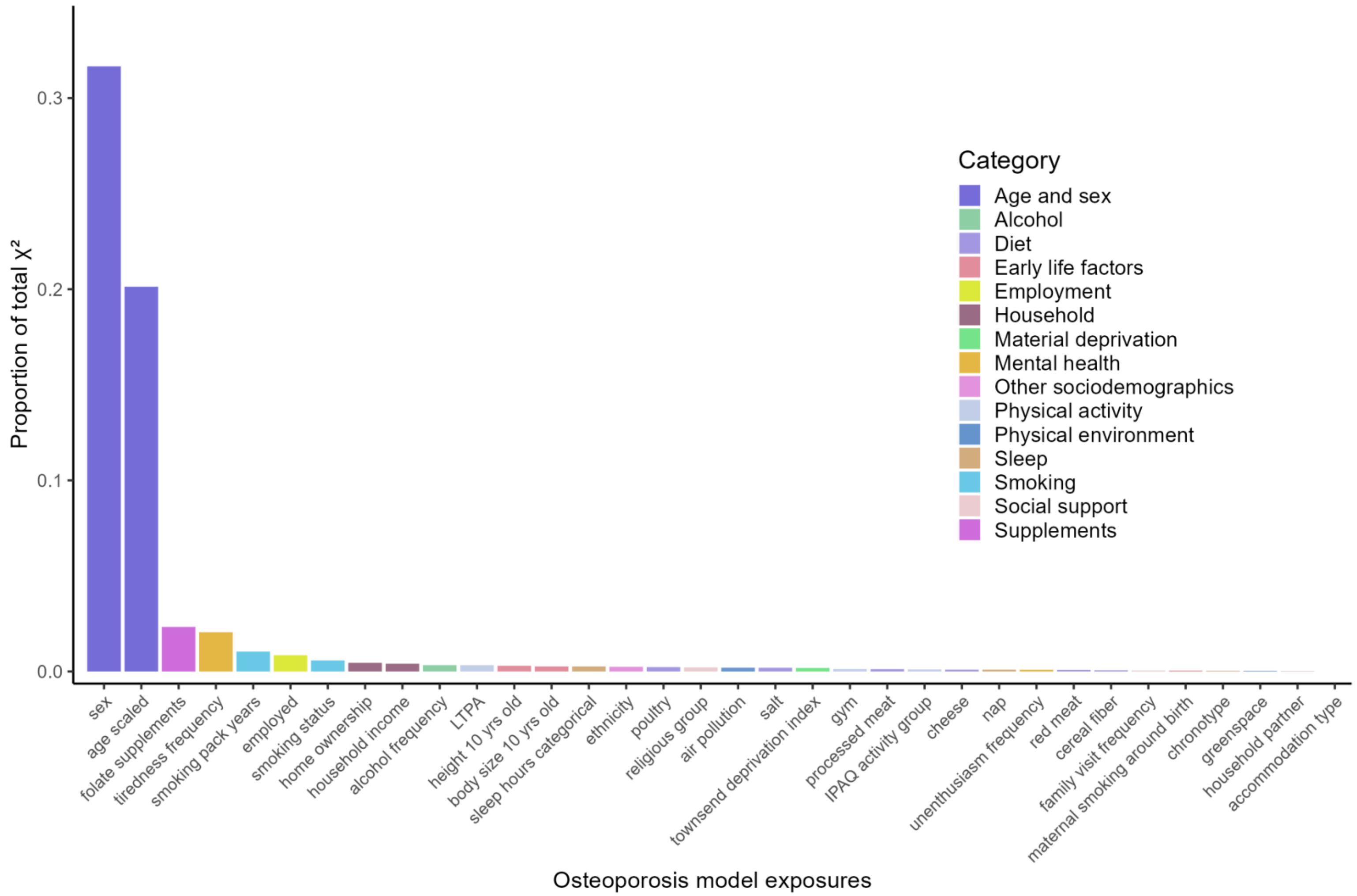

Figure S28. Variable importance according to proportion of total model chi-squared ($X^2$) for each exposure in the final multivariable osteoporosis exposome Cox model.

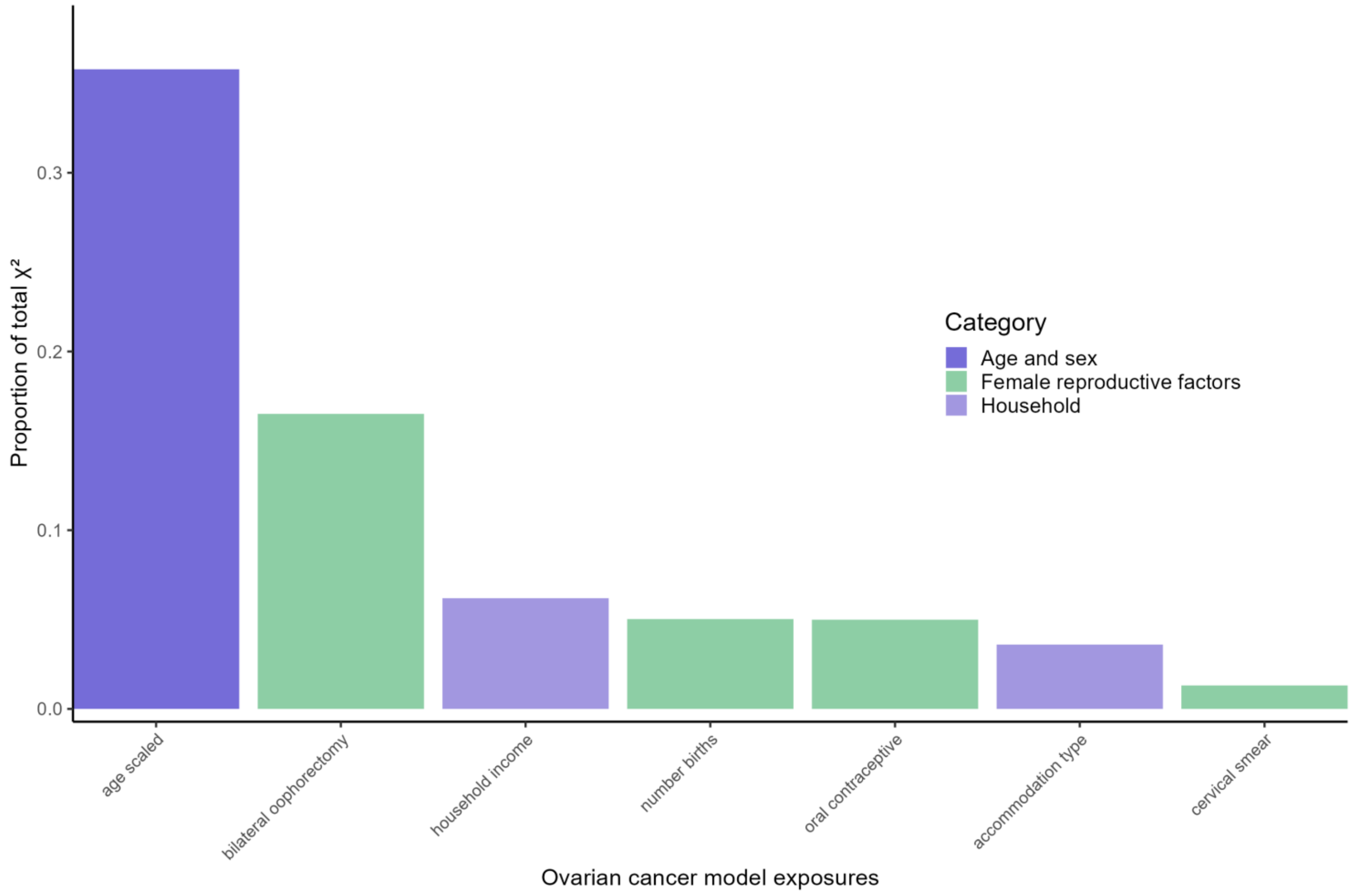

Figure S29. Variable importance according to proportion of total model chi-squared ($X^2$) for each exposure in the final multivariable ovarian cancer exposome Cox model.

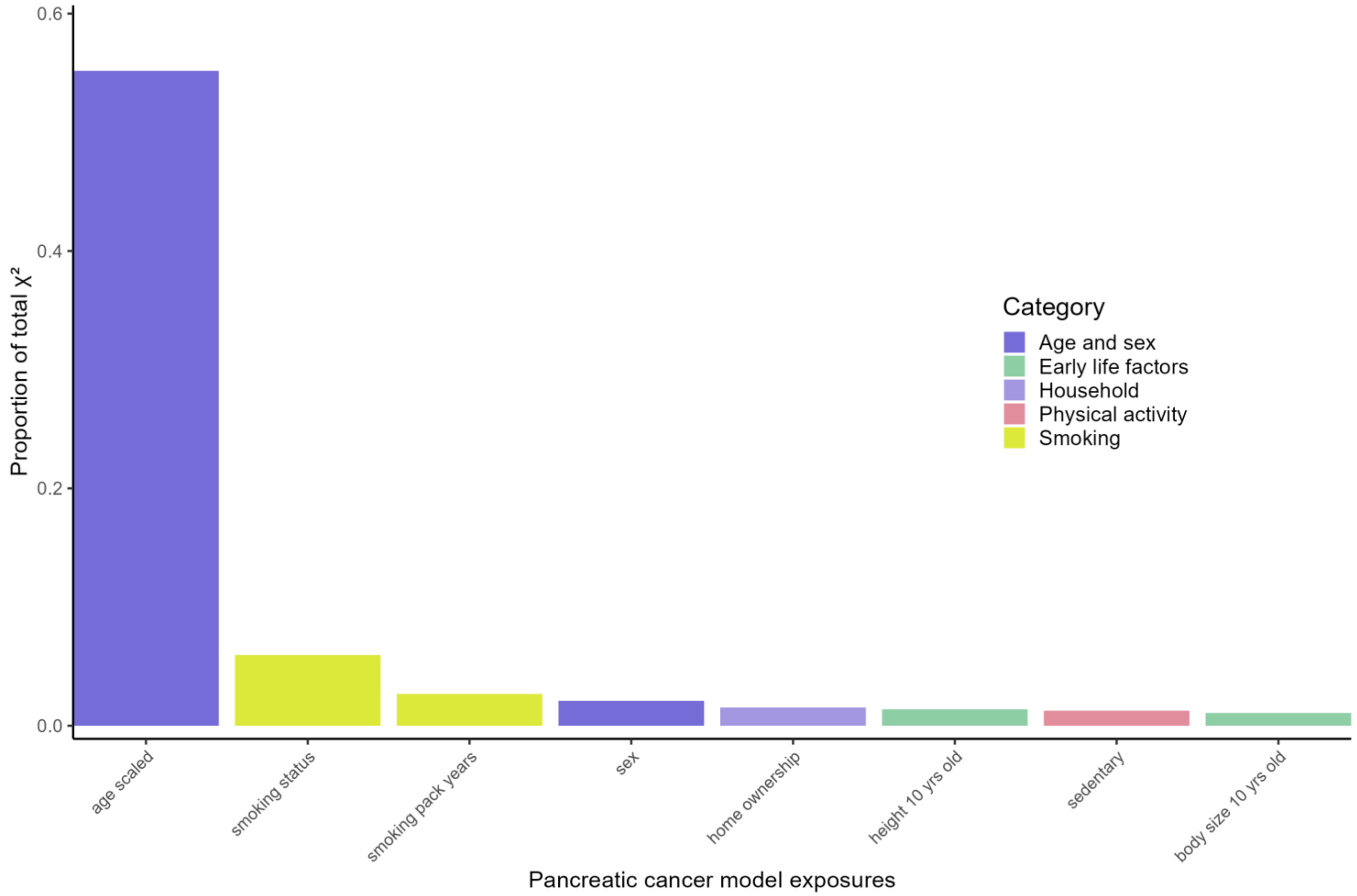

Figure S30. Variable importance according to proportion of total model chi-squared ($X^2$) for each exposure in the final multivariable pancreatic cancer exposome Cox model.

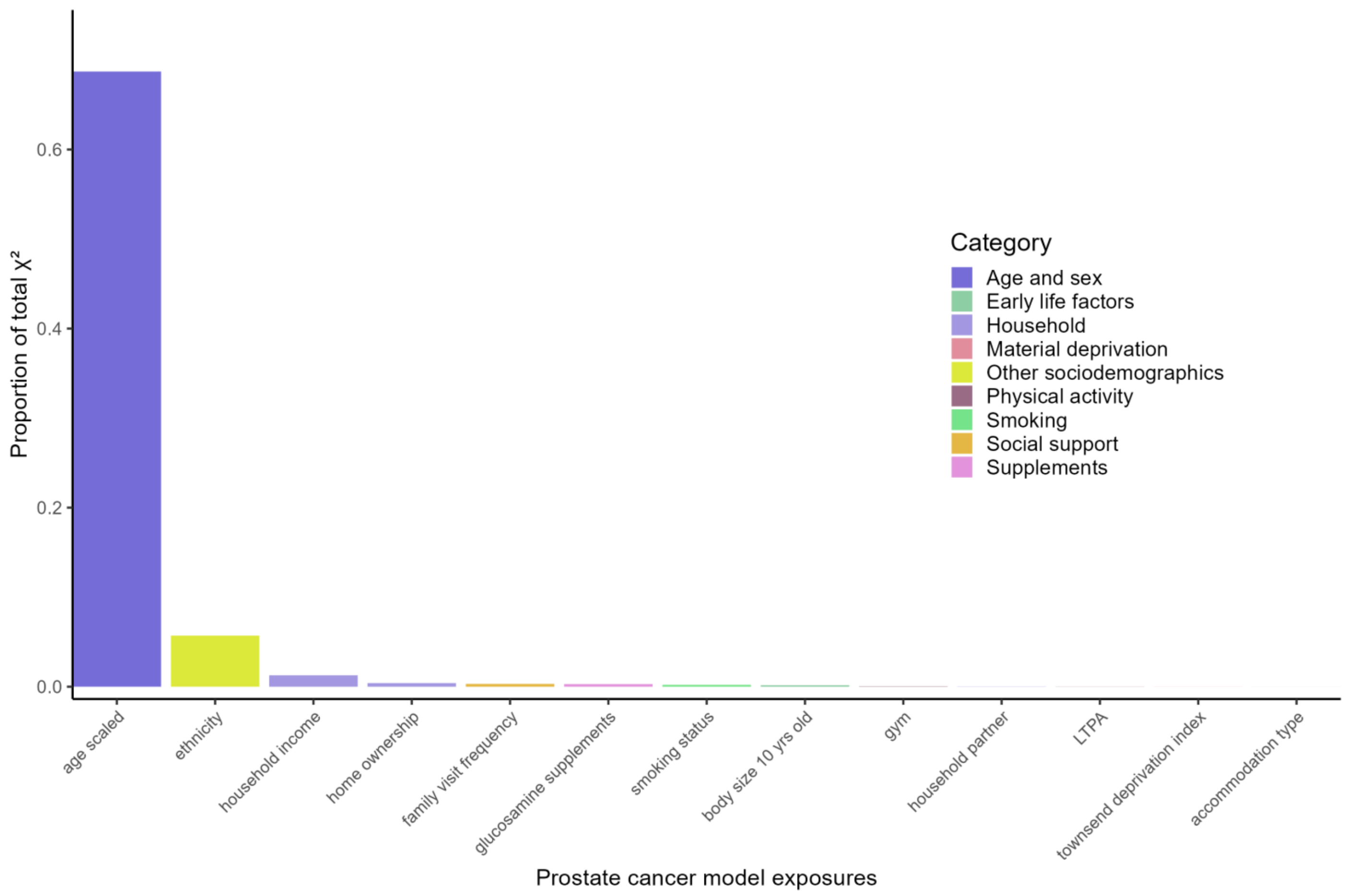

Figure S31. Variable importance according to proportion of total model chi-squared ($X^2$) for each exposure in the final multivariable prostate cancer exposome Cox model.

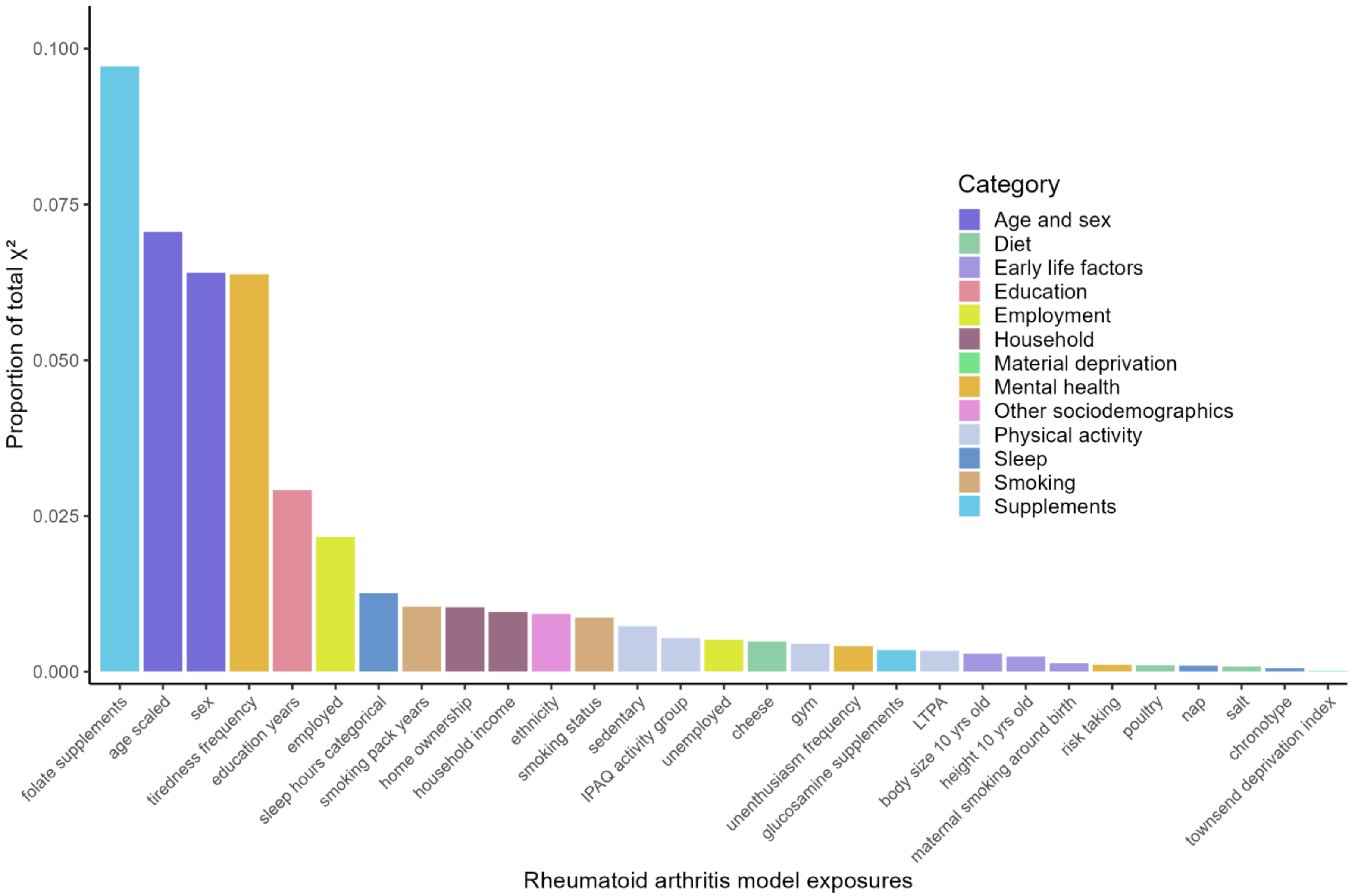

Figure S32. Variable importance according to proportion of total model chi-squared ($X^2$) for each exposure in the final multivariable rheumatoid arthritis exposome Cox model.

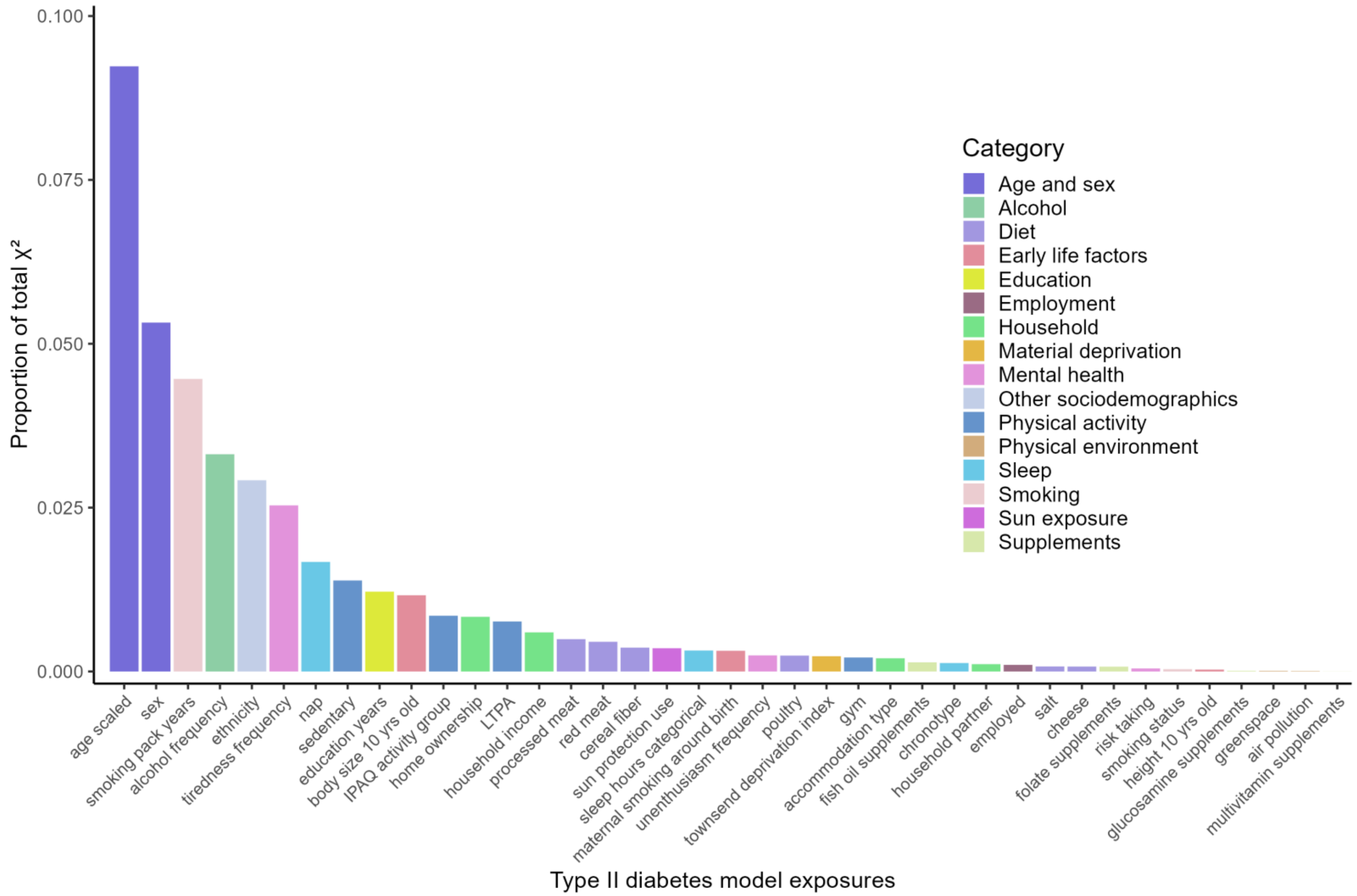

Figure S33. Variable importance according to proportion of total model chi-squared ($X^2$) for each exposure in the final multivariable type II diabetes exposome Cox model.

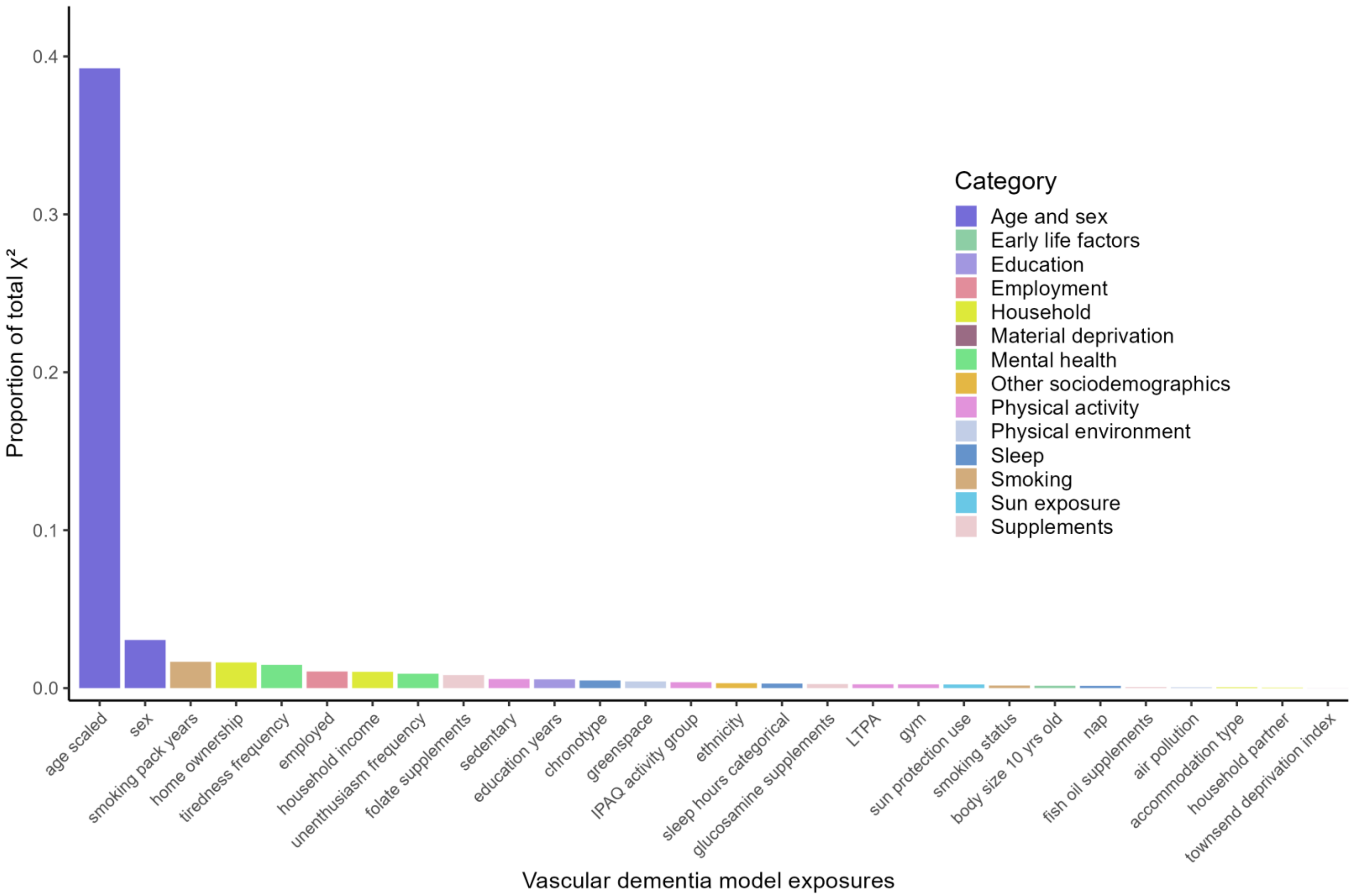

Figure S34. Variable importance according to proportion of total model chi-squared ($X^2$) for each exposure in the final multivariable vascular dementia exposome Cox model.

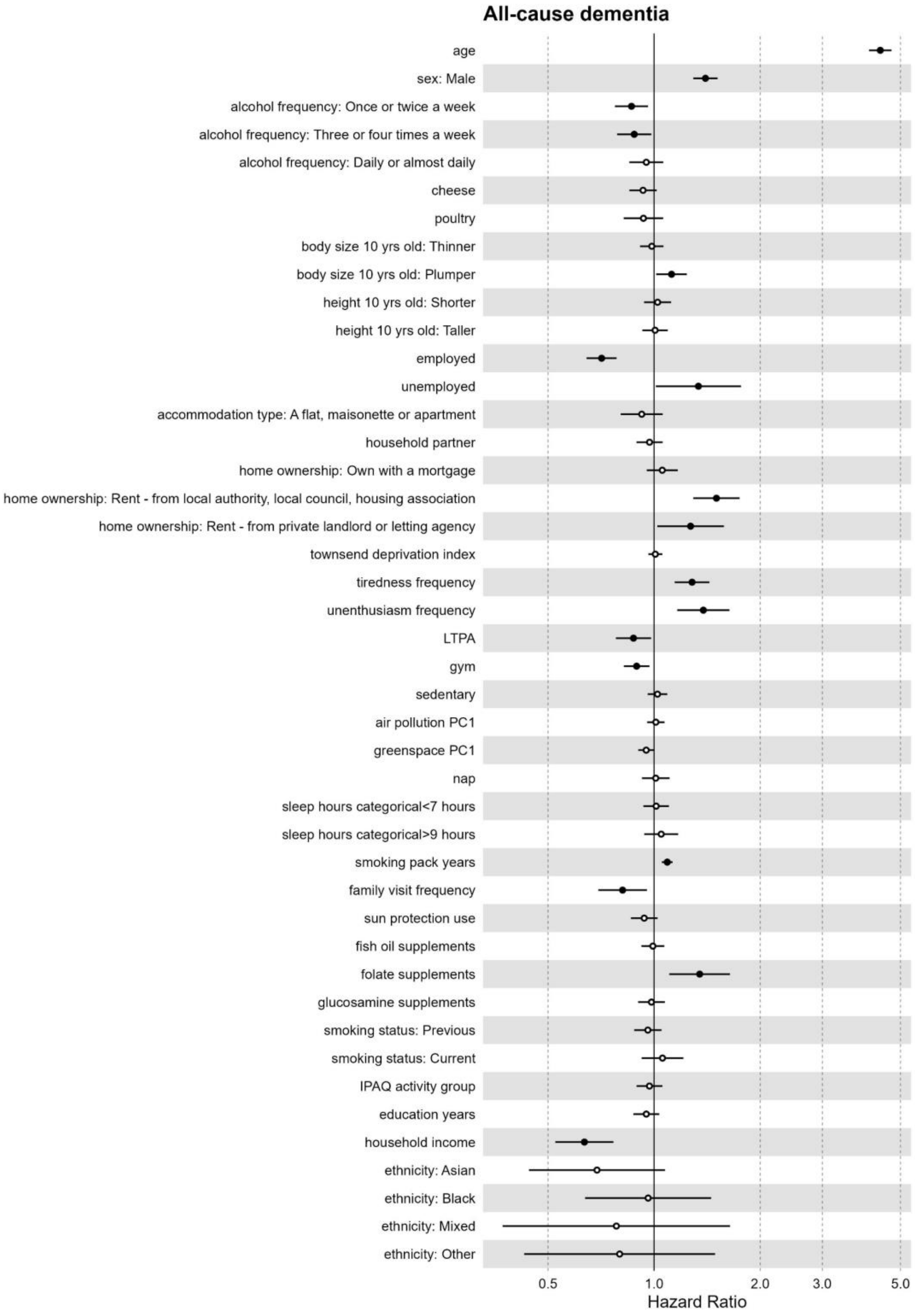

Figure S35. Effect estimates from the full exposome multivariable Cox model for All-cause dementia in UK Biobank Participants recruited in England (n=436,891).

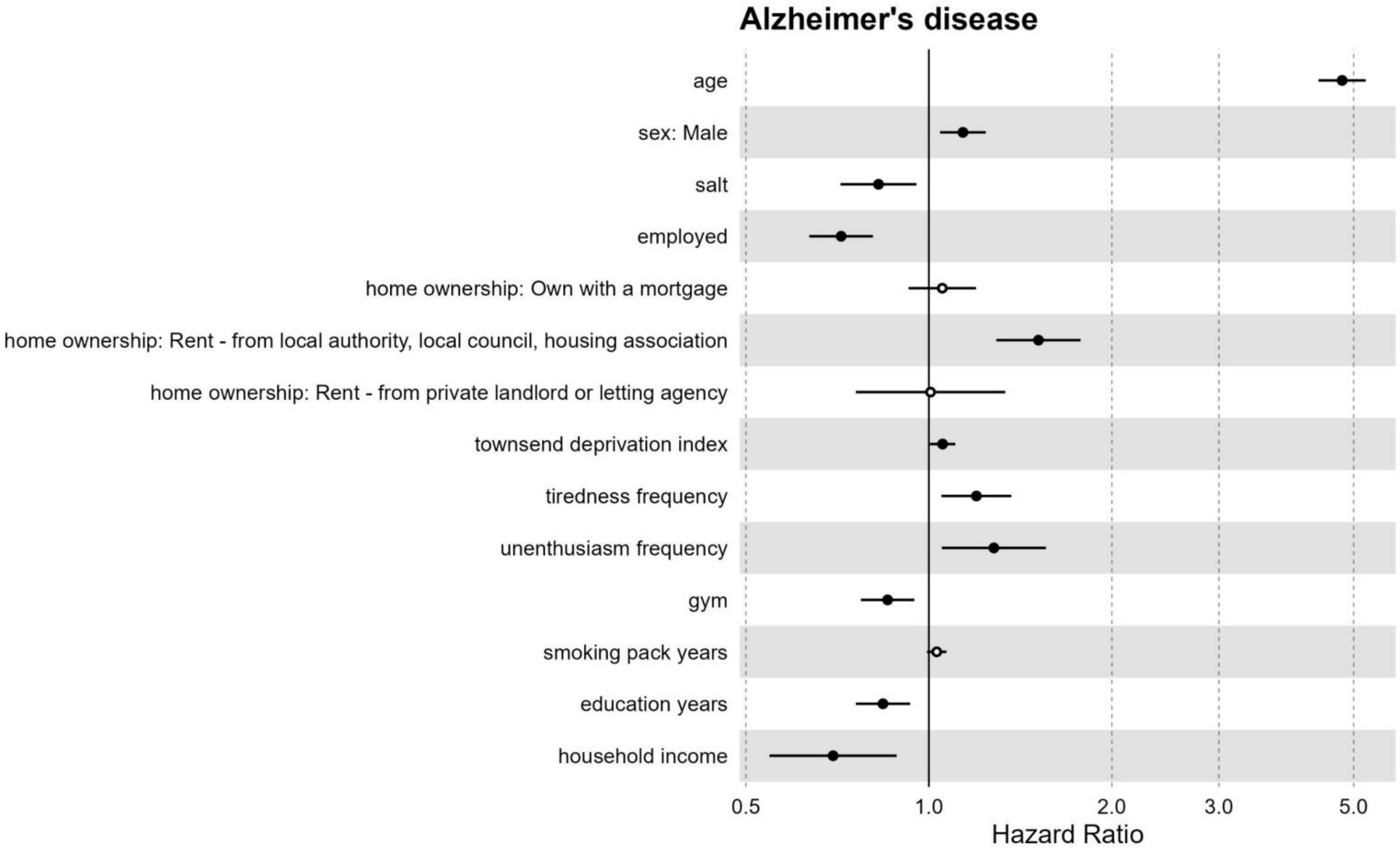

Figure S36. Effect estimates from the full exposome multivariable Cox model for Alzheimer’s disease in UK Biobank Participants recruited in England (n=436,891).

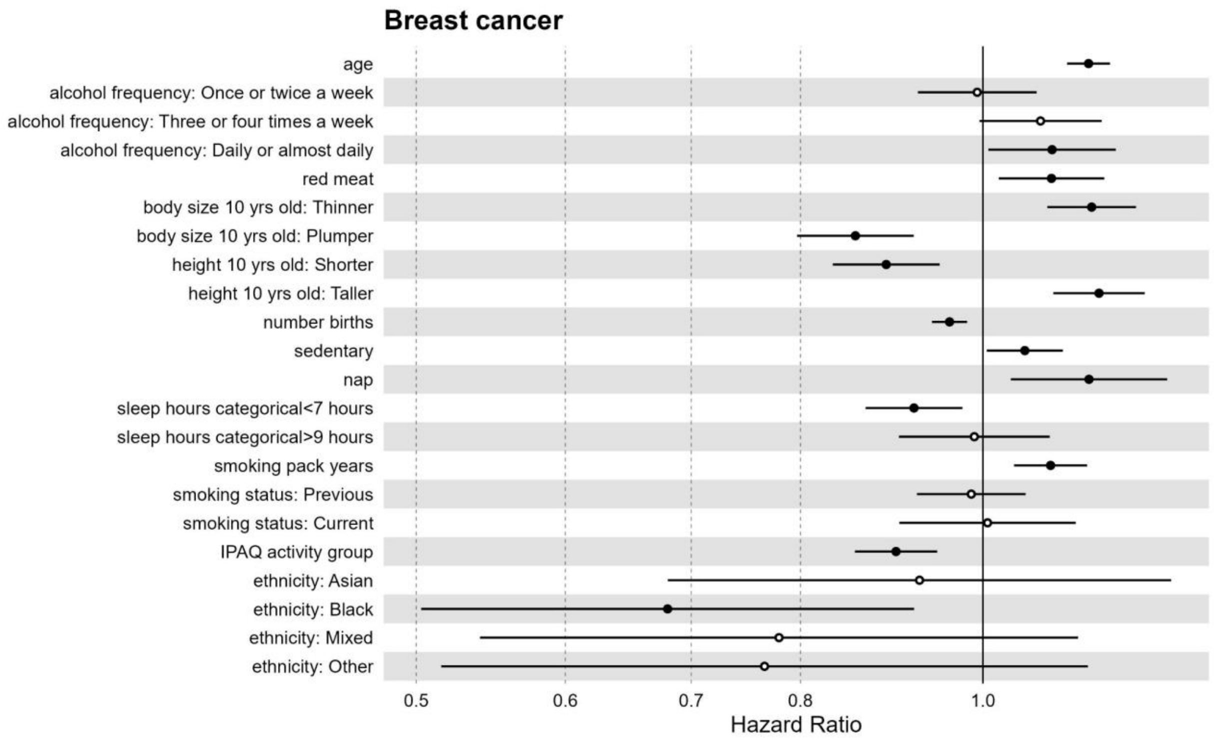

Figure S37. Effect estimates from the full exposome multivariable Cox model for breast cancer in UK Biobank women recruited in England (n=237,634).

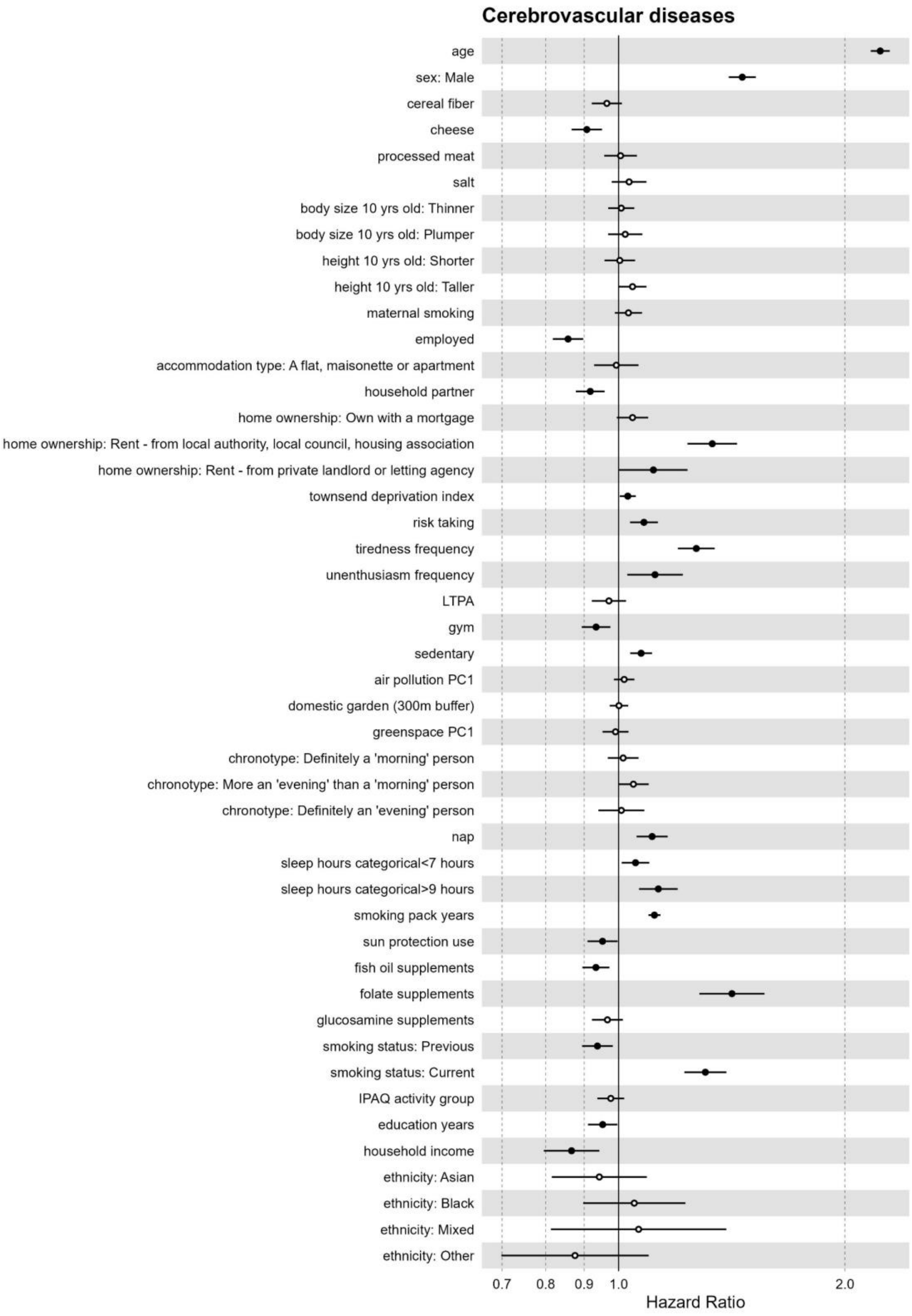

Figure S38. Effect estimates from the full exposome multivariable Cox model for cerebrovascular diseases in UK Biobank Participants recruited in England (n=436,891).

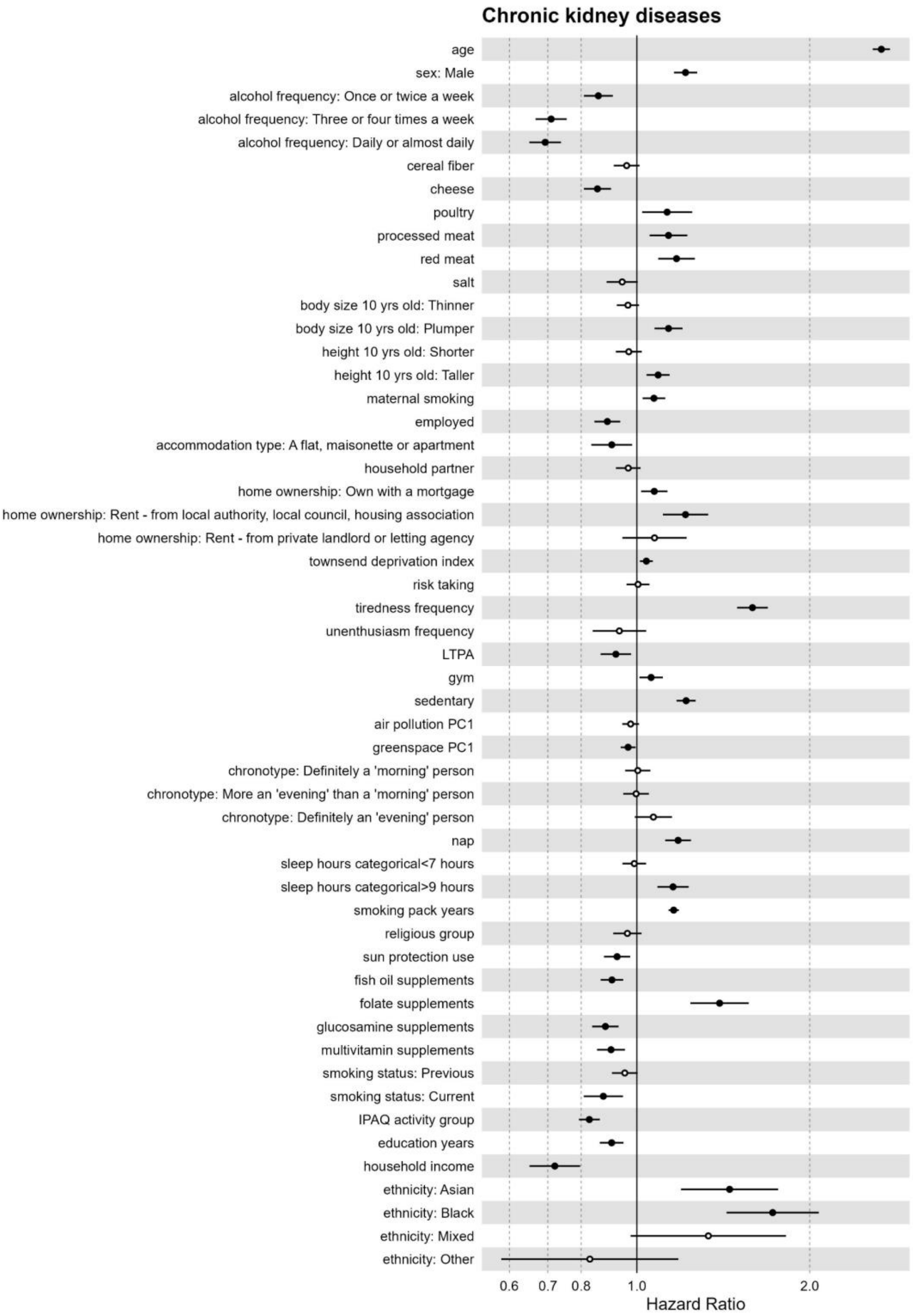

Figure S39. Effect estimates from the full exposome multivariable Cox model for chronic kidney diseases in UK Biobank Participants recruited in England (n=436,891).

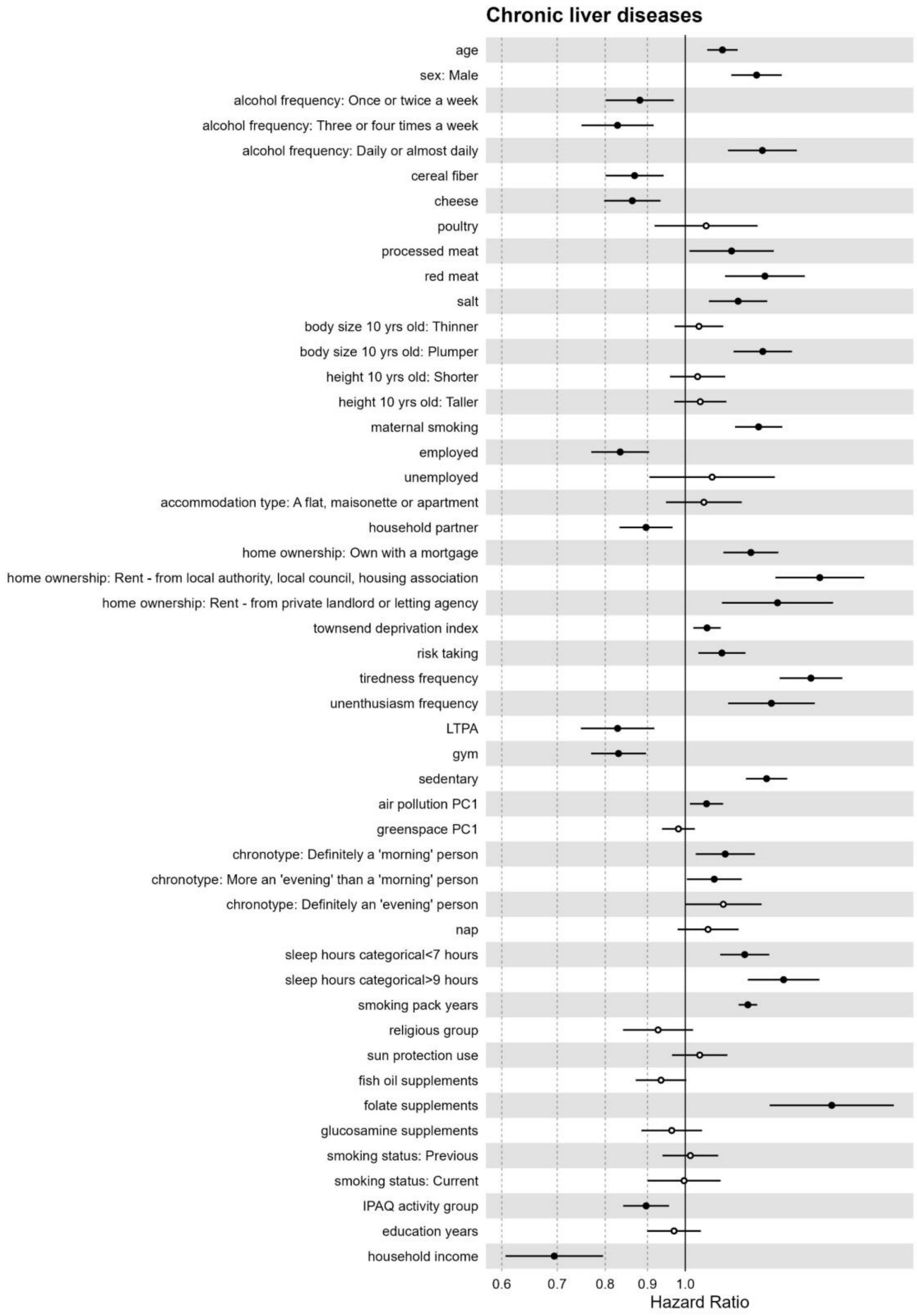

Figure S40. Effect estimates from the full exposome multivariable Cox model for chronic liver diseases in UK Biobank Participants recruited in England (n=436,891).

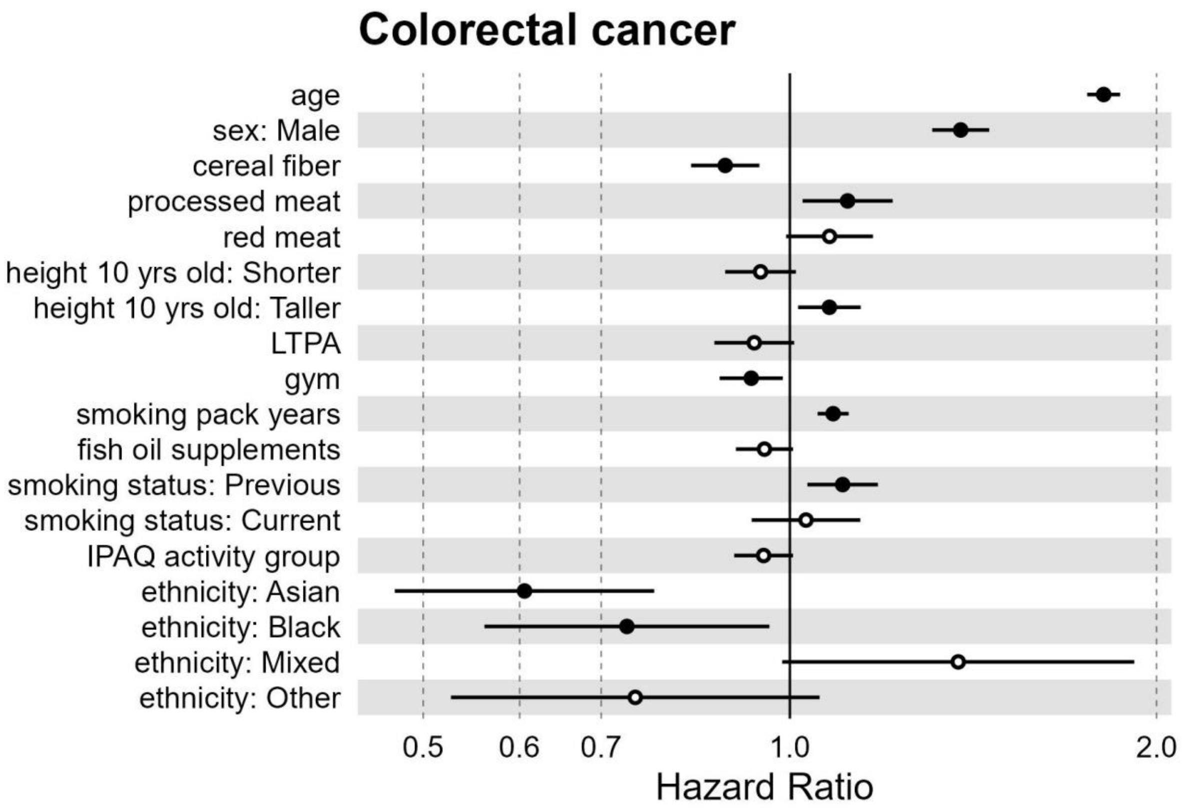

Figure S41. Effect estimates from the full exposome multivariable Cox model for colorectal cancer in UK Biobank Participants recruited in England (n=436,891).

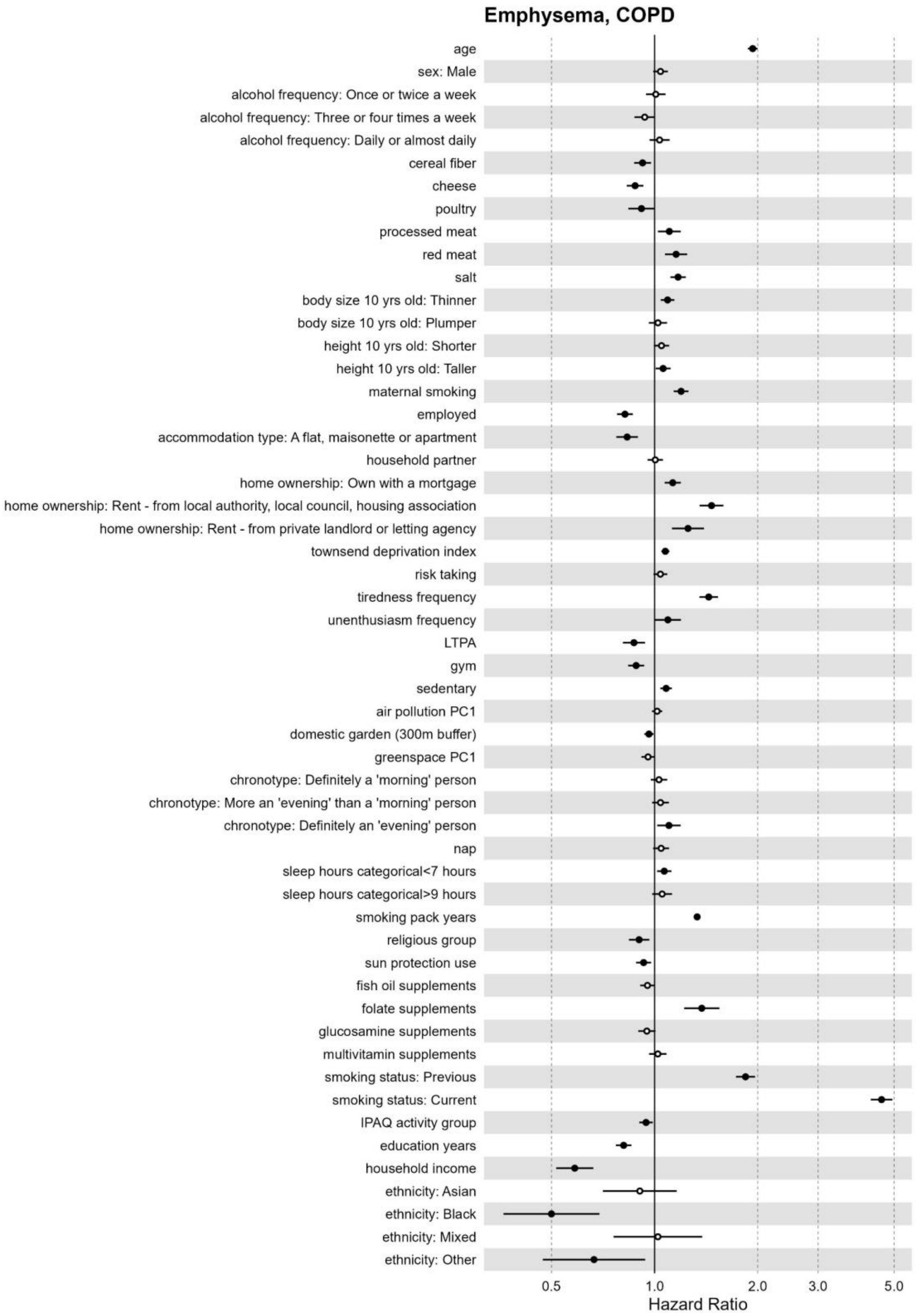

Figure S42. Effect estimates from the full exposome multivariable Cox model for emphysema, COPD in UK Biobank Participants recruited in England (n=436,891).

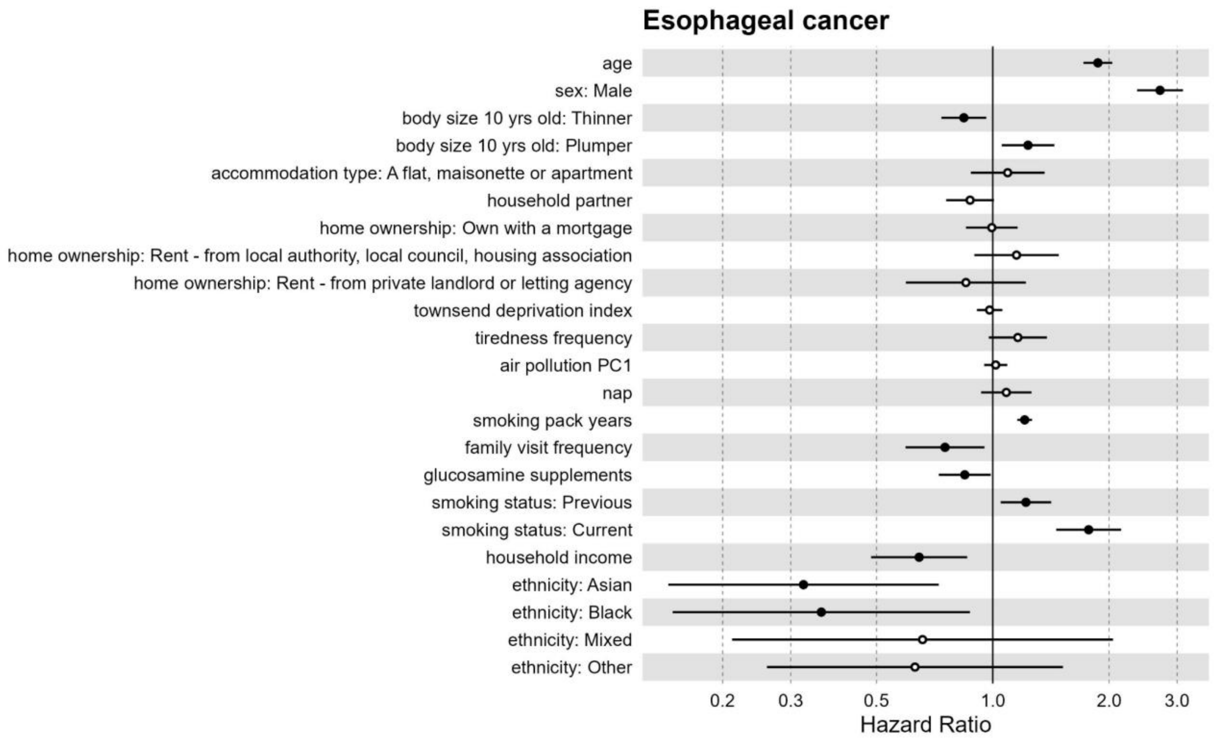

Figure S43. Effect estimates from the full exposome multivariable Cox model for esophageal cancer in UK Biobank Participants recruited in England (n=436,891).

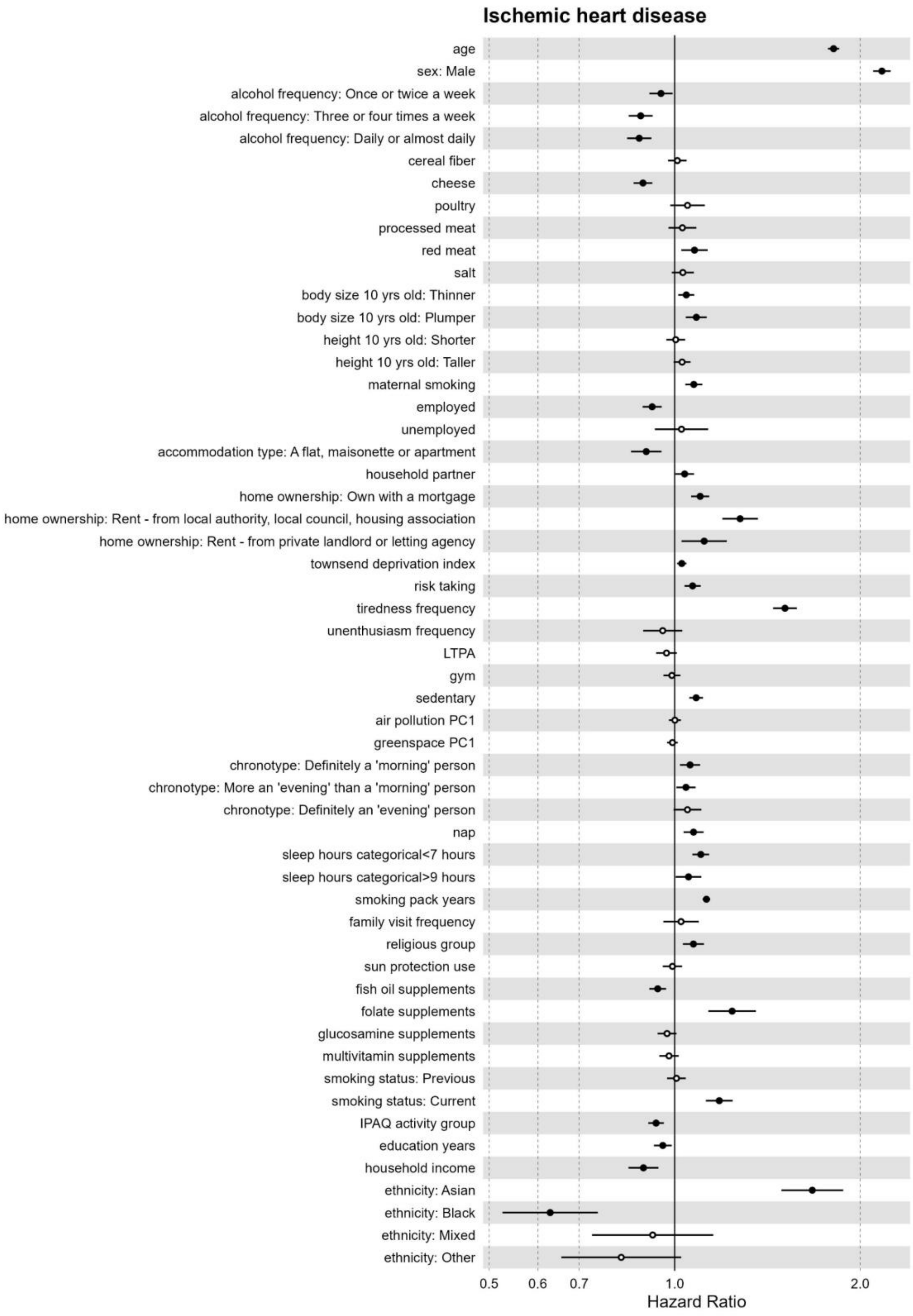

Figure S44. Effect estimates from the full exposome multivariable Cox model for ischemic heart disease in UK Biobank Participants recruited in England (n=436,891).

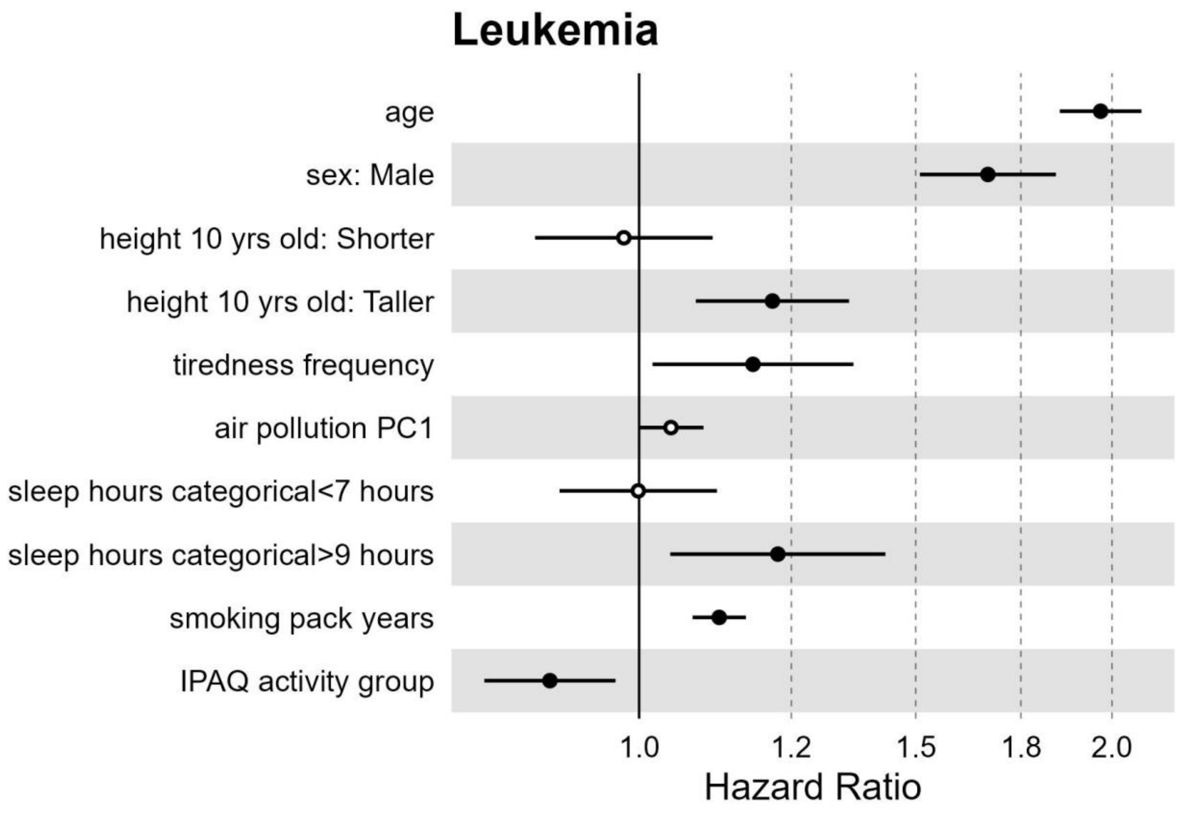

Figure S45. Effect estimates from the full exposome multivariable Cox model for leukemia in UK Biobank Participants recruited in England (n=436,891).

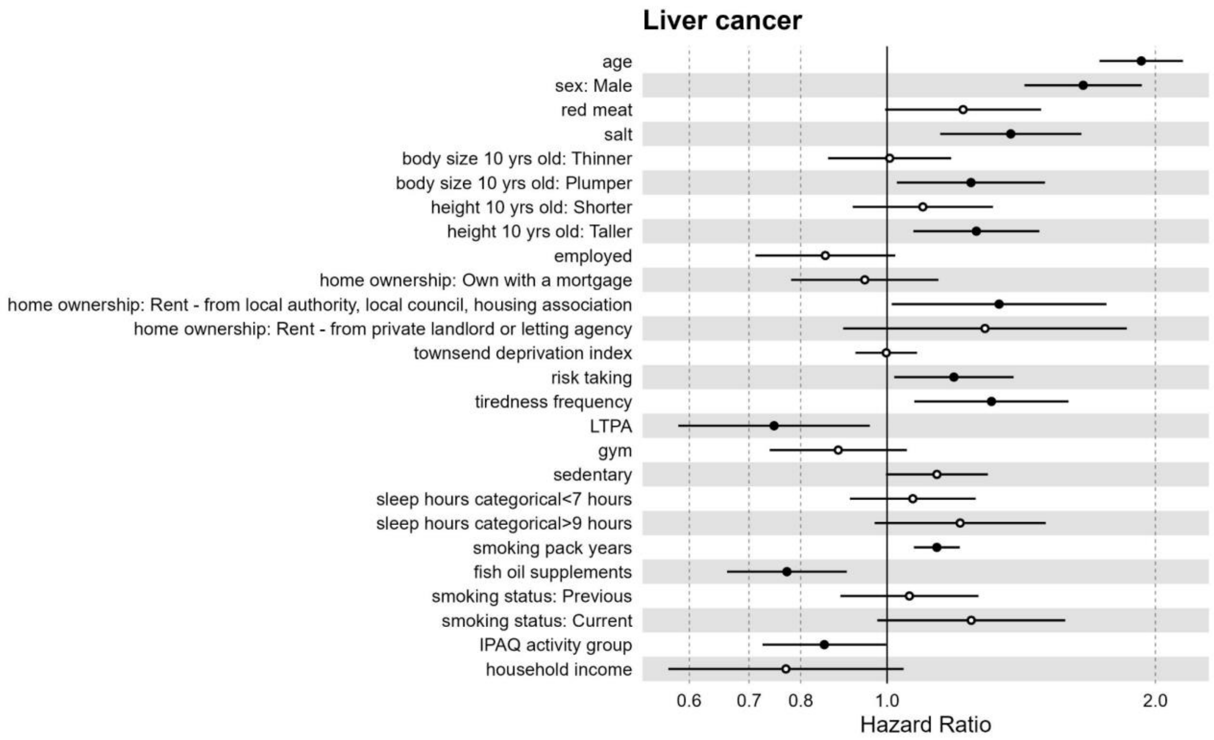

Figure S46. Effect estimates from the full exposome multivariable Cox model for liver cancer in UK Biobank Participants recruited in England (n=436,891).

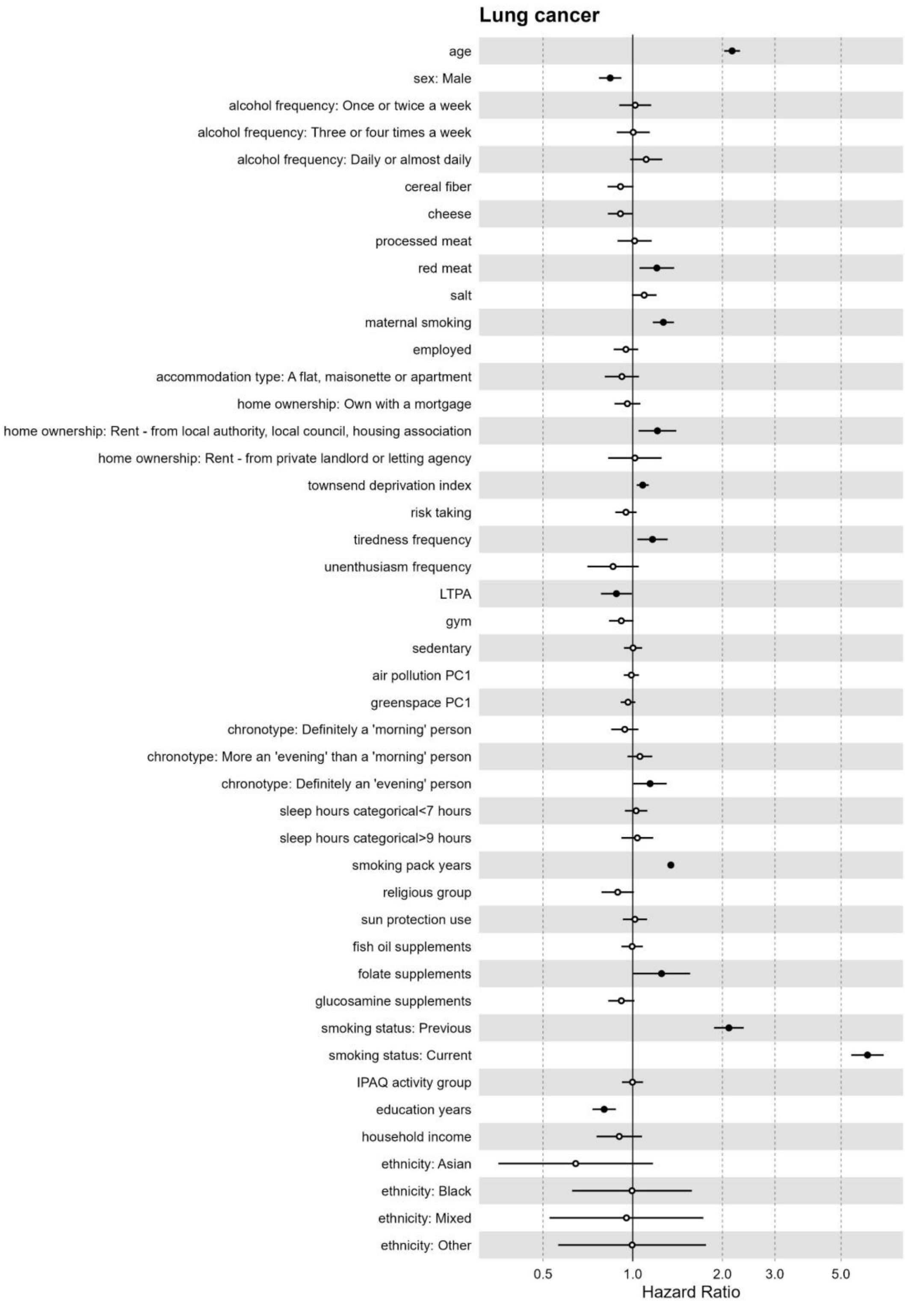

Figure S47. Effect estimates from the full exposome multivariable Cox model for lung cancer in UK Biobank Participants recruited in England (n=436,891).

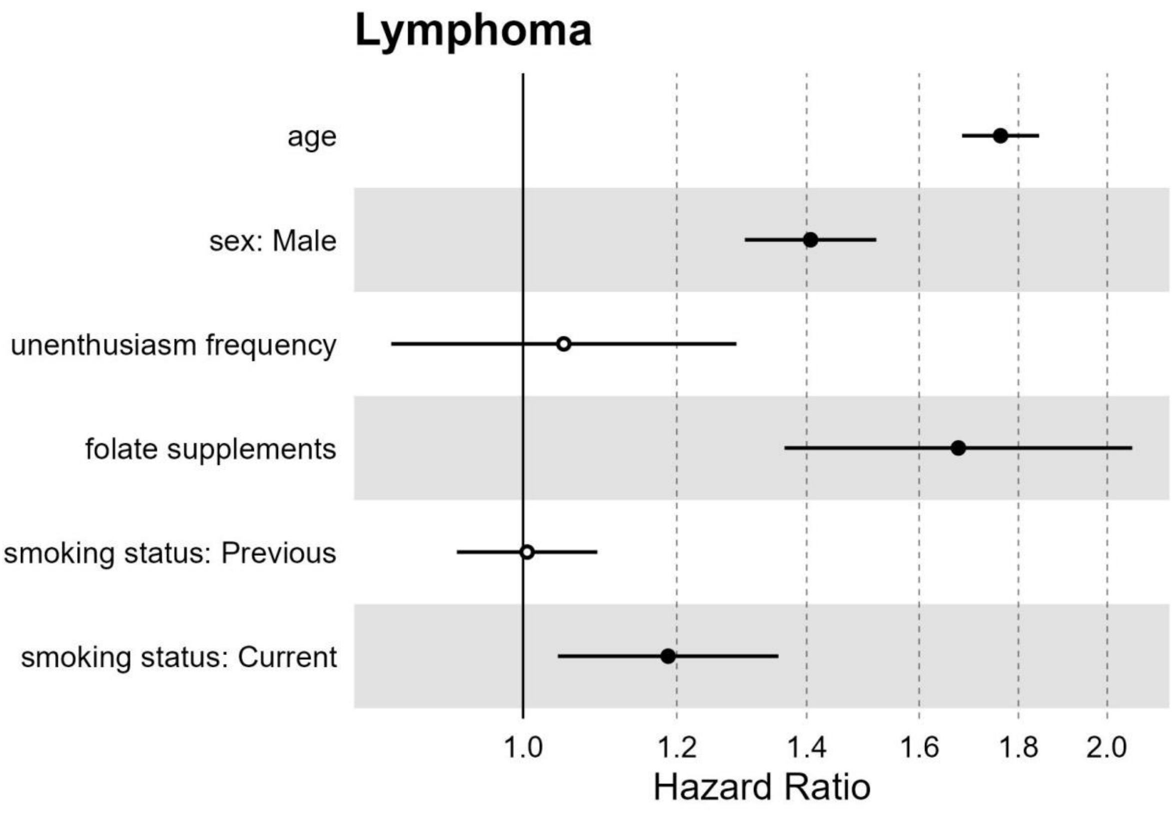

Figure S48. Effect estimates from the full exposome multivariable Cox model for lymphoma in UK Biobank Participants recruited in England (n=436,891).

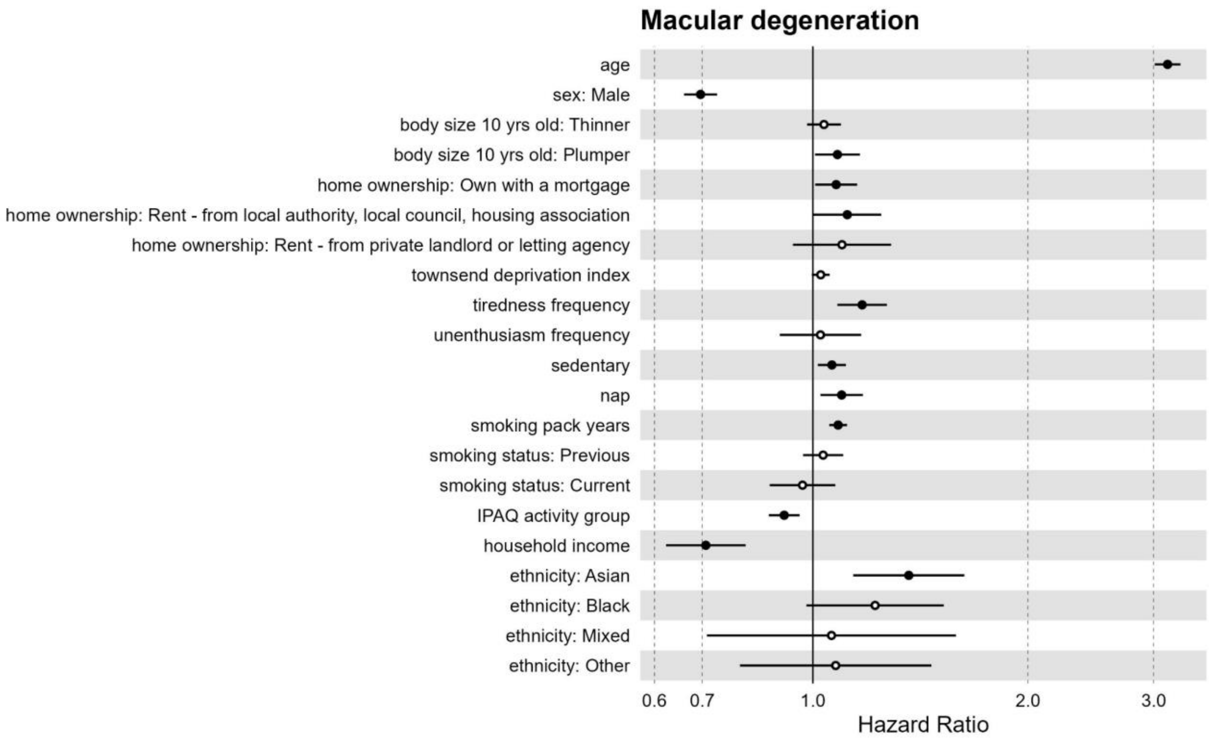

Figure S49. Effect estimates from the full exposome multivariable Cox model for macular degeneration in UK Biobank Participants recruited in England (n=436,891).

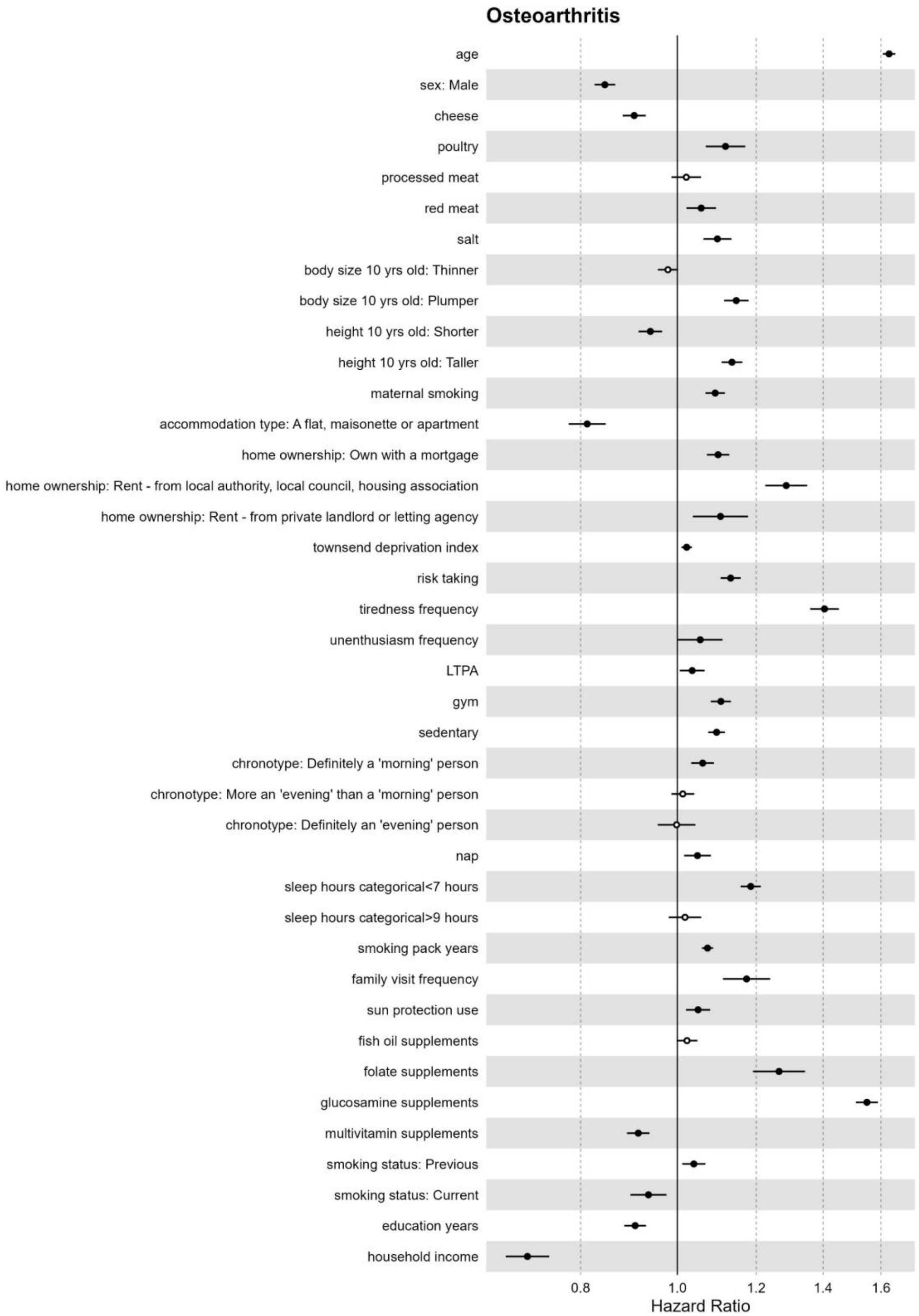

Figure S50. Effect estimates from the full exposome multivariable Cox model for osteoarthritis in UK Biobank Participants recruited in England (n=436,891).

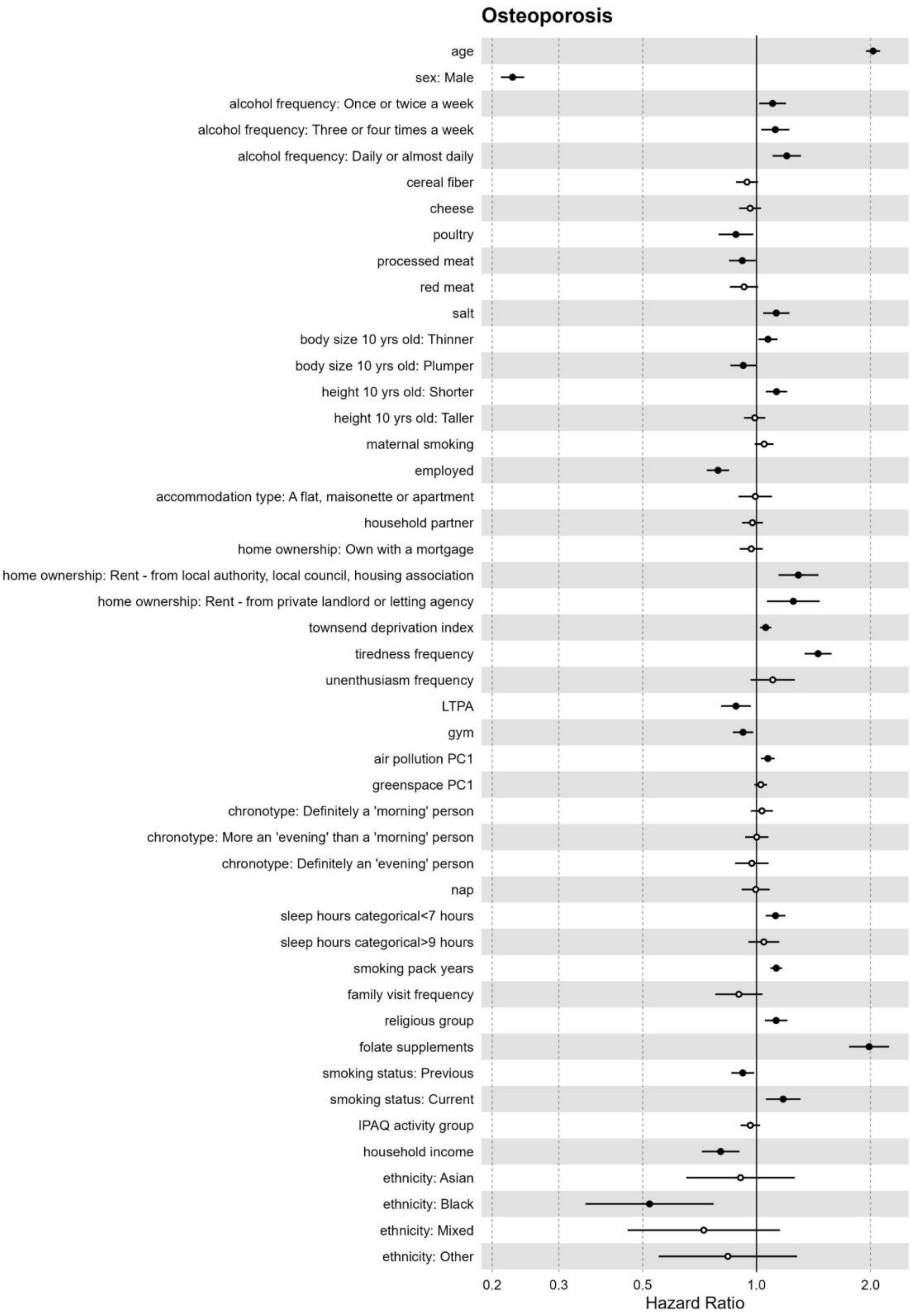

Figure S51. Effect estimates from the full exposome multivariable Cox model for osteoporosis in UK Biobank Participants recruited in England (n=436,891).

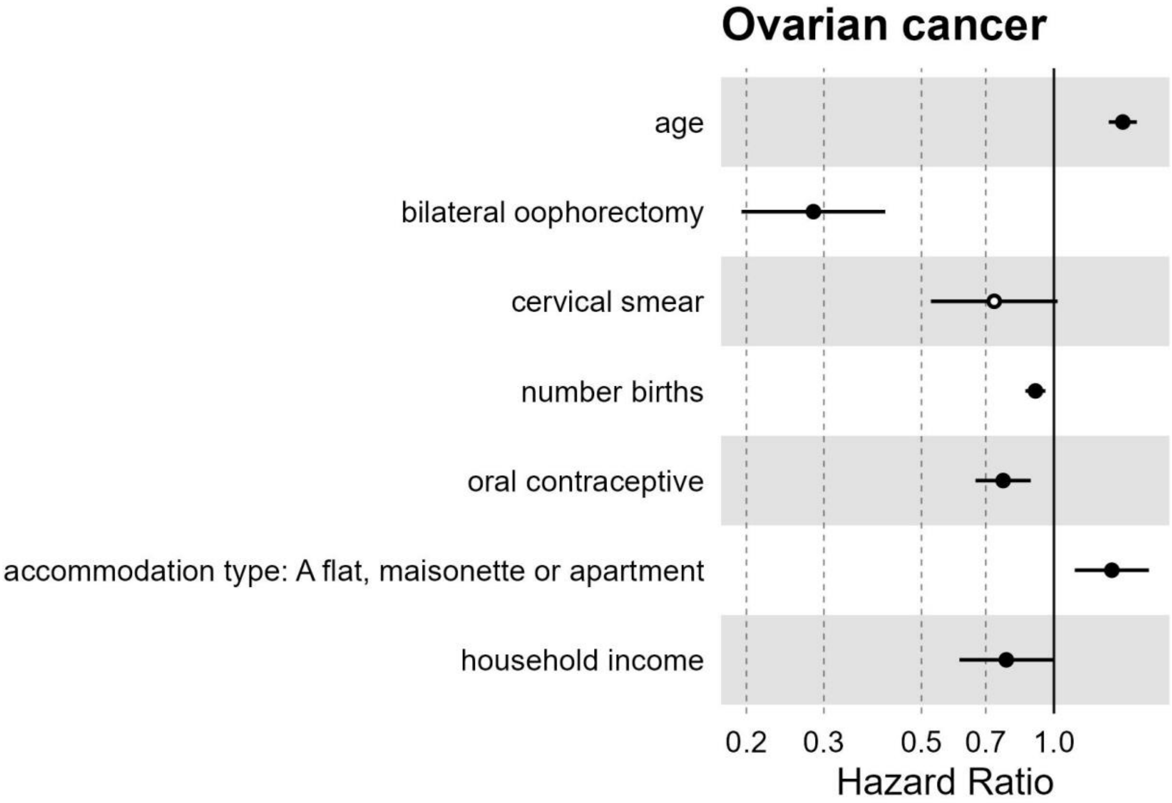

Figure S52. Effect estimates from the full exposome multivariable Cox model for ovarian cancer in UK Biobank women recruited in England (n=237,634).

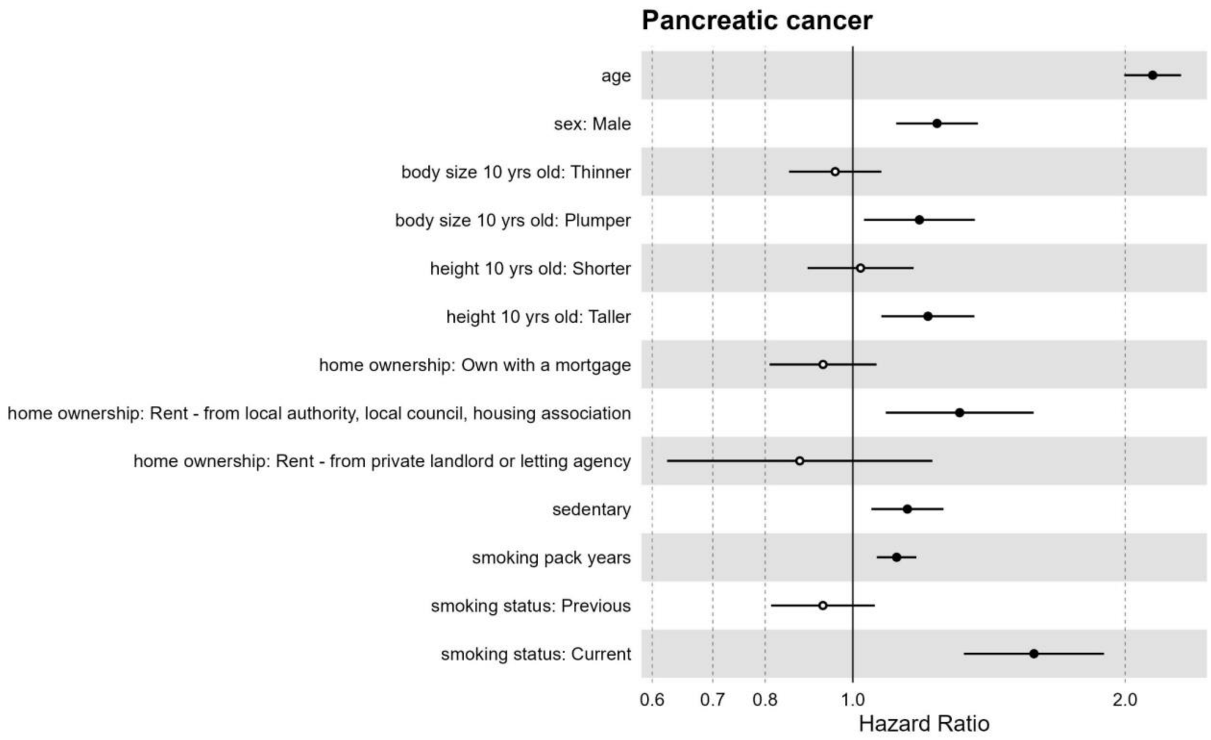

Figure S53. Effect estimates from the full exposome multivariable Cox model for pancreatic cancer in UK Biobank Participants recruited in England (n=436,891). Model also included covariates for age and sex.

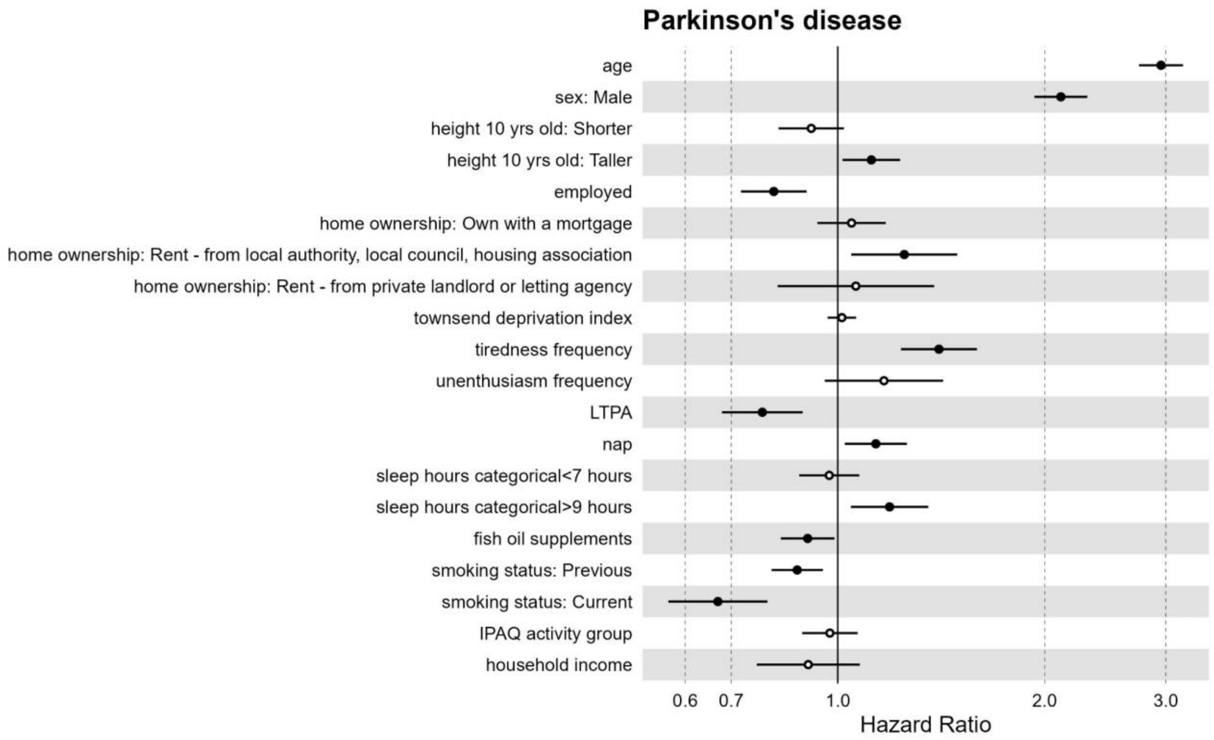

Figure S54. Effect estimates from the full exposome multivariable Cox model for Parkinson's disease in UK Biobank Participants recruited in England (n=436,891).

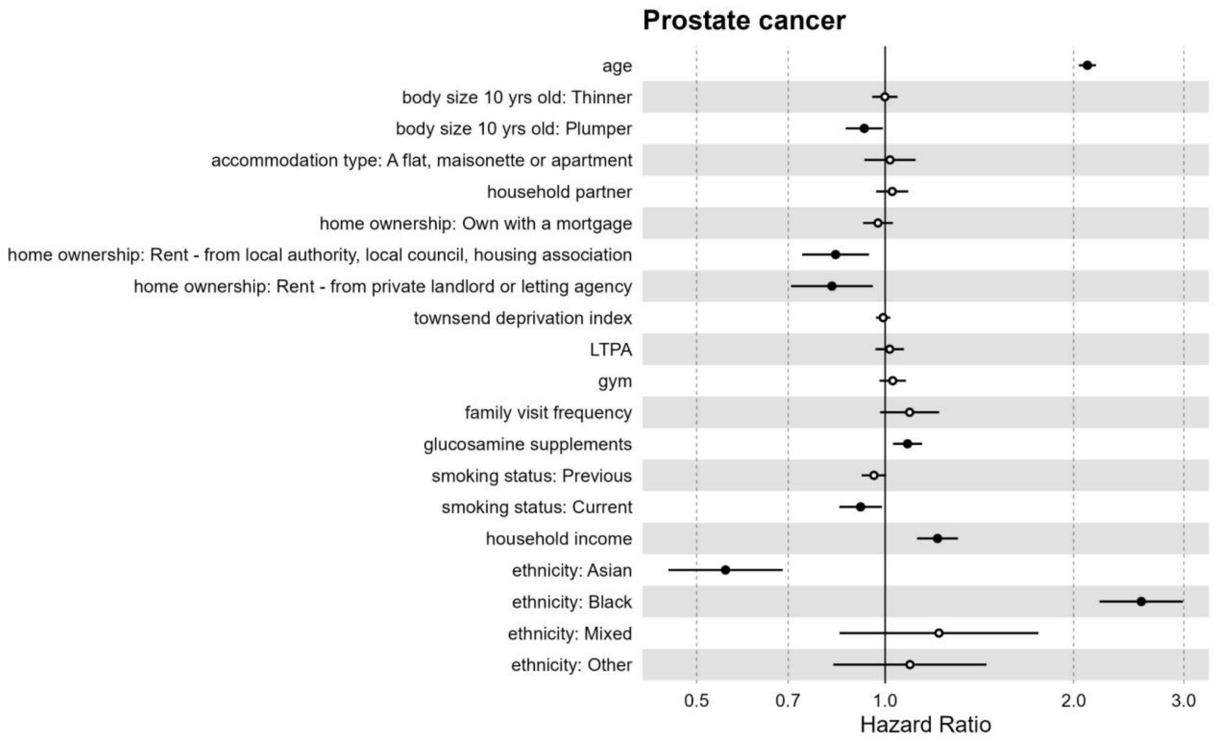

Figure S55. Effect estimates from the full exposome multivariable Cox model for prostate cancer in UK Biobank men recruited in England (n=199,257).

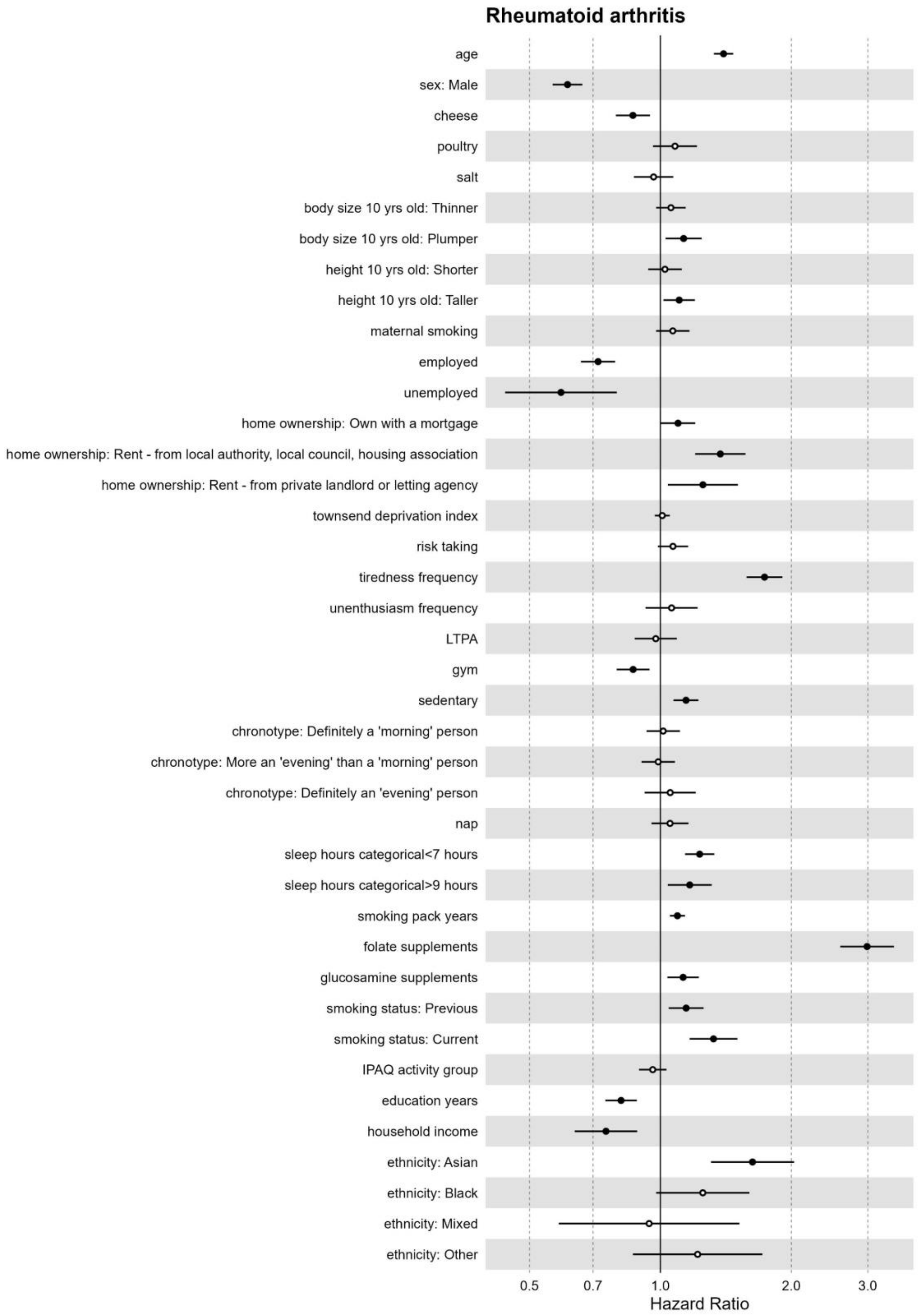

Figure S56. Effect estimates from the full exposome multivariable Cox model for Rheumatoid arthritis in UK Biobank Participants recruited in England (n=436,891).

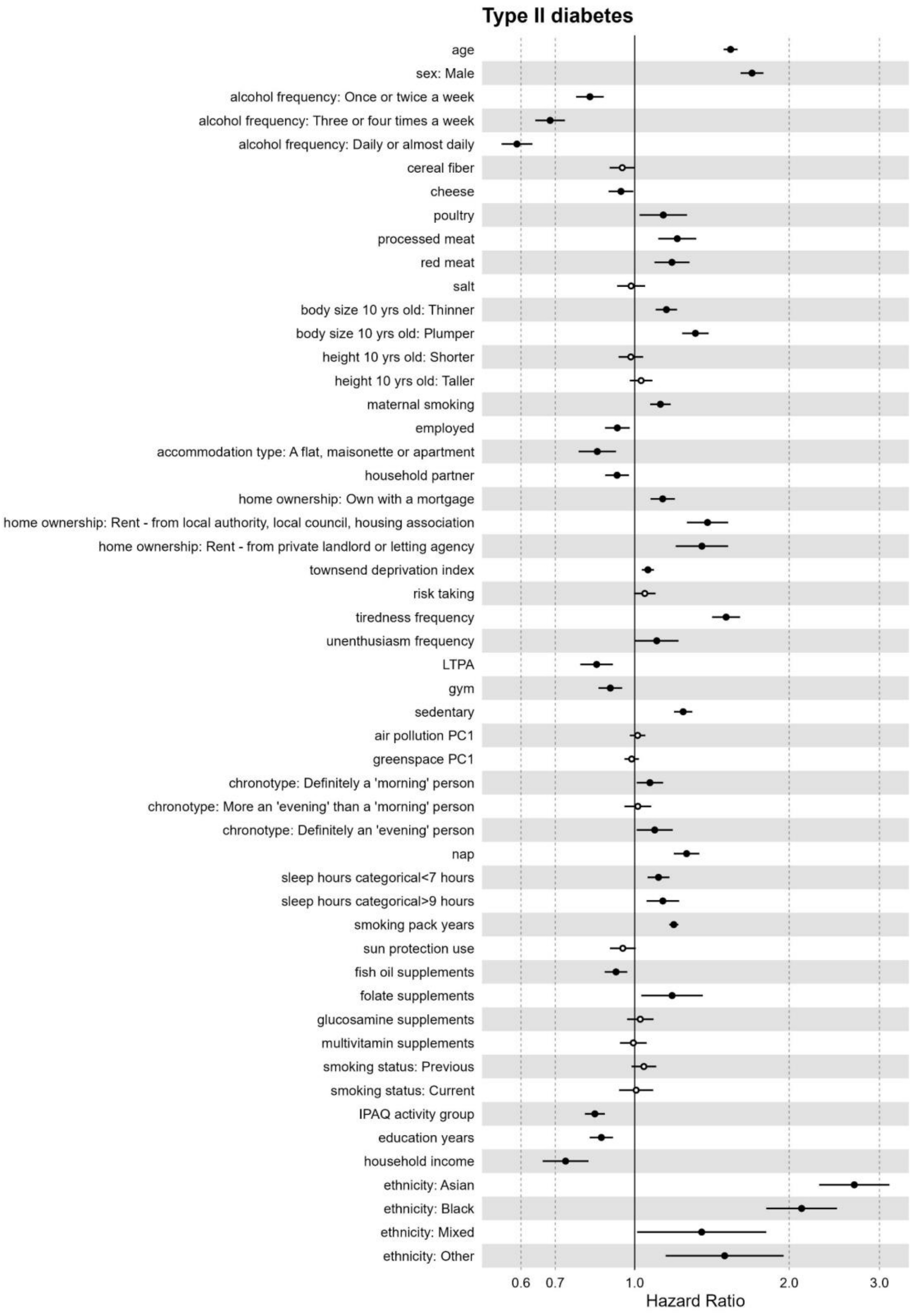

Figure S57. Effect estimates from the full exposome multivariable Cox model for type 2 diabetes in UK Biobank Participants recruited in England (n=436,891).

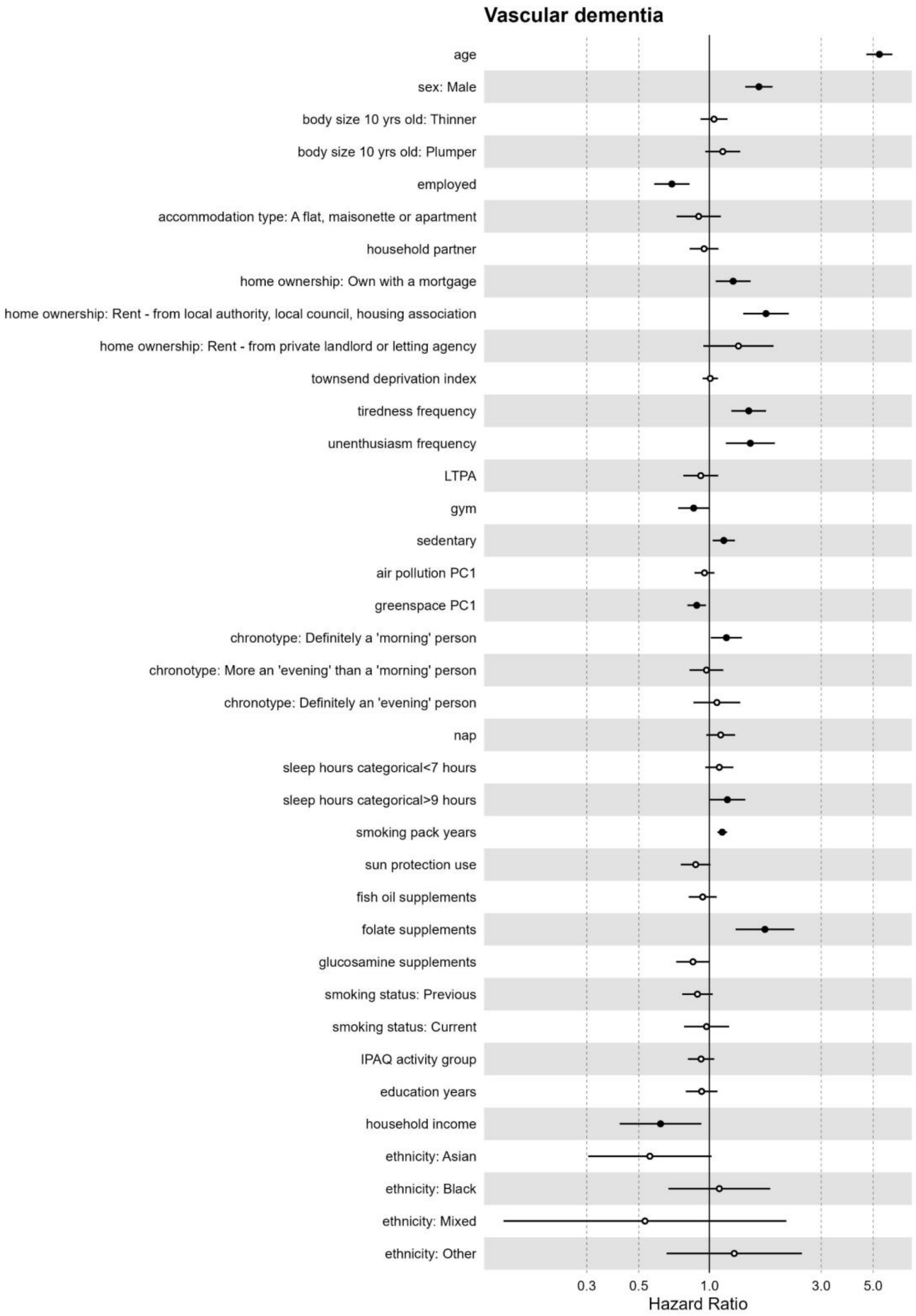

Figure S58. Effect estimates from the full exposome multivariable Cox model for vascular dementia in UK Biobank Participants recruited in England (n=436,891).

### Supplementary tables

**Table S1**. Baseline descriptive statistics - UK Biobank participants recruited in England

| | Female (N=237,634) | Male (N=199,257) | Total (N=436,891) |
|---|---|---|---|
| **Age** | | | |
| Mean (SD) | 56 (8.0) | 57 (8.2) | 57 (8.1) |
| **Household income** | | | |
| Less than 18,000 | 52,139 (21.9%) | 38,416 (19.3%) | 90,555 (20.7%) |
| 18,000 to 30,999 | 58,496 (24.6%) | 45,827 (23.0%) | 104,323 (23.9%) |
| 31,000 to 51,999 | 52,229 (22.0%) | 48,178 (24.2%) | 100,407 (23.0%) |
| 52,000 to 100,000 | 37,443 (15.8%) | 39,514 (19.8%) | 76,957 (17.6%) |
| Greater than 100,000 | 9,742 (4.1%) | 10,884 (5.5%) | 20,626 (4.7%) |
| **Education years** | | | |
| 7 years | 39,642 (16.7%) | 33,716 (16.9%) | 73,358 (16.8%) |
| 10 years | 46,951 (19.8%) | 27,632 (13.9%) | 74,583 (17.1%) |
| 13 years | 13,922 (5.9%) | 10,134 (5.1%) | 24,056 (5.5%) |
| 15 years | 31,779 (13.4%) | 20,463 (10.3%) | 52,242 (12.0%) |
| 19 years | 30,058 (12.6%) | 38,388 (19.3%) | 68,446 (15.7%) |
| 20 years | 72,867 (30.7%) | 66,742 (33.5%) | 139,609 (32.0%) |
| **Ethnicity** | | | |
| White | 223,428 (94.0%) | 187,256 (94.0%) | 410,684 (94.0%) |
| Asian | 5,172 (2.2%) | 5,344 (2.7%) | 10,516 (2.4%) |
| Black | 4,452 (1.9%) | 3,210 (1.6%) | 7,662 (1.8%) |
| Mixed | 1,610 (0.7%) | 938 (0.5%) | 2,548 (0.6%) |
| Other | 2,388 (1.0%) | 1,737 (0.9%) | 4,125 (0.9%) |
| **BMI** | | | |
| Mean (SD) | 27 (5.2) | 28 (4.2) | 27 (4.8) |
| **Smoking status** | | | |
| Never | 141,414 (59.5%) | 97,119 (48.7%) | 238,533 (54.6%) |
| Previous | 74,753 (31.5%) | 77,122 (38.7%) | 151,875 (34.8%) |
| Current | 20,591 (8.7%) | 24,223 (12.2%) | 44,814 (10.3%) |
| **Home area population density** | | | |
| Urban | 203,583 (85.7%) | 171,299 (86.0%) | 374,882 (85.8%) |
| Rural | 34,051 (14.3%) | 27,958 (14.0%) | 62,009 (14.2%) |
| **Mortality** | | | |
| Alive | 224,740 (94.6%) | 180,435 (90.6%) | 405,175 (92.7%) |
| Dead | 12,894 (5.4%) | 18,822 (9.4%) | 31,716 (7.3%) |

Mortality rates are for the 11-15 year study follow up period. Descriptive statistics are calculated using the first imputed analysis dataset and are not pooled across imputed datasets. BMI: body mass index; SD: standard deviation.

| **Table S2.** Baseline descriptive statistics - UK Biobank participants with no prevalent disease | | | |
|---|---|---|---|
| | Female (N=163,415) | Male (N=137,905) | Total (N=301,320) |
| **Age** | | | |
| Mean (SD) | 55 (8.0) | 55 (8.2) | 55 (8.1) |
| **Household income** | | | |
| Less than 18,000 | 29,254 (17.9%) | 20,104 (14.6%) | 49,358 (16.4%) |
| 18,000 to 30,999 | 39,009 (23.9%) | 30,209 (21.9%) | 69,218 (23.0%) |
| 31,000 to 51,999 | 39,122 (23.9%) | 36,101 (26.2%) | 75,223 (25.0%) |
| 52,000 to 100,000 | 30,113 (18.4%) | 31,891 (23.1%) | 62,004 (20.6%) |
| Greater than 100,000 | 7,984 (4.9%) | 9,057 (6.6%) | 17,041 (5.7%) |
| **Education years** | | | |
| 7 years | 21,857 (13.4%) | 18,072 (13.1%) | 39,929 (13.3%) |
| 10 years | 32,872 (20.1%) | 19,198 (13.9%) | 52,070 (17.3%) |
| 13 years | 10,116 (6.2%) | 7,321 (5.3%) | 17,437 (5.8%) |
| 15 years | 21,435 (13.1%) | 13,934 (10.1%) | 35,369 (11.7%) |
| 19 years | 21,210 (13.0%) | 27,055 (19.6%) | 48,265 (16.0%) |
| 20 years | 54,466 (33.3%) | 51,046 (37.0%) | 105,512 (35.0%) |
| **Ethnicity** | | | |
| White | 153,607 (94.0%) | 129,698 (94.0%) | 283,305 (94.0%) |
| Asian | 3,481 (2.1%) | 3,416 (2.5%) | 6,897 (2.3%) |
| Black | 3,095 (1.9%) | 2,343 (1.7%) | 5,438 (1.8%) |
| Mixed | 1,185 (0.7%) | 696 (0.5%) | 1,881 (0.6%) |
| Other | 1,647 (1.0%) | 1,245 (0.9%) | 2,892 (1.0%) |
| **BMI** | | | |
| Mean (SD) | 26 (4.8) | 27 (3.9) | 27 (4.4) |
| **Smoking status** | | | |
| Never | 100,764 (61.7%) | 72,787 (52.8%) | 173,551 (57.6%) |
| Previous | 48,831 (29.9%) | 48,644 (35.3%) | 97,475 (32.3%) |
| Current | 13,300 (8.1%) | 16,046 (11.6%) | 29,346 (9.7%) |
| **Home area population density** | | | |
| Urban | 139,489 (85.4%) | 117,969 (85.5%) | 257,458 (85.4%) |
| Rural | 23,926 (14.6%) | 19,936 (14.5%) | 43,862 (14.6%) |
| **Mortality** | | | |
| Alive | 157,783 (96.6%) | 129,820 (94.1%) | 287,603 (95.4%) |
| Dead | 5,632 (3.4%) | 8,085 (5.9%) | 13,717 (4.6%) |

Baseline descriptive statistics for the subsample of UK Biobank participants with no disease at baseline (used for disease sensitivity analyses). Mortality rates are for the 11-15 year study follow up period. Descriptive statistics are calculated using the first imputed analysis dataset and are not pooled across imputed datasets. SD: standard deviation.

**Table S3.** Baseline descriptive statistics - UK Biobank participants recruited in Scotland/Wales

| | Female (N=30,707) | Male (N=24,969) | Total (N=55,676) |
|---|---|---|---|
| **Age** | | | |
| Mean (SD) | 56 (8.0) | 57 (8.1) | 56 (8.0) |
| **Household income** | | | |
| Less than 18,000 | 7,454 (24.3%) | 4,843 (19.4%) | 12,297 (22.1%) |
| 18,000 to 30,999 | 7,060 (23.0%) | 5,571 (22.3%) | 12,631 (22.7%) |
| 31,000 to 51,999 | 7,018 (22.9%) | 6,370 (25.5%) | 13,388 (24.0%) |
| 52,000 to 100,000 | 5,037 (16.4%) | 5,258 (21.1%) | 10,295 (18.5%) |
| Greater than 100,000 | 1,127 (3.7%) | 1,307 (5.2%) | 2,434 (4.4%) |
| **Education years** | | | |
| 7 years | 5,571 (18.1%) | 4,669 (18.7%) | 10,240 (18.4%) |
| 10 years | 5,000 (16.3%) | 2,828 (11.3%) | 7,828 (14.1%) |
| 13 years | 1,893 (6.2%) | 1,391 (5.6%) | 3,284 (5.9%) |
| 15 years | 3,520 (11.5%) | 2,453 (9.8%) | 5,973 (10.7%) |
| 19 years | 3,466 (11.3%) | 4,289 (17.2%) | 7,755 (13.9%) |
| 20 years | 10,966 (35.7%) | 9,104 (36.5%) | 20,070 (36.0%) |
| **Ethnicity** | | | |
| White | 30,101 (98.0%) | 24,383 (97.7%) | 54,484 (97.9%) |
| Asian | 247 (0.8%) | 266 (1.1%) | 513 (0.9%) |
| Black | 73 (0.2%) | 72 (0.3%) | 145 (0.3%) |
| Mixed | 125 (0.4%) | 86 (0.3%) | 211 (0.4%) |
| Other | 114 (0.4%) | 104 (0.4%) | 218 (0.4%) |
| **BMI** | | | |
| Mean (SD) | 27 (5.3) | 28 (4.3) | 28 (4.9) |
| **Smoking status** | | | |
| Never | 18,264 (59.5%) | 12,575 (50.4%) | 30,839 (55.4%) |
| Previous | 9,221 (30.0%) | 8,845 (35.4%) | 18,066 (32.4%) |
| Current | 3,128 (10.2%) | 3,476 (13.9%) | 6,604 (11.9%) |
| **Home area population density** | | | |
| Urban | 27,481 (89.5%) | 22,310 (89.4%) | 49,791 (89.4%) |
| Rural | 3,219 (10.5%) | 2,648 (10.6%) | 5,867 (10.5%) |
| **Mortality** | | | |
| Alive | 28,518 (92.9%) | 21,891 (87.7%) | 50,409 (90.5%) |
| Dead | 2,189 (7.1%) | 3,078 (12.3%) | 5,267 (9.5%) |

Mortality rates are for the 11-15 year study follow up period. Descriptive statistics are calculated using the first imputed analysis dataset and are not pooled across imputed datasets. SD: standard deviation.

**Table S4.** Mortality by cause of death - UK Biobank participants recruited in England

| | Total deaths (N=31,716) |
|---|---|
| All diseases* | 22,201 (70.0%) |
| Ischemic heart disease | 5,320 (16.8%) |
| Lung cancer | 2,845 (9.0%) |
| Cerebrovascular diseases | 2,349 (7.4%) |
| Emphysema, COPD | 2,266 (7.1%) |
| Colorectal cancer | 1,624 (5.1%) |
| Breast cancer | 1,404 (4.4%) |
| Type II diabetes | 1,394 (4.4%) |
| All-cause dementia | 1,281 (4.0%) |
| Pancreatic cancer | 1,228 (3.9%) |
| Prostate cancer | 1,216 (3.8%) |
| Chronic kidney diseases | 1,160 (3.7%) |
| Esophageal cancer | 816 (2.6%) |
| Lymphoma | 764 (2.4%) |
| Parkinson's disease | 722 (2.3%) |
| Chronic liver diseases | 776 (2.4%) |
| Alzheimer's disease | 661 (2.1%) |
| Leukemia | 656 (2.1%) |
| Ovarian cancer | 590 (1.9%) |
| Liver cancer | 580 (1.8%) |
| Vascular dementia | 362 (1.1%) |
| Rheumatoid arthritis | 193 (0.6%) |
| Osteoporosis | 63 (0.2%) |
| Osteoarthritis | 36 (0.1%) |
| Macular degeneration | 1 (0.0%) |

**Table S5.** Mortality by cause of death - UK Biobank participants recruited in Scotland/Wales

| | Total deaths (N=5,267) |
|---|---|
| All diseases* | 3,762 (71.4%) |
| Ischemic heart disease | 936 (17.8%) |
| Lung cancer | 558 (10.6%) |
| Cerebrovascular diseases | 434 (8.2%) |
| Emphysema, COPD | 400 (7.6%) |
| Type II diabetes | 323 (6.1%) |
| Colorectal cancer | 276 (5.2%) |
| All-cause dementia | 251 (4.8%) |
| Breast cancer | 216 (4.1%) |
| Pancreatic cancer | 195 (3.7%) |
| Alzheimer's disease | 181 (3.4%) |
| Chronic kidney diseases | 178 (3.4%) |
| Prostate cancer | 163 (3.1%) |
| Esophageal cancer | 122 (2.3%) |
| Chronic liver diseases | 129 (2.4%) |
| Lymphoma | 112 (2.1%) |
| Parkinson's disease | 106 (2.0%) |
| Liver cancer | 103 (2.0%) |
| Ovarian cancer | 97 (1.8%) |
| Vascular dementia | 94 (1.8%) |
| Leukemia | 91 (1.7%) |
| Rheumatoid arthritis | 46 (0.9%) |
| Osteoporosis | 12 (0.2%) |
| Osteoarthritis | 3 (0.1%) |
| Macular degeneration | 0 (0%) |

Numbers and percentages represent the number of deaths for which each disease was listed as either the primary or contributory cause of death. Only diseases that were associated with at least one exposure are listed.

*All diseases encompasses only those diseases listed in this table and is an indicator of the number of participants with any of the diseases in this table listed as a primary or contributory cause of death

**Table S6.** Chronic disease and clinical risk factor diagnosis rates - UK Biobank participants recruited in England

| | Female (N=237,634) | Male (N=199,257) | Total (N=436,891) |
|---|---|---|---|
| **Colorectal cancer** | | | |
| No diagnosis | 233,700 (98.3%) | 194,334 (97.5%) | 428,034 (98.0%) |
| Incident diagnosis | 2,834 (1.2%) | 3,516 (1.8%) | 6,350 (1.5%) |
| Prevalent diagnosis | 1,100 (0.5%) | 1,407 (0.7%) | 2,507 (0.6%) |
| **Lung cancer** | | | |
| No diagnosis | 235,068 (98.9%) | 196,424 (98.6%) | 431,492 (98.8%) |
| Incident diagnosis | 2,305 (1.0%) | 2,423 (1.2%) | 4,728 (1.1%) |
| Prevalent diagnosis | 261 (0.1%) | 410 (0.2%) | 671 (0.2%) |
| **Esophageal cancer** | | | |
| No diagnosis | 237,210 (99.8%) | 198,099 (99.4%) | 435,309 (99.6%) |
| Incident diagnosis | 373 (0.2%) | 986 (0.5%) | 1,359 (0.3%) |
| Prevalent diagnosis | 51 (0.0%) | 172 (0.1%) | 223 (0.1%) |
| **Liver cancer** | | | |
| No diagnosis | 237,178 (99.8%) | 198,576 (99.7%) | 435,754 (99.7%) |
| Incident diagnosis | 382 (0.2%) | 570 (0.3%) | 952 (0.2%) |
| Prevalent diagnosis | 74 (0.0%) | 111 (0.1%) | 185 (0.0%) |
| **Pancreatic cancer** | | | |
| No diagnosis | 236,855 (99.7%) | 198,358 (99.5%) | 435,213 (99.6%) |
| Incident diagnosis | 736 (0.3%) | 843 (0.4%) | 1,579 (0.4%) |
| Prevalent diagnosis | 43 (0.0%) | 56 (0.0%) | 99 (0.0%) |
| **Brain cancer** | | | |
| No diagnosis | 237,133 (99.8%) | 198,668 (99.7%) | 435,801 (99.8%) |
| Incident diagnosis | 379 (0.2%) | 477 (0.2%) | 856 (0.2%) |
| Prevalent diagnosis | 122 (0.1%) | 112 (0.1%) | 234 (0.1%) |
| **Leukemia** | | | |
| No diagnosis | 236,728 (99.6%) | 197,946 (99.3%) | 434,674 (99.5%) |
| Incident diagnosis | 692 (0.3%) | 1,020 (0.5%) | 1,712 (0.4%) |
| Prevalent diagnosis | 214 (0.1%) | 291 (0.1%) | 505 (0.1%) |
| **Lymphoma** | | | |
| No diagnosis | 235,985 (99.3%) | 197,254 (99.0%) | 433,239 (99.2%) |
| Incident diagnosis | 1,201 (0.5%) | 1,429 (0.7%) | 2,630 (0.6%) |
| Prevalent diagnosis | 448 (0.2%) | 574 (0.3%) | 1,022 (0.2%) |
| **Breast cancer** | | | |
| No diagnosis | 218,845 (92.1%) | 199,105 (99.9%) | 417,950 (95.7%) |
| Incident diagnosis | 8,843 (3.7%) | 90 (0.0%) | 8,933 (2.0%) |
| Prevalent diagnosis | 9,946 (4.2%) | 62 (0.0%) | 10,008 (2.3%) |
| **Ovarian cancer** | | | |
| No diagnosis | 235,622 (99.2%) | 199,254 (100.0%) | 434,876 (99.5%) |
| Incident diagnosis | 1,190 (0.5%) | 3 (0.0%) | 1,193 (0.3%) |
| Prevalent diagnosis | 822 (0.3%) | 0 (0%) | 822 (0.2%) |

| **Table S6.** Chronic disease and clinical risk factor diagnosis rates - UK Biobank participants recruited in England | | | |
|---|---|---|---|
| | Female (N=237,634) | Male (N=199,257) | Total (N=436,891) |
| **Prostate cancer** | | | |
| No diagnosis | 237,626 (100.0%) | 186,308 (93.5%) | 423,934 (97.0%) |
| Incident diagnosis | 5 (0.0%) | 9,805 (4.9%) | 9,810 (2.2%) |
| Prevalent diagnosis | 3 (0.0%) | 3,144 (1.6%) | 3,147 (0.7%) |
| **Type II diabetes** | | | |
| No diagnosis | 220,185 (92.7%) | 174,531 (87.6%) | 394,716 (90.3%) |
| Incident diagnosis | 6,838 (2.9%) | 8,592 (4.3%) | 15,430 (3.5%) |
| Prevalent diagnosis | 10,611 (4.5%) | 16,134 (8.1%) | 26,745 (6.1%) |
| **Ischemic heart disease** | | | |
| No diagnosis | 218,285 (91.9%) | 163,606 (82.1%) | 381,891 (87.4%) |
| Incident diagnosis | 11,852 (5.0%) | 19,369 (9.7%) | 31,221 (7.1%) |
| Prevalent diagnosis | 7,497 (3.2%) | 16,282 (8.2%) | 23,779 (5.4%) |
| **Cerebrovascular diseases** | | | |
| No diagnosis | 227,488 (95.7%) | 186,034 (93.4%) | 413,522 (94.7%) |
| Incident diagnosis | 6,765 (2.8%) | 8,640 (4.3%) | 15,405 (3.5%) |
| Prevalent diagnosis | 3,381 (1.4%) | 4,583 (2.3%) | 7,964 (1.8%) |
| **Emphysema, COPD** | | | |
| No diagnosis | 226,215 (95.2%) | 186,023 (93.4%) | 412,238 (94.4%) |
| Incident diagnosis | 6,970 (2.9%) | 8,485 (4.3%) | 15,455 (3.5%) |
| Prevalent diagnosis | 4,449 (1.9%) | 4,749 (2.4%) | 9,198 (2.1%) |
| **Chronic liver diseases** | | | |
| No diagnosis | 233,239 (98.2%) | 194,463 (97.6%) | 427,702 (97.9%) |
| Incident diagnosis | 3,621 (1.5%) | 3,793 (1.9%) | 7,414 (1.7%) |
| Prevalent diagnosis | 774 (0.3%) | 1,001 (0.5%) | 1,775 (0.4%) |
| **Chronic kidney diseases** | | | |
| No diagnosis | 228,299 (96.1%) | 189,208 (95.0%) | 417,507 (95.6%) |
| Incident diagnosis | 8,858 (3.7%) | 9,381 (4.7%) | 18,239 (4.2%) |
| Prevalent diagnosis | 477 (0.2%) | 668 (0.3%) | 1,145 (0.3%) |
| **All-cause dementia** | | | |
| No diagnosis | 234,741 (98.8%) | 195,953 (98.3%) | 430,694 (98.6%) |
| Incident diagnosis | 2,819 (1.2%) | 3,214 (1.6%) | 6,033 (1.4%) |
| Prevalent diagnosis | 74 (0.0%) | 90 (0.0%) | 164 (0.0%) |
| **Vascular dementia** | | | |
| No diagnosis | 236,967 (99.7%) | 198,308 (99.5%) | 435,275 (99.6%) |
| Incident diagnosis | 614 (0.3%) | 884 (0.4%) | 1,498 (0.3%) |
| Prevalent diagnosis | 53 (0.0%) | 65 (0.0%) | 118 (0.0%) |
| **Alzheimer's** | | | |
| No diagnosis | 236,139 (99.4%) | 197,823 (99.3%) | 433,962 (99.3%) |
| Incident diagnosis | 1,441 (0.6%) | 1,368 (0.7%) | 2,809 (0.6%) |
| Prevalent diagnosis | 54 (0.0%) | 66 (0.0%) | 120 (0.0%) |

**Table S6.** Chronic disease and clinical risk factor diagnosis rates - UK Biobank participants recruited in England

| | Female (N=237,634) | Male (N=199,257) | Total (N=436,891) |
|---|---|---|---|
| **Parkinson's** | | | |
| No diagnosis | 236,302 (99.4%) | 197,026 (98.9%) | 433,328 (99.2%) |
| Incident diagnosis | 1,035 (0.4%) | 1,719 (0.9%) | 2,754 (0.6%) |
| Prevalent diagnosis | 297 (0.1%) | 512 (0.3%) | 809 (0.2%) |
| **Rheumatoid arthritis** | | | |
| No diagnosis | 231,489 (97.4%) | 196,363 (98.5%) | 427,852 (97.9%) |
| Incident diagnosis | 2,668 (1.1%) | 1,362 (0.7%) | 4,030 (0.9%) |
| Prevalent diagnosis | 3,477 (1.5%) | 1,532 (0.8%) | 5,009 (1.1%) |
| **Macular degeneration** | | | |
| No diagnosis | 232,478 (97.8%) | 195,968 (98.3%) | 428,446 (98.1%) |
| Incident diagnosis | 4,625 (1.9%) | 2,973 (1.5%) | 7,598 (1.7%) |
| Prevalent diagnosis | 531 (0.2%) | 316 (0.2%) | 847 (0.2%) |
| **Osteoporosis** | | | |
| No diagnosis | 222,980 (93.8%) | 196,689 (98.7%) | 419,669 (96.1%) |
| Incident diagnosis | 8,307 (3.5%) | 1,686 (0.8%) | 9,993 (2.3%) |
| Prevalent diagnosis | 6,347 (2.7%) | 882 (0.4%) | 7,229 (1.7%) |
| **Osteoarthritis** | | | |
| No diagnosis | 183,497 (77.2%) | 162,615 (81.6%) | 346,112 (79.2%) |
| Incident diagnosis | 26,469 (11.1%) | 19,410 (9.7%) | 45,879 (10.5%) |
| Prevalent diagnosis | 27,668 (11.6%) | 17,232 (8.6%) | 44,900 (10.3%) |
| **Hypertension** | | | |
| No diagnosis | 45,992 (19.4%) | 28,391 (14.2%) | 74,383 (17.0%) |
| Prevalent diagnosis | 191,642 (80.6%) | 170,866 (85.8%) | 362,508 (83.0%) |
| **Obesity** | | | |
| No diagnosis | 181,877 (76.5%) | 148,720 (74.6%) | 330,597 (75.7%) |
| Prevalent diagnosis | 55,757 (23.5%) | 50,537 (25.4%) | 106,294 (24.3%) |
| **Dyslipidemia** | | | |
| No diagnosis | 108,118 (45.5%) | 67,006 (33.6%) | 175,124 (40.1%) |
| Prevalent diagnosis | 129,516 (54.5%) | 132,251 (66.4%) | 261,767 (59.9%) |

Prevalent chronic disease rates are calculated as those with a corresponding ICD diagnosis date before or on the date of recruitment into the UK Biobank cohort. Additionally, those who self-reported a physician diagnosis of cancer, diabetes, heart attack, stroke, or bronchitis/emphysema during the baseline verbal interview were also used to count prevalent cases for cancer, diabetes, heart disease, cerebrovascular disease, and chronic lower respiratory diseases, respectively. Incident disease rates are for the 11-14 year study follow up period and exclude those with prevalent disease at baseline. Descriptive statistics are calculated using the first imputed analysis dataset and are not pooled across imputed datasets.

**Table S7.** Biomarker summary statistics by sex - UK Biobank participants recruited in England

| | Female (N=237,634) | Male (N=199,257) | Total (N=436,891) |
|---|---|---|---|
| Alanine Aminotransferase (U/L) | 20 (12) | 27 (15) | 23 (14) |
| Albumin (g/L) | 45 (2.6) | 46 (2.6) | 45 (2.6) |
| Alkaline phosphatase (U/L) | 85 (27) | 82 (25) | 84 (26) |
| Apolipoprotein A (g/L) | 1.6 (0.27) | 1.4 (0.23) | 1.5 (0.28) |
| Apolipoprotein B (g/L) | 1.0 (0.24) | 1.0 (0.24) | 1.0 (0.24) |
| Aspartate aminotransferase (U/L) | 24 (9.4) | 28 (12) | 26 (11) |
| C-reactive protein (mg/L) | 2.7 (4.4) | 2.5 (4.4) | 2.6 (4.4) |
| Cholesterol (mmol/L) | 5.9 (1.1) | 5.5 (1.1) | 5.7 (1.1) |
| Creatinine (umol/L) | 64 (14) | 82 (19) | 72 (19) |
| Cystatin C (mg/L) | 0.88 (0.16) | 0.94 (0.18) | 0.91 (0.18) |
| Direct bilirubin (umol/L) | 1.5 (0.64) | 2.0 (0.93) | 1.7 (0.82) |
| Gamma glutamyltransferase (U/L) | 30 (34) | 46 (50) | 37 (42) |
| Glucose (mmol/L) | 5.1 (1.1) | 5.2 (1.4) | 5.1 (1.2) |
| Glycated haemoglobin (HbA1c) (mmol/mol) | 36 (5.9) | 37 (7.6) | 36 (6.7) |
| HDL cholesterol | 1.6 (0.38) | 1.3 (0.31) | 1.5 (0.39) |
| Insulin-like growth factor 1 (IGF-1) (nmol/L) | 21 (5.8) | 22 (5.6) | 21 (5.7) |
| LDL direct (mmol/L) | 3.6 (0.87) | 3.5 (0.86) | 3.6 (0.87) |
| Leukocyte telomere length (T/S ratio) | 0.84 (0.13) | 0.82 (0.13) | 0.83 (0.13) |
| Lipoprotein(a) (nmol/L) | 45 (50) | 44 (49) | 45 (49) |
| Phosphate (mmol/L) | 1.2 (0.15) | 1.1 (0.16) | 1.2 (0.16) |
| Total bilirubin (umol/L) | 8.1 (3.7) | 10 (4.9) | 9.1 (4.4) |
| Triglycerides (mmol/L) | 1.5 (0.85) | 2.0 (1.2) | 1.7 (1.0) |
| Urate (umol/L) | 270 (66) | 360 (72) | 310 (81) |
| Urea (mmol/L) | 5.2 (1.3) | 5.6 (1.5) | 5.4 (1.4) |
| Vitamin D (nmol/L) | 49 (21) | 49 (21) | 49 (21) |

**Table S8.** Variables used for biomarker analyses

| Biomarker | UK Biobank field ID |
|---|---|
| Alanine aminotransferase | 30620 |
| Albumin | 30600 |
| Alkaline phosphatase | 30610 |
| Apolipoprotein A | 30630 |
| Apolipoprotein B | 30640 |
| Aspartate aminotransferase | 30650 |
| High sensitivity C-reactive protein | 30710 |
| Cholesterol | 30690 |
| Creatinine | 30700 |
| Cystatin C | 30720 |
| Direct bilirubin | 30660 |
| Total bilirubin | 30840 |
| Gamma glutamyltransferase | 30730 |
| Glucose | 30740 |
| Glycated hemoglobin (hbA1c) | 30750 |
| HDL cholesterol | 30760 |
| Insulin-like growth factor 1 (IGF-1) | 30770 |
| LDL direct | 30780 |
| Lipoprotein A | 30790 |
| Phosphate | 30810 |
| Triglycerides | 30870 |
| Urate | 30880 |
| Urea | 30670 |
| Vitamin D | 30890 |
| Leukocyte telomere length (LTL) | 22192 |

**Table S9.** Variables used to calculate prevalence and incidence of chronic diseases and clinical risk factors

| **Chronic disease** | **Baseline measures (field ID)** | **Baseline verbal interview diagnosis codes** | **ICD-10 codes** | **ICD-9 codes** |
|---|---|---|---|---|
| Colorectal cancer | - | 1022, 1023 | C18-C20 | 153, 154 |
| Lung cancer | - | 1001, 1027, 1028, 1080 | C33, C34 | 162 |
| Esophageal cancer | - | 1017 | C15 | 150 |
| Liver cancer | - | 1024 | C22 | 155 |
| Pancreatic cancer | - | 1026 | C25 | 157 |
| Brain cancer | - | 1032 | C71 | 191 |
| Leukemia | - | 1048 | C91-C95 | 204-208 |
| Lymphoma | - | 1047 | C81-C86, C88 | 201-203 |
| Breast cancer | - | 1002 | C50 | 174-175 |
| Ovarian cancer | - | 1039 | C56 | 183 |
| Prostate cancer | - | 1044 | C61 | 185 |
| Type 2 diabetes | Taking insulin medication (6153, 6177)<br>Diabetes diagnosed by physician (2443)<br>Non-fasting blood hbA1c $\geq$ 48 mmol/mol (30750)<br>Non-fasting blood glucose $\geq$ 11.1 mmol/L (30740) | 1223 | E11 | 250 |
| Ischemic heart disease | Heart attack diagnosis by physician (6150)<br>Angina diagnosis by physician (6150) | 1074, 1075 | I20-I25 | 410-414 |
| Cerebrovascular diseases | Stroke diagnosis by physician (6150) | 1081, 1086, 1491, 1583 | I60-I69 | 430-438 |
| Emphysema, COPD | Bronchitis/emphysema diagnosis by physician (6152) | 1112, 1472 | J43-J44 | 492 |
| Chronic liver diseases | - | 1157, 1158, 1604 | K70, K73-K74, K75.8, K76.0 | 571 |
| Chronic kidney diseases | - | 1192, 1193, 1194 | N18 | 585 |

| | | | | |
|---|---|---|---|---|
| All-cause dementia | - | 1263 | A81.0, F00-F03, F05.1, F10.6, G30-G31, I67.3 | 331.0, 290.4, 331.1, 290.2, 290.3, 291.2, 294.1, 331.2, 331.5 |
| Vascular dementia | - | 1263 | F01, I67.3 | 290.4 |
| Alzheimer's disease | - | 1263 | F00, G30 | 331.0 |
| Parkinson's disease and parkinsonism | - | 1262 | G20-G22 | 332 |
| Rheumatoid arthritis | - | 1464 | M05-M06 | 714 |
| Macular degeneration | - | 1528 | H35.3 | 362.5 |
| Osteoporosis | - | 1309 | M80-M81 | 733.0 |
| Osteoarthritis | - | 1465 | M15-M19 | 715 |
| **Clinical risk factors** | **Baseline measures (field ID)** | **Baseline verbal interview diagnosis codes** | **ICD-10 codes** | **ICD-9 codes** |
| Hypertension | High blood pressure diagnosis by physician (6150)<br>Taking medication for high blood pressure (6153, 6177)<br>Blood pressure SBP/DBP ≥ 140/90 mmHg (4079, 4080) | 1065, 1072 | I10-I15 | 401-405 |
| Obesity | BMI ≥ 30 $kg/m^2$ (21001) | - | E66 | 278.0 |
| Dyslipidemia | Taking cholesterol lowering medication (6153, 6177)<br>Blood cholesterol ≥ 240 / 38.67 (30690)<br>Blood LDL ≥ 160 / 38.67 (30780)<br>Blood HDL < 40 / 38.67 (30760)<br>Blood triglycerides ≥ 200 / 88.57 (30870) | - | E78 | 272 |

Verbal interview diagnosis codes are contained in the cancer (field ID 20001) and non-cancer illness (field ID 20002) variables. Field IDs for ICD variables: ICD-10 summary diagnoses (41270) and date of diagnosis (41280); ICD-9 summary diagnoses (41271) and date of diagnosis (41281). Incident disease cases were also identified using ICD-10 codes from cause of death information from linked death register data. Baseline prevalence for all diseases and clinical risk factors was calculated for all participants using baseline measures (including verbal interview diagnosis codes) + those with an ICD diagnosis before or on the date of recruitment into the UK Biobank. Incident cases are defined as those with an ICD date of diagnosis after the date of recruitment who do not have any prevalent diagnosis. Unless specific ICD subcategories are already given with dot separators, all ICD codes listed also include all subcategories (e.g., J44 includes J44, J44.0, J44.1, J44.8, J44.9). COPD: chronic obstructive pulmonary disease; BMI: body mass index: SBP: systolic blood pressure; DBP: diastolic blood pressure.

**Table S10.** Components and calculation of the daily partial fiber score

| Food variable (UK Biobank field ID) | Portion specified in touchscreen question | Portion size | Estimated fiber content/portion (g) |
|---|---|---|---|
| Bread intake (1438)<br>Bread type (1448) | Slices | White bread: 36 g<br>Brown bread: 36 g<br>Wholemeal bread: 36 g<br>Other type of bread: 36 g | 0.68<br>1.26<br>1.80<br>1.25 |
| Breakfast cereal intake (1458)<br>Breakfast cereal type (1468) | Bowls | Bran cereal: 40 g<br>Biscuit cereal: 40 g<br>Oat cereal: 160 g<br>Muesli: 55 g<br>Other (e.g., cornflakes): 30 g | 7.16<br>2.92<br>1.92<br>4.18<br>0.54 |

Estimated fiber content/portion take from Bradbury et al. (2018) [21].

**Table S11.** Components and calculation of the LTPA and OPA scores

| **LTPA (field ID)** | **METs** |
|---|---|
| Walking for pleasure (6164, 981, 971) | 3.3 METs * mins/week |
| Strenuous sports (6164, 1001, 991) | 8.0 METs * mins/week |
| Other exercises (6164, 3647, 3637) | 4.5 METs * mins/week |
| Light DIY (6164, 1021, 1011) | 2.25 METs * mins/week |
| Heavy DIY (6164, 2634, 2624) | 4.5 METs * mins/week |
| **OPA (field ID)** | |
| Heavy physical work (816, 767) | 4.5 METs * mins/week |
| Walking/standing work (806, 767) | 2.25 METs * mins/week |

LTPA: leisure-time physical activity; OPA: occupational physical activity; MET: metabolic equivalent of task; DIY: do-it-yourself;. METs used for each type of activity were taken from Pearce et al. (2020) [22].

**Table S12.** Variables used for calculation of polygenic risk scores (PRS).

| Outcome | UK Biobank derived PRS measure(s) used (field ID) |
|---|---|
| All-cause mortality | Bowel cancer (26218); breast cancer (26220); ovarian cancer (26232); prostate cancer (26267); type 2 diabetes (26285); cardiovascular disease (26223); coronary artery disease (26227); ischemic stroke (26248); Alzheimer's disease (26206); Parkinson's (26260); rheumatoid arthritis (26273); macular degeneration (26204); osteoporosis (26258); lung cancer; esophageal cancer; pancreatic cancer; leukemia; emphysema/COPD |
| Colorectal cancer | Bowel cancer (26218) |
| Lung cancer | (PGS000078) by Graff et al. (2021) [54] |
| Esophageal cancer | (PGS002298) by Choi et al. (2020) [55] |
| Liver cancer | None |
| Pancreatic cancer | (PGS000083) by Graff et al. (2021) [54] |
| Brain cancer | None |
| Leukemia | (PGS000077) by Graff et al. (2021) [54] |
| Lymphoma | None |
| Breast cancer | Breast cancer (26220) |
| Ovarian cancer | Ovarian cancer (26232) |
| Prostate cancer | Prostate cancer (26267) |
| Type 2 diabetes | Type 2 diabetes (26285) |
| Ischemic heart disease | Cardiovascular disease (26223); coronary artery disease (26227) |
| Cerebrovascular diseases | Ischemic stroke (26248) |
| Emphysema, COPD | (PGS001788) by Wang et al. (2021) [56] |
| Chronic liver diseases | Non-alcoholic fatty liver disease (PGS002282) by Schnurr et al. (2022) [58]<br>Liver cirrhosis (PGS000726) by Emdin et al. (2020) [59] |
| Chronic kidney diseases | (PGS000859) by Mansour Aly et al. (2021) [57] |
| All-cause dementia | Alzheimer's disease (26206) |
| Vascular dementia | Alzheimer's disease (26206) |
| Alzheimer's disease | Alzheimer's disease (26206) |
| Parkinson's disease and parkinsonism | Parkinson's (26260) |
| Rheumatoid arthritis | Rheumatoid arthritis (26273) |
| Macular degeneration | Macular degeneration (26204) |
| Osteoporosis | Osteoporosis (26258) |
| Osteoarthritis | Knee osteoarthritis (PGS002729) by Sedaghati-Khayat et al. (2022) [60] |

| **Table S13.** Accelerometer vs. self-reported physical activity measures in relation to mortality | | | | |
|---|---|---|---|---|
| | **Hazard Ratio [95% CI]** | **p-value** | **Hazard Ratio [95% CI]** | **p-value** |
| Overall acceleration average (milli-gravity) | 0.95 [0.94, 0.95] | < 0.001 | | |
| IPAQ physical activity group | | | 0.82 [0.80, 0.84] | < 0.001 |
| Leisure time physical activity (LTPA) | | | 0.85 [0.82, 0.88] | < 0.001 |
| Occupational physical activity (OPA) | | | 0.87 [0.84, 0.90] | < 0.001 |
| Total sedentary time | | | 1.10 [1.08, 1.13] | < 0.001 |
| $R^2$ | 0.59 | | 0.56 | |

Both models include covariates for age, sex, UK Biobank assessment center, household income, years of education, ethnicity, smoking status, and Townsend deprivation index.

**Table S14.** Smoking associations with prostate cancer according to PSA test

| | No PSA test | | Has PSA test | | PSA test as covariate | |
|---|---|---|---|---|---|---|
| | **Hazard Ratio [95% CI]** | **p-value** | **Hazard Ratio [95% CI]** | **p-value** | **Hazard Ratio [95% CI]** | **p-value** |
| Never smoker (reference) | - | - | - | - | - | - |
| Previous smoker | 0.97 [0.91, 1.03] | 0.31 | 0.93 [0.87, 1.00] | 0.04 | 0.95 [0.91, 1.00] | 0.04 |
| Current smoker | 0.90 [0.81, 0.99] | 0.02 | 0.86 [0.75, 0.98] | 0.02 | 0.88 [0.81, 0.95] | < 0.001 |

All models are Cox models with age as the timescale, stratified by 5-year birth cohorts, and with covariates for UK Biobank assessment center, household income, years of education, ethnicity, and IPAQ activity level. Sample sizes are n=137,598 for those with no PSA test, n=58,425 for those with a PSA test, and n=196,113 for final model not stratified by PSA test but including PSA test as a covariate.

**Table S15.** Explained variation and C-index across multivariable models in UK Biobank participants recruited in England

| Disease | Model 1 C-index | Model 2 C-index | Model 3 C-index | Model 4 C-index | Model 1 $R^2$ | Model 2 $R^2$ | Model 3 $R^2$ | Model 4 $R^2$ | Cases | Sample |
|---|---|---|---|---|---|---|---|---|---|---|
| All-cause mortality | 0.7070 | 0.7138 | 0.7557 | 0.7583 | 0.4300 | 0.4559 | 0.6043 | 0.6134 | 18,885 - 31,716 | 293,043 - 436,891 |
| Vascular dementia | 0.8288 | 0.8472 | 0.8667 | 0.8779 | 0.7890 | 0.8371 | 0.8711 | 0.8983 | 1,102 - 1,498 | 358,524 - 436,773 |
| Emphysema, COPD | 0.6841 | 0.7243 | 0.8432 | 0.8489 | 0.3469 | 0.4818 | 0.8449 | 0.8590 | 9,561 - 16,722 | 289,238 - 428,960 |
| Lung cancer | 0.6959 | 0.7173 | 0.8313 | 0.8326 | 0.3885 | 0.4608 | 0.8332 | 0.8390 | 2,846 - 4,728 | 293,713 - 436,220 |
| All-cause dementia | 0.8095 | 0.8334 | 0.8365 | 0.8552 | 0.7386 | 0.8097 | 0.8095 | 0.8611 | 3,284 - 6,033 | 293,615 - 436,727 |
| Alzheimer's disease | 0.8191 | 0.8578 | 0.8343 | 0.8684 | 0.7620 | 0.8643 | 0.8020 | 0.8879 | 2,196 - 2,809 | 369,360 - 436,771 |
| Chronic kidney diseases | 0.7301 | 0.7366 | 0.7793 | 0.7816 | 0.4954 | 0.5178 | 0.6623 | 0.6692 | 10,150 - 18,239 | 293,178 - 435,746 |
| Parkinson's disease | 0.7638 | 0.7759 | 0.7813 | 0.7942 | 0.6054 | 0.6434 | 0.6617 | 0.6986 | 2,262 - 2,754 | 366,631 - 436,082 |
| Osteoporosis | 0.7536 | 0.7662 | 0.7801 | 0.7912 | 0.5713 | 0.6135 | 0.6568 | 0.6902 | 5,684 - 9,993 | 291,882 - 429,662 |
| Esophageal cancer | 0.7311 | 0.7390 | 0.7716 | 0.7738 | 0.4898 | 0.5199 | 0.6309 | 0.6406 | 1,139 - 1,359 | 368,104 - 436,668 |
| Macular degeneration | 0.7521 | 0.7658 | 0.7612 | 0.7722 | 0.5703 | 0.6216 | 0.5977 | 0.6443 | 6,278 - 7,598 | 369,031 - 436,044 |
| Liver cancer | 0.6943 | - | 0.7428 | - | 0.3798 | - | 0.5651 | - | 824 - 952 | 379,149 - 436,706 |
| Type II diabetes | 0.6230 | 0.7044 | 0.7499 | 0.7812 | 0.1676 | 0.4126 | 0.5611 | 0.6599 | 8,487 - 15,430 | 279,155 - 410,146 |
| Cerebrovascular diseases | 0.7096 | 0.7175 | 0.7408 | 0.7447 | 0.4319 | 0.4583 | 0.5367 | 0.5499 | 12,123 - 15,405 | 351,976 - 428,927 |
| Chronic liver diseases | 0.5512 | 0.5948 | 0.7261 | 0.7370 | 0.0278 | 0.1036 | 0.4979 | 0.5374 | 4,295 - 7,414 | 293,473 - 435,116 |
| Rheumatoid arthritis | 0.6356 | 0.6590 | 0.7254 | 0.7360 | 0.2057 | 0.2720 | 0.4929 | 0.5282 | 3,212 - 4,030 | 359,686 - 431,882 |
| Ischemic heart disease | 0.6837 | 0.7045 | 0.7242 | 0.7392 | 0.3431 | 0.4118 | 0.4716 | 0.5192 | 21,815 - 35,125 | 281,867 - 417,016 |
| Pancreatic cancer | 0.6931 | 0.7106 | 0.7087 | 0.7257 | 0.3798 | 0.4395 | 0.4359 | 0.4898 | 1,489 - 1,579 | 412,225 - 436,792 |
| Leukemia | 0.6897 | 0.7010 | 0.7003 | 0.7098 | 0.3651 | 0.3997 | 0.4000 | 0.4309 | 1,658 - 1,712 | 422,026 - 436,386 |
| Prostate cancer | 0.6774 | 0.7572 | 0.6863 | 0.7602 | 0.3307 | 0.5720 | 0.3573 | 0.5833 | 8,377 - 9,805 | 168,204 - 196,113 |
| Osteoarthritis | 0.6410 | 0.6433 | 0.6803 | 0.6815 | 0.2206 | 0.2272 | 0.3366 | 0.3408 | 36,934 - 45,879 | 324,316 - 391,991 |
| Colorectal cancer | 0.6677 | 0.6982 | 0.6757 | 0.7031 | 0.2927 | 0.3927 | 0.3220 | 0.4125 | 5,983 - 6,350 | 411,358 - 434,384 |
| Lymphoma | 0.6547 | - | 0.6588 | - | 0.2520 | - | 0.2640 | - | 2,595 - 2,630 | 430,959 - 435,869 |
| Ovarian cancer | 0.6061 | 0.6330 | 0.6441 | 0.6663 | 0.1246 | 0.1992 | 0.2334 | 0.3116 | 989 - 1,190 | 200,098 - 236,812 |
| Breast cancer | 0.5416 | 0.6552 | 0.5676 | 0.6628 | 0.0202 | 0.2603 | 0.0549 | 0.2858 | 6,547 - 8,843 | 165,248 - 227,688 |

Model 1: age, sex. Model 2: age, sex, polygenic risk scores (PRS; including genetic principal components, and genotyping batch). Model 3: age, sex, exposome. Model 4: age, sex, exposome, PRS. For diseases, the PRS for that specific disease was added. For all-cause mortality, all PRS for all other diseases in this table were added. If a PRS was not available for a particular outcome, then model 4 was not calculated for that outcome (and a dash is shown). Cases and sample sizes are shown as ranges due to varying levels of missing data across variables used in the different models.

**Table S16.** Explained variation and C-index across multivariable models in UK Biobank participants recruited in Scotland/Wales

| Disease | Model 1 C-index | Model 2 C-index | Model 3 C-index | Model 4 C-index | Model 1 $R^2$ | Model 2 $R^2$ | Model 3 $R^2$ | Model 4 $R^2$ | Cases | Sample |
|---|---|---|---|---|---|---|---|---|---|---|
| All-cause mortality | 0.7190 | 0.7258 | 0.7739 | 0.7741 | 0.4653 | 0.4891 | 0.6554 | 0.6581 | 5,267 | 55,676 |
| Vascular dementia | 0.8619 | 0.8749 | 0.9016 | 0.9084 | 0.8467 | 0.8693 | 0.9154 | 0.9253 | 193 | 55,668 |
| Emphysema, COPD | 0.7008 | 0.7471 | 0.8870 | 0.8920 | 0.3839 | 0.5510 | 0.9143 | 0.9238 | 1,281 | 54,556 |
| All-cause dementia | 0.8243 | 0.8526 | 0.8637 | 0.8802 | 0.7738 | 0.8423 | 0.8677 | 0.9011 | 643 | 55,660 |
| Lung cancer | 0.7036 | 0.7236 | 0.8473 | 0.8494 | 0.4090 | 0.4812 | 0.8612 | 0.8675 | 710 | 55,570 |
| Alzheimer's disease | 0.8194 | 0.8581 | 0.8327 | 0.8640 | 0.7572 | 0.8531 | 0.7960 | 0.8714 | 329 | 55,666 |
| Chronic kidney diseases | 0.7306 | 0.7381 | 0.8120 | 0.8106 | 0.5041 | 0.5260 | 0.7688 | 0.7619 | 966 | 55,505 |
| Esophageal cancer | 0.7450 | 0.7547 | 0.7905 | 0.7954 | 0.5316 | 0.5696 | 0.6979 | 0.7094 | 186 | 55,650 |
| Parkinson's disease | 0.7789 | 0.7981 | 0.7774 | 0.7964 | 0.6596 | 0.7152 | 0.6472 | 0.7022 | 197 | 55,580 |
| Type II diabetes | 0.6440 | 0.7133 | 0.7737 | 0.8107 | 0.2176 | 0.4222 | 0.6360 | 0.7218 | 965 | 52,402 |
| Chronic liver diseases | 0.6093 | 0.6208 | 0.7667 | 0.7762 | 0.1346 | 0.1846 | 0.6095 | 0.6368 | 477 | 55,413 |
| Osteoporosis | 0.7457 | 0.7572 | 0.7647 | 0.7656 | 0.5557 | 0.5879 | 0.6072 | 0.6101 | 112 | 54,629 |
| Macular degeneration | 0.7420 | 0.7495 | 0.7539 | 0.7574 | 0.5398 | 0.5654 | 0.5742 | 0.5926 | 414 | 55,614 |
| Liver cancer | 0.7196 | - | 0.7537 | - | 0.4365 | - | 0.5678 | - | 143 | 55,648 |
| Cerebrovascular diseases | 0.7114 | 0.7216 | 0.7480 | 0.7529 | 0.4466 | 0.4782 | 0.5618 | 0.5753 | 1,549 | 54,508 |
| Ischemic heart disease | 0.6980 | 0.7256 | 0.7390 | 0.7614 | 0.3891 | 0.4827 | 0.5172 | 0.5904 | 3,428 | 52,827 |
| Rheumatoid arthritis | 0.5751 | 0.6421 | 0.7161 | 0.7551 | 0.0855 | 0.2300 | 0.4677 | 0.5846 | 53 | 54,888 |
| Leukemia | 0.7141 | 0.7074 | 0.7213 | 0.7144 | 0.4500 | 0.4447 | 0.4658 | 0.4563 | 176 | 55,609 |
| Prostate cancer | 0.6985 | 0.7735 | 0.6981 | 0.7728 | 0.3827 | 0.6162 | 0.3803 | 0.6142 | 874 | 24,570 |
| Pancreatic cancer | 0.6859 | 0.7070 | 0.6981 | 0.7177 | 0.3390 | 0.3863 | 0.3713 | 0.4159 | 226 | 55,657 |
| Colorectal cancer | 0.6705 | 0.7015 | 0.6790 | 0.7072 | 0.2962 | 0.4036 | 0.3246 | 0.4215 | 760 | 55,315 |
| Osteoarthritis | 0.6380 | 0.6373 | 0.6769 | 0.6764 | 0.2050 | 0.2022 | 0.3072 | 0.3053 | 2,941 | 50,525 |
| Ovarian cancer | 0.6214 | 0.6755 | 0.6552 | 0.6951 | 0.1523 | 0.3034 | 0.2533 | 0.3833 | 154 | 30,590 |
| Lymphoma | 0.6544 | - | 0.6499 | - | 0.2554 | - | 0.2509 | - | 288 | 55,528 |
| Breast cancer | 0.5434 | 0.6447 | 0.5402 | 0.6447 | 0.0185 | 0.2233 | 0.0161 | 0.2225 | 1,042 | 29,447 |

Model 1: age, sex. Model 2: age, sex, polygenic risk scores (PRS; including genetic principal components, and genotyping batch). Model 3: age, sex, exposome. Model 4: age, sex, exposome, PRS. For diseases, the PRS for that specific disease was added. For all-cause mortality, all PRS for all other diseases in this table were added. If a PRS was not available for a particular outcome, then model 4 was not calculated for that outcome (and a dash is shown). Results were calculated using linear predicted values based on model results from the participants recruited in England (n=436,891) and outcome rates from the independent validation sample of participants recruited in Scotland/Wales (n=55,676).

**Supplementary file titles and summaries**

**Supplementary File SF1. Data dictionary for all variables used in multiple imputation.** Summary information about all baseline variables collected from the UK Biobank that were used in multiple imputation after variable exclusions. Information includes variable name used in analysis, UK Biobank field ID, original variable name in UK Biobank dataset provided to us, and URL link for each variable to the corresponding webpage on the UK Biobank showcase giving extensive detail for each variable.

**Supplementary File SF2. Data dictionary for exposures used in XWAS analyses.** Summary information about all exposome analyzed in the mortality XWAS, including the sex-specific reproduction factors analyzed in the sex-specific XWAS only. Information includes variable name used in analysis, UK Biobank field ID, original variable name in UK Biobank dataset provided to us, and URL link for each variable to the corresponding webpage on the UK Biobank showcase giving extensive detail for each variable.

**Supplementary File SF3. Female XWAS output.** We report all female-specific XWAS summary statistics. All effect estimates (hazard ratios, confidence intervals) shown are for analyses in the discovery set (n=118,815). FDR corrected p-values are given for both the discovery and replication analyses (FDR p-values will be NA for variables with an FDR p-value ≥ 0.05 in the discovery analysis, as these variables would not have been tested in the replication stage).

**Supplementary File SF4. Male XWAS output.** We report all male-specific XWAS summary statistics. All effect estimates (hazard ratios, confidence intervals) shown are for analyses in the discovery set (n=99,631). FDR corrected p-values are given for both the discovery and replication analyses (FDR p-values will be NA for variables with an FDR p-value ≥ 0.05 in the discovery analysis, as these variables would not have been tested in the replication stage).

**Supplementary File SF5. Pooled XWAS output.** We report all XWAS summary statistics from the final pooled XWAS. All effect estimates (hazard ratios, confidence intervals) shown are for analyses in the discovery set (n=218,446). FDR corrected p-values are given for both the discovery and replication analyses (FDR p-values will be NA for variables with an FDR p-value ≥ 0.05 in the discovery analysis, as these variables would not have been tested in the replication stage).

**Supplementary File SF6. Disease interaction sensitivity output.** We report all summary statistics from the disease sensitivity analysis conducted among UK Biobank participants where an interaction term was added between each exposure and a binary indicator of poor health at baseline. All effect estimates (hazard ratios, confidence intervals) shown are for analyses in the pooled dataset (n=436,891).

**Supplementary File SF7. XWAS survival time exclusion sensitivity output.** We report all summary statistics from the sensitivity analysis wherein we conducted a mortality XWAS excluding all UK Biobank participants who died within 4 years of baseline (n=431,394). All effect estimates (hazard ratios, confidence intervals) shown are for analyses in the pooled dataset.

**Supplementary Files SF8-SF32. Aging biomarker analysis output.** We report all summary statistics from the aging biomarker analysis testing associations between 25 blood biomarkers and exposures still significant after cluster multivariable and disease sensitivity analyses (n=436,891).

**Supplementary Files SF33-SF60. Incident disease and cardiometabolic risk factor analysis output.** We report all summary statistics from the incident chronic disease and clinical risk factor analysis testing associations between all 28 diseases/risk factors and exposures still significant after cluster multivariable and disease sensitivity analyses (n=436,891).

**Supplementary File SF61. Data dictionary of phenotypes used in phenome-wide association study (PheWAS) analyses.** Summary information about all phenotypes analyzed in per-exposure PheWAS. Information includes variable name used in analysis, UK Biobank field ID, original variable name in UK Biobank dataset provided to us, and URL link for each variable to the corresponding webpage on the UK Biobank showcase giving extensive detail for each variable.

**Supplementary Files SF62-SF177. PheWAS output.** We report all summary statistics from PheWAS testing associations between all exposures still significant after cluster multivariable and disease sensitivity analyses and all baseline phenotypes in the UK Biobank (n=436,891).
